# Probability of vertical HIV transmission: A systematic review and meta-regression

**DOI:** 10.1101/2024.12.03.24318418

**Authors:** Magdalene K. Walters, Michelle A. Bulterys, Michael Barry, Sarah Hicks, Ann Richey, Margalit Sabin, Diana Louden, Mary Mahy, John Stover, Robert Glaubius, Hmwe Kyu, Marie-Claude Boily, Lynne Mofenson, Kathleen Powis, Jeffrey W. Imai-Eaton

**Affiliations:** MRC Centre for Global Infectious Disease Analysis, School of Public Health, Imperial College London, London, United Kingdom; Department of Epidemiology, School of Public Health, University of Washington, Seattle, WA, USA; Herbert Wertheim School of Public Health, University of California San Diego, La Jolla, CA, USA; University Libraries, University of Washington, Seattle, WA, USA; Simmons University, Boston, MA, USA; Data for Impact Department, Joint United Nations Programme on HIV/AIDS, Geneva, Switzerland; Center for Modeling, Planning and Policy Analysis, Avenir Health, Glastonbury, CT, USA; Department of Health Metrics Sciences, School of Medicine, and Institute for Health Metrics and Evaluation, University of Washington, Seattle, WA, USA; Research Program, Elizabeth Glaser Pediatric AIDS Foundation, Washington, DC, USA; Departments of Internal Medicine and Paediatrics, Massachusetts General Hospital, Boston, MA, USA; Department of Immunology and Infectious Diseases, Harvard T.H. Chan School of Public Health, Boston, MA, USA; Center for Communicable Disease Dynamics, Department of Epidemiology, Harvard T.H. Chan School of Public Health, Boston, MA, USA

**Author notes:** Authors contributed equally (Magdalene K. Walters, Michelle A. Bulterys).

## Abstract

**Background:** Eliminating HIV vertical transmission (VT) is a global priority and monitored by estimating paediatric HIV infections with the Joint United Nations Programme on HIV/AIDS supported Spectrum AIDS Impact Module (Spectrum-AIM). Recent innovations in antiretroviral therapy (ART) service delivery models and first-line regimens aimed to reduce VT probabilities. We conducted a systematic review and meta-analysis to estimate VT probabilities by maternal immunologic and treatment status.

**Methods:** We combined an updated systematic review with previous data in meta-regression models to estimate VT probabilities and determinants. We searched multiple databases for peer-reviewed English-language studies from all regions published between January 2018 and February 2024 with VT data stratified by maternal immunologic or treatment status from randomized trials, cohort, or observational studies. Four meta-regression models estimated VT probabilities. We assessed model sensitivity and compared estimates to Spectrum-AIM’s previous results. Finally, we fit a meta-regression model to assess the association of ART class and initiation timing on viral load suppression (VLS) at delivery.

**Findings:** The updated review identified 24 new studies, yielding 110 total studies included in meta-regression analysis. For women not receiving ART, higher CD4 was associated with lower odds of perinatal VT (odds ratio [OR] 0.80 (95% CI: 0.75-0.84) per 100 CD4 cells/*µL* increase). For pregnant women on ART, each additional week on ART before delivery reduced odds of VT by 5.6% (3.2%–7.0%). The odds ratio of perinatal VT among pregnant women initiating integrase inhibitor-based ART 20 weeks pre-delivery was 0.36 (0.14–0.94) compared to those initiating non-nucleoside reverse transcriptase inhibitors (NNRTI)-based ART. This association was confounded by study region. Odds of VLS were lower when ART was initiated late in pregnancy (OR: 0.37 (0.21-0.68) for the reference regimen (NNRTI)), without significant difference by ART regimen.

**Interpretation:** VT probability varies by maternal immunologic stage, treatment regimen, and timing of treatment initiation. These estimates have been incorporated into Spectrum-AIM for UNAIDS 2025 HIV estimates. Earlier ART initiation is associated with higher odds of VLS at delivery. Further evidence is needed on the effects of recent ART innovations on VT outcomes.

**Funding:** NIH, UNAIDS, and UKRI

**RESEARCH IN CONTEXT:** *Evidence before this study:* Three previous systematic reviews have assessed vertical transmission probability to inform assumptions for modelled estimates of perinatal and breastfeeding vertical HIV transmission with the Spectrum AIDS Impact Module (Spectrum- AIM). The most recent systematic review, conducted in 2018, calculated vertical transmission probabilities as the weighted average of literature published before 2018. Since 2018, dolutegravir has been widely adopted as first-line antiretroviral for all people living with HIV, differentiated service delivery models for pregnant and breastfeeding women have aimed to improve care engagement, and immediate lifelong ART initiation following HIV diagnosis has become standard for all people living with HIV. These biomedical and service delivery interventions aim to increase the proportion of pregnant and breastfeeding women who are virally suppressed before delivery, but evidence is limited about their impact on vertical transmission outcomes. To identify any previous systematic review and meta-regression studies addressing vertical transmission rates and impacts of these innovations therein, we searched PubMed from database inception to 4 January 2025 for published articles using the search terms (“vertical transmission” OR “mother-to-child transmission” OR “MTCT” AND “HIV” AND “systematic review” AND “meta-regression” OR “meta-analysis”), resulting in 27 relevant articles. No articles estimated vertical transmission probability stratified by prevention of vertical transmission intervention. Articles that estimated cumulative vertical transmission probabilities were restricted to a country or a region.

*Added value of this study:* This study systematically reviewed literature published before 8 February 2024 on HIV vertical transmission probabilities stratified by maternal clinical or treatment status. It is the first analysis using meta-regression to estimate vertical transmission probabilities that are compatible with Spectrum-AIM. Compared to previous analyses, the meta-regression approach improves accounting for between-study heterogeneity and quantifies statistical uncertainty associated with the vertical transmission probabilities used in Spectrum-AIM. Overall patterns of relative vertical transmission probabilities were similar to previous reviews, but we estimated marginally higher transmission probabilities among breastfeeding women. Initiation of antiretrovirals before the second trimester of pregnancy was associated with higher odds of maternal viral load suppression at delivery. The systematic review identified limited new data about impacts of new first-line drug regimens or service delivery models outside of high-income settings.

*Implications of all the available evidence:* When implemented in Spectrum-AIM, the vertical transmission probabilities estimated here result in more infections during breastfeeding than previous estimates. This underscores the importance of testing HIV-exposed infants for final HIV status outcome after breastfeeding has ended. Additionally, our results show the importance of initiating antiretrovirals as early as possible during or before pregnancy to preserve the woman’s health and achieve viral suppression by delivery to reduce the probability of vertical transmission. Further evidence is needed on the effect of integrase inhibitor-based regimen on vertical transmission in diverse global regions.

## Introduction

Eliminating HIV vertical transmission (VT) and reaching children living with HIV with effective lifelong antiretroviral therapy (ART) are global priorities.^1^ These efforts require accurate data on paediatric HIV infection levels and effectiveness of strategies to prevent vertical transmission (PVT). Due to incomplete HIV testing among pregnant or breastfeeding women, infants, and children, most countries rely on mathematical models to create estimates, namely the Spectrum AIDS Impact Module (Spectrum-AIM) supported by the Joint United Nations Programme on HIV/AIDS (UNAIDS).^2^ Modelled estimates indicate that VT rates have dramatically declined over the past two decades, coinciding with successful PVT programme scale-up.^3^

PVT programmes provide pregnant and breastfeeding women living with HIV (WLHIV), and potentially their infants, with antiretrovirals to suppress viral replication.^4–6^ Strategies recommended by the World Health Organization (WHO) have evolved with therapeutic advancements and scientific evidence.^7–9^ Initial 2003 guidelines recommended short-course antiretrovirals during pregnancy. In 2013 and 2015, WHO recommended immediate lifelong antiretroviral therapy (ART) initiation first for all pregnant WLHIV (referred to as ‘Option B+’) and then for all people living with HIV (PLHIV).^5,6^ To reflect evolving PVT strategies and their efficacies, Spectrum-AIM models final VT rates over time using national data on the number of pregnant WLHIV,^2^ maternal immunologic status, transmission timing (perinatal or breastfeeding), breastfeeding duration among WLHIV,^10^ and VT probability according to PVT strategy.^11^ VT probabilities are stratified by the following groups: WLHIV who did not receive any ARVs, women who seroconverted during pregnancy or breastfeeding and did not receive ARVs, women who received short-course ARVs for PVT (maternal single-dose Nevirapine (sdNVP),^12^ WHO 2006 dual ARV prophylaxis,^13^ Option A,^4^ and Option B^4^), and women on lifelong ART (definitions on Appendix p3-4).

VT probabilities for Spectrum-AIM were initially estimated in a 2012 systematic review^14^ and subsequently updated with new systematic reviews in 2015^15^ and 2018,^16^ as empirical data on newer PVT strategies accumulated. While PVT guidelines have not changed since the 2015 immediate lifelong ART recommendation,^6^ innovations in ART regimens and service delivery models have aimed to improve HIV treatment coverage and effectiveness, including PVT. In 2019, dolutegravir was recommended as first-line ART for all PLHIV, including pregnant WLHIV.^17^ PLHIV on dolutegravir achieve rapid viral load suppression (VLS), which may reduce VT when initiated late in pregnancy compared to previous first-line regimens.^18,19^ Since the 2018 review, Universal Test and Treat and differentiated service delivery have aimed to increase early ART initiation and retention among pregnant and breastfeeding WLHIV.^20^

This study (1) updated previous systematic reviews of VT probabilities with new data since 2018, (2) applied meta-regression to synthesise VT probabilities from pooled data identified in 2012, 2015, 2018, and 2024 systematic reviews, (3) assessed ART regimen class and weeks on ART before delivery as VLS determinants at delivery, and (4) quantified the impact of updated VT probabilities on modelled estimated paediatric infections using Spectrum-AIM.

## Methods

This review was pre-registered on PROSPERO (CRD: <u>42024511011</u>) and reported according to Preferred Reporting Items for Systematic Reviews and Meta-Analyses (PRISMA) guidelines (Appendix p5-8, Tables 2.2.1-2.2.2). Ethical approval was obtained from the Imperial College Research Ethics Committee (Reference: 6300528).

### 2024 systematic review update

We searched PubMed, Embase, Global Health Database, WHO Global Index Medicus, CINAHL Complete, and Cochrane CENTRAL with search term domains mentioning “HIV”, “transmission”, “perinatal” and “breastfeeding periods”, and “infants born to women living with HIV” or related terms. Complete search strategies are in Appendix p9-13.^21^

Database searches included literature published between 1 January 2018 and 8 February 2024. The systematic review was managed using Covidence software.^22^ Title and abstracts were screened for eligibility and full-text articles reviewed for inclusion by two independent reviewers (MKW, MBu, MBa, SH, AR, MS), with conflicts resolved through consensus.

Inclusion criteria were: English-language articles from all geographic regions that reported VT data from randomized trials, cohort, or observational studies. VT data needed to be stratified by maternal PVT regimen or maternal CD4 category for women not receiving PVT; if WLHIV received ART during pregnancy, ART initiation timing (preconception or during pregnancy) was required. Cross-sectional, case-control, case series, case reports, commentaries, letters to editors, study protocols, grey literature, conference abstracts, and non-human animal studies study designs were excluded.

The primary outcome was VT probability, either explicitly reported in studies or calculated using the proportion of infants diagnosed with HIV among exposed infants during pregnancy or breastfeeding. Extracted data included VT timing, infant feeding (breast vs. formula), study characteristics (authors(s), study title, publication year, geography, study years), PVT strategy, maternal ART initiation timing, and clinical data (e.g., viral load or VLS, and CD4 for women not receiving ARVs) (full list in Appendix p14). Data were independently extracted by two reviewers, resolving discrepancies through consensus.

### Meta-regression of HIV vertical transmission probabilities

We fit four meta-regression models to pooled data from the 2024 and previous systematic reviews to estimate VT probabilities stratified by Spectrum-AIM’s VT categories.^15^ The four models estimated VT probability on the logit scale among different groups: 1) WLHIV not receiving ARVs by CD4, 2) WLHIV who seroconverted during pregnancy or breastfeeding or received short-course ARVs for PVT, 3) WLHIV exposed to perinatal transmission while receiving lifelong ART by initiation timing, and, 4) WLHIV exposed to breastfeeding transmission receiving lifelong ART. We also assessed ART regimen class as a predictor for VT probability and whether VLS at delivery (<50 copies/mL) was associated with ART regimen or initiation timing.

Perinatal and breastfeeding VT definitions remained the same as previous reviews.^14^ Perinatal VT was defined as HIV acquisition occurring before six weeks postpartum. For studies reporting transmission during breastfeeding, we converted cumulative acquisition probabilities to monthly probability for the period starting at the end of the perinatal period (six weeks) and ending at the time of the last HIV test closest to breastfeeding cessation. Most often this period spanned six weeks to six months (see Appendix p15).

Model one estimated VT among WLHIV not receiving ARVs as a function of the study population’s baseline CD4 midpoint:

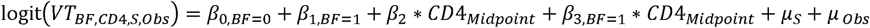

Model one included fixed effects for perinatal (*β*_0,*BF*=0_) and monthly breastfeeding VT (*β*_1,*BF*=1_) intercepts, study population CD4 midpoint (*β*_2_, per 100 cells/*µL,* centred at 500 cells/*µL*), and an interaction between CD4 midpoint and breastfeeding timing (*β*_3,*BF*=1_). CD4 midpoint was extracted as the median CD4 of WLHIV not receiving ARVs or the midpoint of relevant CD4 range if studies reported VT by CD4 categories. More information on CD4 midpoint determination from each study is in Appendix p27-30. Random effects were included for study and observation (*µ_S_* and *µ_Obs_*, respectively).

Model two estimated VT probability among WLHIV who acquired HIV infection during pregnancy or breastfeeding (maternal seroconversions) or received short-course PVT.

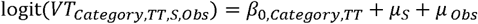

Model two included fixed effects for categories (*β*_0,*Category,TT*_) used in Spectrum-AIM, all stratified by transmission timing (*TT)*: maternal seroconversion, WLHIV receiving WHO 2006 dual ARV regimen, sdNVP, Option A, and Option B. For breastfeeding women receiving sdNVP, transmission rates were stratified by CD4 <350 cells/*µ*L and CD4 ≥350 cells/*µ*L. Random effects were included for study and observation (*µ_S_* and *µ*_*Obs*_, respectively). Model two categories are defined by the common feature that they represented categorical exposures for VT, but did not involve adjustment for continuous covariates, such as median CD4 count (model one) or lifelong ART initiation timing (models three and four) (see Appendix p3-4 for definition details).

Model three estimated perinatal transmission probabilities among women on lifelong ART by maternal ART initiation timing:

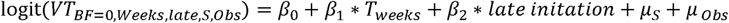

Model three included a fixed intercept (*β*_0_) representing VT when ART is initiated 20 weeks before delivery, slope (*β*_1_) for weeks on ART before delivery (*T_Weeks_*, centred on ART initiated 20 weeks before delivery; specified as 40 weeks for women already on ART at conception), and a fixed effect for ART initiation within four weeks of delivery (*β*_2_, ‘late initiation’). The ‘late initiation’ effect accounted for increased risk of unsuppressed VL at delivery when ART is initiated late in pregnancy. *T_Weeks_* was preferentially extracted as the reported median weeks on ART before delivery, or, otherwise, taken as the range midpoint for studies that reported

ART initiation as a range of gestational weeks. We contacted corresponding authors of 26 studies that reported ART initiation timing as a range to request the median ART initiation gestational week. Assumptions to estimate weeks on ART are described in Appendix p31-36. Random effects were included for study and observation (*µ_S_* and *µ_Obs_*, respectively).

Model four estimated monthly breastfeeding transmission probabilities by ART initiation timing (preconception or during pregnancy):

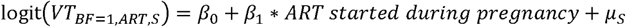

Model four included two fixed effects for ART started preconception (*β*_0_) and for ART started during pregnancy (*β*_1_). For breastfeeding transmission, ART initiation timing was classified as a binary covariate rather than continuous weeks before delivery (as in Model 3 for perinatal transmission) because time on ART during pregnancy is less directly related to VLS during breastfeeding than VLS at delivery. This model included random effects by observation (*µ_S_*). Study level random effects were not included because only two studies had multiple observations.

To assess associations between ART regimen class on perinatal transmission probability, we modified model three to include fixed effects for ART regimen class (non-nucleoside reverse transcriptase inhibitors [NNRTI] (reference), integrase inhibitors [INSTI], protease inhibitors [PI], and miscellaneous). We evaluated geographic region as a confounder of this association (sensitivity analysis, Appendix p36).

Regarding determinants of VLS at delivery, we analysed studies that reported the proportion of WLHIV with VLS (<50 c/mL) at delivery, time on ART, and ART regimen class:

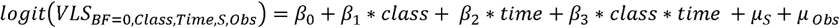

This model included a fixed effect intercept (*β*_0_) for transmission among women initiating NNRTI early (‘early’: before second trimester start) fixed effects for ART regimen class relative to NNRTI for early initiation (*β*_1_), timing of ART initiation (‘late’: after the first trimester and before delivery, relative to early), and an interaction between ART class and timing.

Study and observation random effects were included (*µ_S_* and *µ_Obs_*, respectively).

### Implications of estimated VT probabilities for Spectrum-AIM estimates of paediatric HIV infections

We used predicted values from models 1-4 to produce VT probability estimates compatible with stratifications in Spectrum-AIM. For VT probabilities among WLHIV not receiving PVT stratified by CD4 categories <200, 200-349, and ≥350, we used model one to predict VT probabilities corresponding to CD4 midpoint values 100, 275, and 500, respectively. Model two was used to predict VT probabilities for maternal seroconversion and short-course PVT. For perinatal VT probabilities among women on ART <4 weeks, 4-39 weeks, and pre- conception, model three was used to predict probabilities corresponding to 2, 20, and 40 weeks on ART, respectively. Model four was used to predict breastfeeding VT probabilities for women initiating ART before conception or during pregnancy.

Simulations of impacts on paediatric infections were conducted in an R implementation of the Spectrum-AIM paediatric model that matched Spectrum-AIM v6.37 (https://doi.org/10.5281/zenodo.15166687) in four countries (Rwanda, Malawi, Democratic Republic of Congo (DRC), and Burkina Faso) in 2000, 2010, 2015, and 2023. Spectrum- AIM files were used for UNAIDS 2024 Global HIV Estimates. Results were compared to the 2024 UNAIDS published paediatric HIV infections, using VT probabilities used in Spectrum- AIM for UNAIDS estimates published 2019-2024 (Appendix p3-4; henceforth referred to as the “former” VT probabilities).^3^

Analyses were conducted in R 4.3.1.^23^ Mixed-effect meta-regression models were fit using the R package ‘glmmTMB’ version 1.1.9.^24^ Confidence intervals were calculated as the 2.5th and 97.5th percentiles of 3000 Monte Carlo samples from each model’s joint posterior distribution. Data and code for model analyses are available from https://github.com/mwalte10/hiv_vt_mr.

## Results

Our updated search identified 12,588 records. After deduplication, 6,730 unique titles and abstracts were screened (Figure 1). Full texts were reviewed for 424 studies, 400/424 were excluded, primarily due to only reporting VT aggregated across distinct PVT groups (172/400). Data were extracted from 24 new studies published since 1 January 2018 and combined with data from 30, 36, and 20 studies from the 2012, 2015, and 2018 reviews, respectively (Figure 1), yielding 110 studies included in meta-regression analyses.

**Figure 1.**
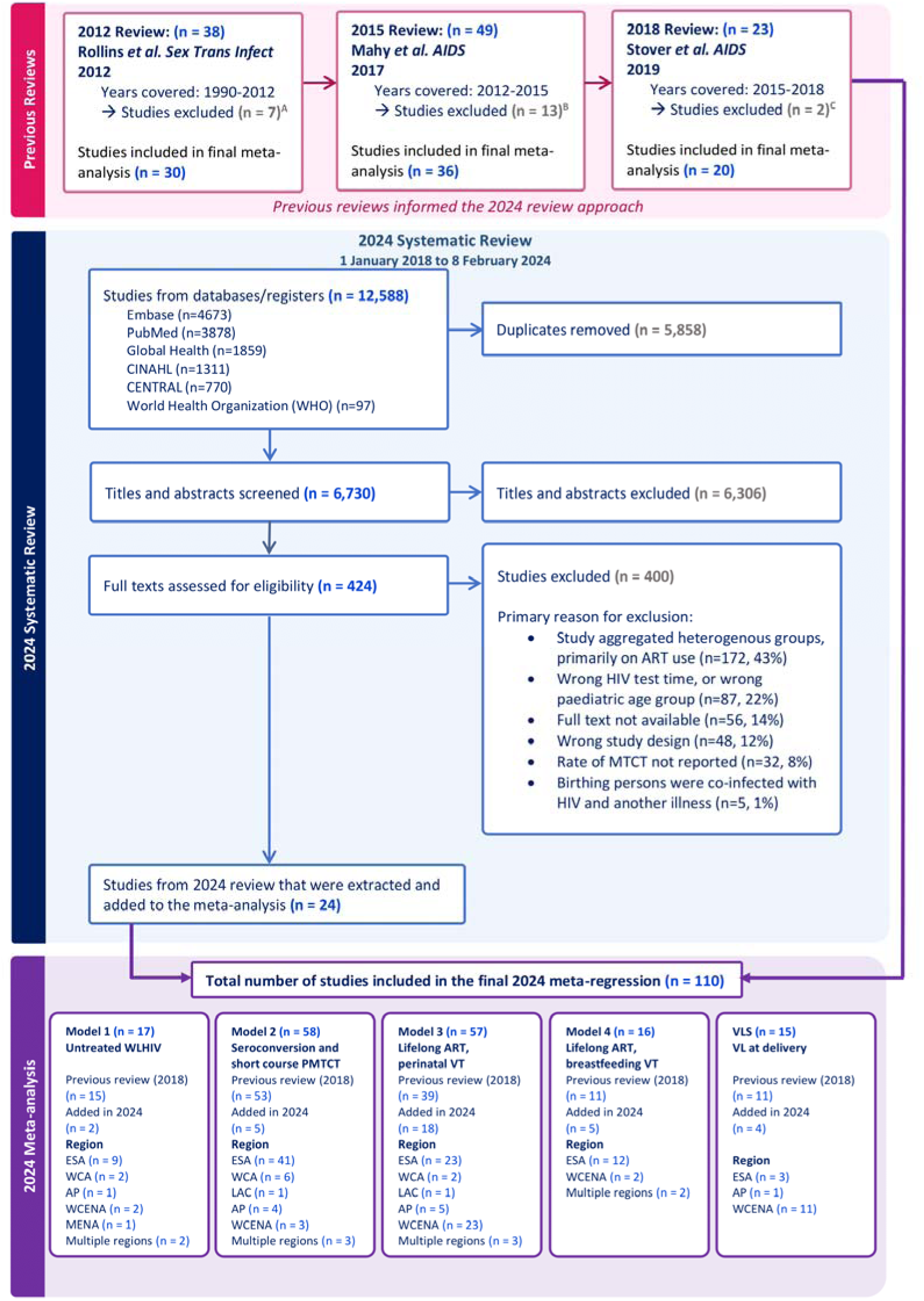
PRISMA Flow Diagram for 2018-2024 review. Studies were excluded from previous reviews for the following reasons: A Not peer-reviewed (3), duplicate data (3), aggregates heterogenous results (1) B Not peer-reviewed (5), duplicate data (4), aggregates heterogeneous results (4) C Not peer-reviewed (2)

Publications occurred between 1988-2023 on data collected between 1982-2022. All global regions were represented, but most studies were conducted in Eastern and Southern Africa (56/110, Appendix p48-56).

Model one included data from 17 studies on VT among women not receiving PVT by CD4 category, 2/17 identified in the 2024 systematic review (Figure 1; forest plots: Appendix p19- 20, Figures 3.2.1 and 3.2.2). 80.2% (9642/12,021) of observations were from studies that stratified VT by CD4 range. Each additional 100 cells/*µ*L in median CD4 was associated with 20% lower VT odds (95% CI: 0.75-0.84) (Appendix p16 Table 3.1.1). The interaction between CD4 midpoint and breastfeeding transmission was not statistically significant (p16 Table 3.1.1). Predicted perinatal transmission probabilities for women with CD4 <200, 200- 350, and >350 were, respectively, 33.4% (27.8–39.1%), 25.1% (21.4–28.9%), and 16.6% (13.9–19.7%) (Table 1). Monthly breastfeeding transmission probabilities were 1.02% (0.32–3.32%), 0.90% (0.49–1.73%), and 0.79% (0.43–1.43%), respectively (Table 2). In sensitivity analyses, fitting the model to only studies that reported study population median CD4 (not CD4 ranges changed the estimated relationship between CD4 and VT (Appendix p29 Table 4.1.1.2). The restricted criteria made the model disproportionately sensitive to observations from three studies published between 1991-1998 reporting very high transmission rates as 80.2% of all observations were omitted (Appendix p27-30).

**Table 1.**
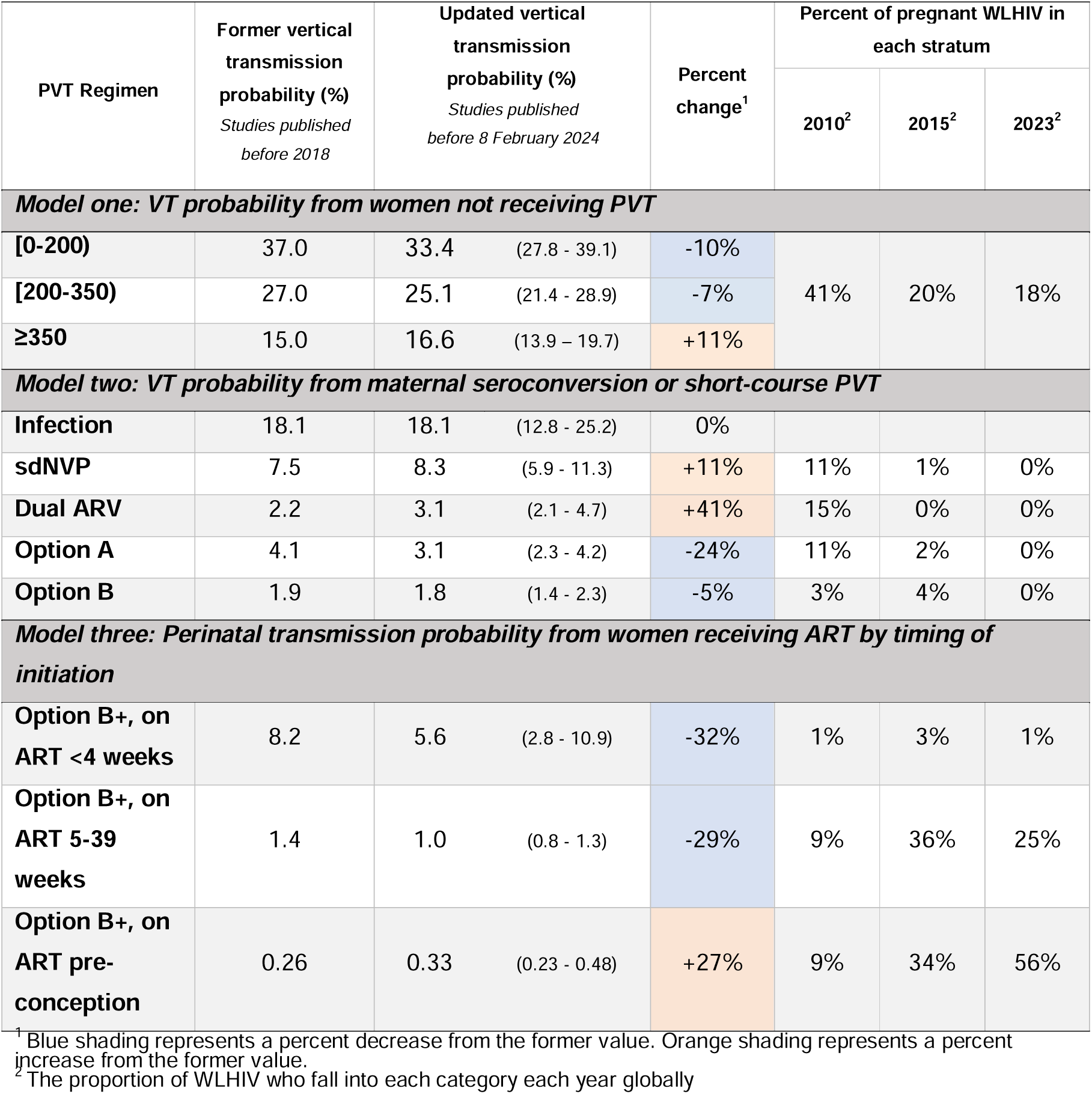
Perinatal vertical transmission probabilities.

**Table 2.** Breastfeeding vertical transmission probabilities. All breastfeeding vertical transmission probabilities are per month, except for maternal seroconversions (“Infection” below) which is a one-time probability for breastfeeding WLHIV.

| PVT Regimen | Former vertical transmission probability (%)<br>*Studies published before 2018* | Updated vertical transmission probability (%)<br>*Studies published before 8 February 2024* |  | Percent change<sup>1</sup> | Percent of pregnant WLHIV in each stratum |  |  |
| --- | --- | --- | --- | --- | --- | --- | --- |
|  |  |  |  |  | 2010<sup>2</sup> | 2015<sup>2</sup> | 2023<sup>2</sup> |
| Model one: VT probability from women not receiving PVT |  |  |  |  |  |  |  |
| [0-200) | 0.89 | 1.02 | (0.32 - 3.32) | +15% | 41% | 20% | 18% |
| [200-350) | 0.81 | 0.90 | (0.49 - 1.73) | +11% |  |  |  |
| ≥350 | 0.51 | 0.79 | (0.43 - 1.43) | +55% |  |  |  |
| Model two: VT probability from maternal seroconversion or short-course PVT |  |  |  |  |  |  |  |
| Infection (one-time, not monthly) | 26.9 | 28.29 | (20.82 – 37.20) | +5% |  |  |  |
| sdNVP, <350 | 0.99 | 0.89 | (0.16 - 4.45) | -10% | 11% | 1% | 0% |
| sdNVP, ≥350 | 0.40 | 0.35 | (0.06 - 1.94) | -13% |  |  |  |
| Dual ARV | 0.18 | 0.20 | (0.06 - 0.74) | +11% | 15% | 0% | 0% |
| Option A | 0.20 | 0.21 | (0.06 - 0.63) | +5% | 11% | 2% | 0% |
| Option B | 0.13 | 0.14 | (0.07 - 0.30) | +8% | 3% | 4% | 0% |
| Model four: Monthly breastfeeding transmission from women receiving lifelong ART |  |  |  |  |  |  |  |
| Option B+, on ART <4 weeks | 0.20 | 0.13 | (0.08 - 0.22) | -35% | 1% | 3% | 1% |
| Option B+, on ART 5-39 weeks | 0.11 |  |  | +18% | 9% | 36% | 25% |
| Option B+, on ART pre-conception | 0.02 | 0.02 | (0.00 - 0.06) | 0% | 9% | 34% | 56% |

<sup>1</sup> Blue shading represents a percent decrease from the former value. Orange shading represents a percent increase from the former value. <sup>2</sup> The proportion of WLHIV who fall into each category each year globally

Model two included data from 58 studies on VT among women who seroconverted or received short-course PVT, 4/58 identified in the 2024 review (Figure 1; forest plots: Appendix p21-24 Figures 3.2.3–3.2.13). Among women who seroconverted during pregnancy, 18.1% (12.8–25.2%) transmitted HIV perinatally (Table 1). Among women who seroconverted during breastfeeding, 28.29% (20.82–37.20%) transmitted HIV infection (Table 2). Among women who received short-course PVT regimens, perinatal transmission probabilities were 8.3% (5.9–11.3%) for sdNVP, 3.1% (2.1–4.7%) for dual ARV, 3.1% (2.3– 4.2%) for Option A, and 1.8% (1.4–2.3%) for Option B (Table 1). Monthly breastfeeding transmission probabilities were 0.89% (0.16–4.45%) and 0.35% (0.06–1.94%) for sdNVP among women with CD4 <350 and ≥350, respectively, 0.20% (0.06–0.74%) for dual ARV, 0.21% (0.06–0.63%) for Option A, and 0.14% (0.07–0.30%) for Option B.

Model three included data from 57 studies on perinatal transmission among women on lifelong ART, 15/57 identified in the 2024 systematic review (Figure 1 and Figure 2; forest plots: Appendix p25 Figure 3.2.14). Five of 26 studies provided additional unpublished data on median gestational week at ART initiation. Each additional week on ART before delivery reduced VT odds by 5.6% (4.2%–7.0%, Appendix p16 Table 3.1.1). Perinatal VT probability was 5.6% (2.8–10.9%) among women who initiated ART late (four weeks before delivery), 1.0% (0.8–1.3%) among women who initiated ART 20 weeks before delivery, and 0.33% (0.23–0.48%) among women who initiated ART preconception (Table 1). When model three was restricted to only studies that reported the median weeks on ART (rather than a range of weeks when ART was initiated) the odds ratio for less than four weeks compared to 20 weeks was 10.1 (6.6–16.1, Appendix p31-36).

**Figure 2.**
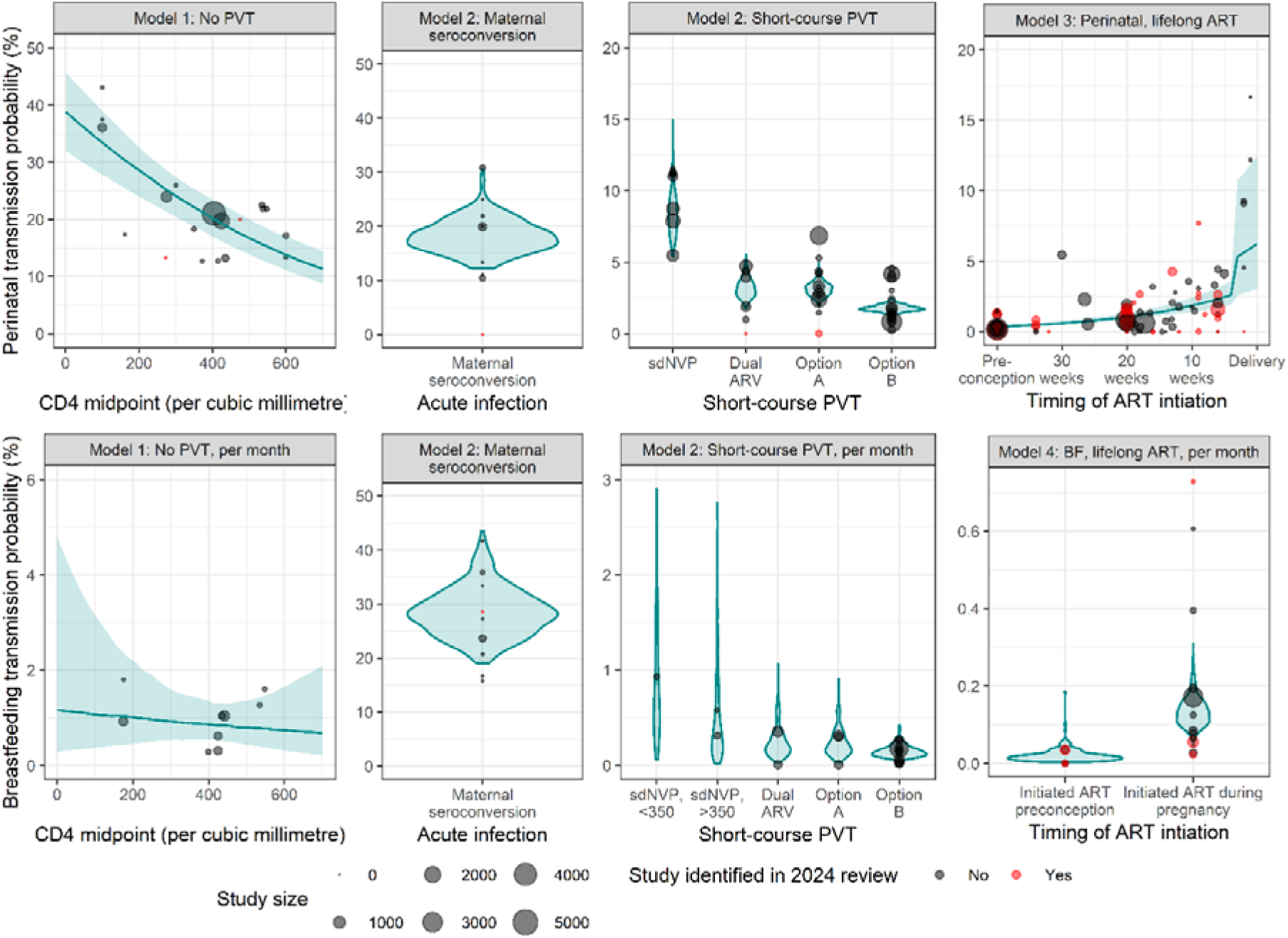
Data used in models one through four and model estimates of VT probability. Point size reflects study size and colour indicates whether the study was identified in the 2024 systematic review. Blue violin plots are used for categorical models (model two and model four) and blue lines with ribbon are used for continuous models (model one and model three). Panels represent the different models used to produce estimates of VT probability compatible with Spectrum-AIM, with the top row displaying perinatal VT probabilities and the bottom row displaying breastfeeding VT probabilities.

Model four included data from 16 studies on breastfeeding transmission among women on lifelong ART; 5/16 identified in the 2024 systematic review (Figure 1, Appendix p26 Figure 3.2.15). The monthly breastfeeding transmission probability was 0.13% (0.08–0.22%) for women who initiated ART during pregnancy and 0.02% (0.00–0.06%) for women who initiated ART preconception (Table 3).

When refitting model three with fixed effects for ART regimen class, WLHIV who initiated an INSTI- versus NNRTI-based regimen had significantly lower VT odds (aOR: 0.36; 0.14–0.94, Appendix p17). After adding geographic region to the model, associations with ART regimen were no longer statistically significant (INSTI vs. NNRTI aOR: 0.48; 0.17–1.32; Appendix p36).

Fifteen studies reported data on the proportion of WLHIV with VL <50 copies/mL at delivery; three were identified in the 2024 systematic review (Figure 1). Most (11/15) studies were from Western and central Europe and North America. VLS probability at delivery was highest among WLHIV who initiated INSTI-based regimens before the second trimester (95.8% (66.8–99.6%), Appendix p17); however, the difference between regimen classes was not statistically significant. VLS probability among women who started ART before the second trimester was 90.6% (80.4–95.4%) for NNRTI, 79.2% (42.3–95.0%) for PI, and 90.0% (62.9–97.8%) for miscellaneous regimens. VLS probability among women who initiated ART after the first trimester was 40.4% (3.3–90.2%) for INSTI, 82.8% (70.2–90.4%) for NNRTI, 65.6% (51.7–76.5%) for PI, and 74.3% (2.8–99.6%) for miscellaneous regimens.

Tables 1 and 2 compare the updated VT probabilities to the former default VT probabilities in Spectrum-AIM. The mean percent difference was 2%. The percent difference was largest for monthly breastfeeding transmission probability among women with CD4 >350 (55% increase from 0.51%/month to 0.79%/month; Table 2). Updated perinatal transmission probabilities were on average lower (mean percent difference: 1.5% lower), whereas updated breastfeeding transmission probabilities were on average higher (mean percent difference: 5.8% higher).

We incorporated the updated VT probabilities (Tables 1 and 2) into Spectrum-AIM to estimate the number of paediatric infections in Rwanda, Malawi, DRC, and Burkina Faso (Appendix p38-46). In 2023, total paediatric infections (perinatal and breastfeeding) from the updated VT probabilities were 8.8% higher than those estimated from the former VT probabilities (Figure 3). Perinatal infections were slightly lower except in DRC (average 2.5% lower) and breastfeeding infections were higher in all countries (average 15.6% higher; Figure 3). Increases in estimated VT infections during breastfeeding were largest among women who did not receive PVT or experienced treatment interruptions (Table 2).

**Figure 3.**
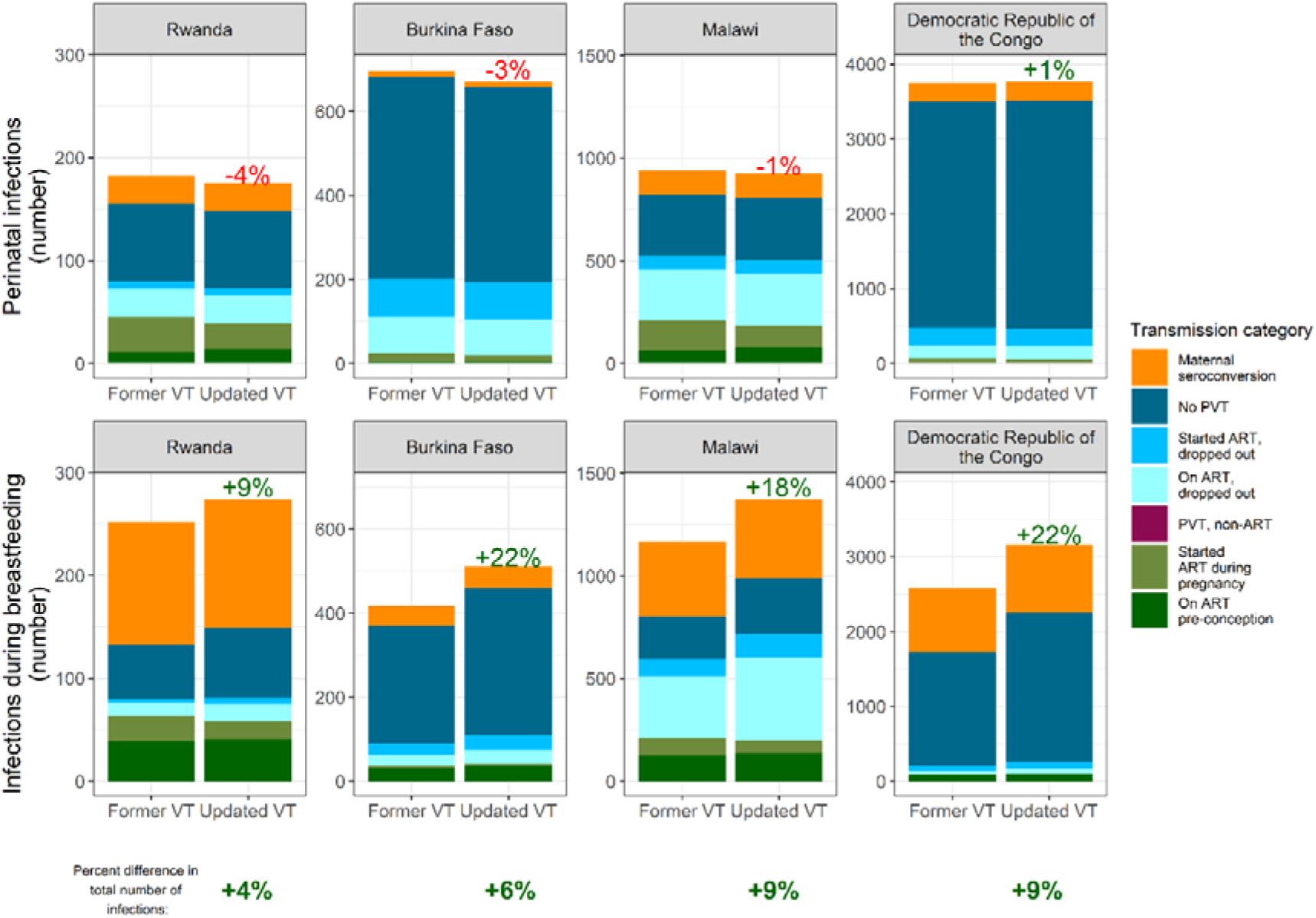
Differences in 2023 number of paediatric infections using the Spectrum-AIM former vs. updated VT probabilities. Coloured percentage points represent the percent difference between the infections using the former versus updated VT probabilities. Green and red percentages denote the relative percent increase and decrease in the number of infections between former and updated VT probabilities.

## Discussion

We systematically updated evidence about HIV VT determinants and probabilities, including the effectiveness of ARV-based strategies to prevent transmission. This information is critical to produce modelled estimates of paediatric HIV infections and guide strategies to eliminate VT. Since the 2018 review, 24 studies were published with data relevant to inform parameters in Spectrum-AIM. Compared to previous assessments, we applied a more systematic meta-regression approach that strengthened pooling of information and quantified statistical uncertainty for the first time.

Patterns of VT probabilities were consistent with previous reviews.^14–16^ VT was highest among women not receiving PVT, lower among women receiving short-course PVT regimens, and lowest among women initiating ART during and before pregnancy. Changes from former default values^2^ are attributable to both new data since the previous review and the meta-regression analytical approach. Perinatal VT probabilities are slightly lower than reported in previous reviews.^16^ Of perinatal VT probabilities, VT probability among women initiating ART within four weeks was the largest relative decrease (8.2% to 5.6%); only 1% of pregnant WLHIV globally initiate ART within four weeks of delivery, so this change will not substantially change estimates of perinatal infections. Updated breastfeeding VT probabilities were higher than previous values. The largest change was among women who did not receive PVT, for which assumed probabilities had not been updated since the 2012 review.

The VT probabilities derived here have been adopted as default values recommended for countries using Spectrum-AIM to update national HIV estimates in 2025. These new values are unlikely to substantially change the number of perinatal infections but will increase estimated infections during breastfeeding. The shift in infection timing to the breastfeeding period has implications for HIV testing strategies. Retaining mother-infant pairs in PVT services until breastfeeding cessation is important to prevent transmission and monitor HIV outcomes. Additionally, universal final HIV outcome ascertainment could be used to compare programmatic data to estimates of new infections from Spectrum-AIM.

We evaluated emerging evidence on scale-up of Universal Test-and-Treat, differentiated service delivery models, and dolutegravir-based first-line regimens at increasing VLS and reducing VT among pregnant and breastfeeding WLHIV.^25^ We found limited data to assess these. ART initiation before the second trimester was associated with higher VLS at delivery, consistent with observed lower transmission probability among women who initiated ART earlier. However, data on VLS at delivery was predominantly limited to studies conducted in high-income countries and where women initiated INSTI-based regimens early in pregnancy.

Regarding differences by ART regimen, women who initiated INSTI-based regimens (including dolutegravir) had the lowest perinatal VT probability. Data were insufficient to assess the impact of ARV drug class on VT probability during breastfeeding. These results should be interpreted cautiously as studies with INSTI-based regimens were predominately in high-income countries; only one study reported VT probability among women receiving INSTI-based regimens was conducted in Africa. The difference in VT probability by regimen was attenuated and not statistically significant when adjusting for geographic region.

Moreover, data indicating different VT rates associated with INSTI-based regimens were predominately among women initiating ART before or early in pregnancy, while cohort studies comparing regimens among the same study populations have found no regimen differences in VT and VLS among women when ART is initiated early,^26^ but with potential benefits when ART is initiated late due to faster VLS associated with INSTI-based regimens. As such, we believe additional data among women initiating INSTI late in pregnancy are needed before recommending modelling VT probability by ART regimen.

Our systematic review has limitations. Restricting to studies that disaggregated VT by PVT strategy and ART initiation timing excluded 43% of full texts that reported unstratified VT. Individual-level information about maternal CD4 or ART initiation timing and VT outcomes may have quantified these relationships more precisely than the broad categories from our systematic review. The association between CD4 count and VT probability was significantly different in studies that reported CD4 range versus CD4 midpoint, driven by three studies conducted in the 1990s that reported CD4 range and very high VT that accounted for only 6% of model one’s observations. Regarding ART initiation timing, the relationship between weeks on ART and VT did not differ between reporting median weeks versus range of weeks. Only including studies that reported median weeks would have excluded 76% of all studies that reported ART initiation occurring during the third trimester. We did not estimate VT probability among women who initiated ART after seroconverting during pregnancy or postpartum, an area for future work as identifying infections during these periods is an increasing focus of PVT programmes through re-testing at late ANC visits, labour and delivery, and postnatally.

In conclusion, vertical HIV transmission probability varies dramatically according to maternal clinical characteristics and ART initiation timing. Our analysis incorporated more data through updated systematic literature search and refined formal statistical synthesis. We found somewhat higher transmission rates during breastfeeding than previous estimates.

Findings reinforce the importance of initiating WLHIV on ART early in pregnancy to progress toward HIV VT elimination. However, data were limited to quantify impacts of recent innovations in ART regimens and service delivery models on VT rates; this should continue to be assessed as new data emerges.

## Data Availability

All data produced in the present study are available upon reasonable request to the authors.

https://github.com/mwalte10/hiv_vt_mr/tree/main/data

## Data sharing

Data and a data codebook are publicly available at https://github.com/mwalte10/hiv_vt_mr/tree/main/data.

## Contributions

MKW, MAB, KP, and JWI-E conceptualized the study as a systematic review and meta-regression. MAB supervised the systematic review methodology and MKW supervised the data extraction and meta-regression. MAB and DL developed the methodology for the search strategy. MKW, MAB, MB, SH, AR, and MS screened abstracts and full texts for inclusion and extracted data. MKW, JWI-E, and M-CB developed the meta-regression methodology and HK, KP, LM, MM, JS, and RG provided feedback on study design and analytic approaches. MKW conducted the analysis and drafted the initial manuscript with assistance from MAB. Figures were created by MKW, MAB, and MB. All authors revised the manuscript for intellectual content and approved the final manuscript.

## Declaration of interests

SH declares grants from NICHD F31HD116617 for work outside the submitted work. MCB declares funding from the NIH to HPTN and Wellcome Trust paid to her institution. MM is an employee of UNAIDS and received financial support through her institution. LM declares funding from ViiV to her employer (Elizabeth Glaser Pediatric AIDS Foundation) as well as paid consultancy fees from the WHO. KP declares funding from the National Institute of Health, Harvard T.H. Chan School of Public Health, and John Hopkins University paid to her institution. KP declares involvement on a data safety monitory board for Bifidobacterium infantis for Infant Immune Development, an RCT evaluating use of probiotics in infants (outside of submitted work). JWI-E declares grants from UNAIDS and Gates Foundation, consulting fees from BAO systems, and meeting travel support from UNAIDS, Gates Foundation, and International AIDS Society outside the submitted work.

A preliminary analysis of this work was presented in October 2024 to a virtual meeting of the UNAIDS Reference Group on HIV Estimates, Modelling, and Projections.

## Acknowledgements

This research was supported by the National Institute of Allergy and Infectious Diseases of the National Institutes of Health under award number 1R01AI152721-01A1 (MKW, MB, HK, and JWI-E), UNAIDS (MKW, MM, and JWI-E), and the MRC Centre for Global Infectious Disease Analysis (reference MR/R015600/1, MKW, MCB, JWI-E), jointly funded by the UK Medical Research Council (MRC) and the UK Foreign, Commonwealth & Development Office (FCDO), under the MRC/FCDO Concordat agreement and is also part of the EDCTP2 programme supported by the European Union.

The authors acknowledge and thank participants in the UNAIDS Reference Group on HIV Estimates, Modelling, and Projections for suggestions to refine the analysis, Caitlin Dugdale and Andrea Ciaranello for insights on the viral load analysis, and Edmond Brewer and Megan Verma for assistance with initial title abstract screening.

For the purpose of open access, the author has applied a ‘Creative Commons Attribution’ (CC BY) licence to any Author Accepted Manuscript version arising. The content is solely the responsibility of the authors and does not necessarily represent the official views of the funders.

