## Appendix for "Probability of vertical HIV transmission: A systematic review and meta-regression"

*Authors contributed equally

### 1. Definitions, VT probabilities used from 2019-2014, and source of vertical transmission probabilities used in Spectrum-AIM

Table 1.1 describes the definition of vertical transmission categories in the Spectrum-AIM model^1^ and reports the VT probabilities used in model versions for UNAIDS global HIV estimates published 2019 through 2024 and their sources. Most VT probabilities used between 2019-2024 values are weighted averages of studies identified in the 2018 review by Mofenson.^2^ Estimates of perinatal transmission probability for women who did not receive PVT have not been updated since the 2012 Rollins *et al.* systematic review.^3^

**Table 1.1.** Default perinatal vertical HIV transmission probabilities in Spectrum-AIM model estimates published by UNAIDS from 2019-2024

| Transmission category | Definition | VT probability | Source of VT probability |
| --- | --- | --- | --- |
| CD4 < 200 | Existing infection, mother did not use PVT and had a CD4 < 200. | 0.37 | Median of studies from Rollins *et al.* 2012 systematic review |
| CD4 200-350 | Existing infection, mother did not use PVT and had a CD4 200-350. | 0.27 | Median of studies from Rollins *et al.* 2012 systematic review |
| CD4 ≥ 350 | Existing infection, mother did not use PVT and had a CD4 > 350. | 0.15 | Median of studies from Rollins *et al.* 2012 systematic review |
| Maternal seroconversion | Mother seroconverted during pregnancy and did not receive ARVs. | 0.181 | Weighted average from Mofenson 2018 systematic review update |
| Maternal single dose nevirapine | Mother received only single dose nevirapine as part of PVT. | 0.075 | Weighted average from Mofenson 2018 systematic review update |
| WHO 2006 Dual ARV regimen | Mother utilized two ARV regimens for PVT. | 0.022 | Weighted average of studies with women with CD4 > 350 from Mofenson 2018 systematic review update |
| Option A | Mothers with CD4 > 350 used AZT from week 14 of gestation and single dose nevirapine at the onset of labor. Daily AZT/ 3TC used through 7 days postpartum. | 0.041 | Weighted average from Mofenson 2015 systematic review update |
| Option B | Mothers with CD4 > 350 used triple ARVs starting at week 14 of gestation and continued through breastfeeding cessation. | 0.019 | Weighted average of studies with breastfeeding populations from Mofenson 2018 systematic review update |
| Mother on ART <4 weeks before delivery | Triple ARVs were initiated <4 weeks before delivery and continued for life. | 0.082 | Weighted average from Mofenson 2018 systematic review update |
| Mother on ART >4 weeks before delivery | Triple ARVs were initiated >4 weeks before delivery (but after conception) and continued for life. | 0.014 | Weighted average from Mofenson 2018 systematic review update |
| Mother on ART preconception | Mother was on triple ARVs at conception and continued for life. | 0.0026 | Weighted average from Mofenson 2018 systematic review update |

**Table 1.2.** Default vertical transmission probabilities among breastfeeding women in Spectrum-AIM model estimates published by UNAIDS from 2019-2024

| Transmission category | Definition | VT probability | Source of VT probability |
| --- | --- | --- | --- |
| CD4 < 200  (monthly) | Existing infection, mother did not use PVT and had a CD4 < 200. | 0.0089 | Weighted average from Mofenson 2018 systematic review update of studies without a CD4 restriction |
| CD4 200-350 (monthly) | Existing infection, mother did not use PVT and had a CD4 200-350. | 0.0081 | It is unclear where this VT probability came from, this was not explicitly estimated in the 2012, 2015, or 2018 reviews. |
| CD4 ≥ 350 (monthly) | Existing infection, mother did not use PVT and had a CD4 > 350. | 0.0051 | Rollins *et al.* 2012 systematic review |
| Maternal seroconversion | Mother seroconverted during pregnancy and did not receive ARVs. | 0.269 | Weighted average from Mofenson 2018 systematic review update |
| Maternal single dose nevirapine  CD4 < 350  (monthly) | Mother received only single dose nevirapine as part of PVT. | 0.0099 | Weighted average from Mofenson 2018 systematic review update |
| Maternal single dose nevirapine  CD4 ≥ 350  (monthly) | Mother received only single dose nevirapine as part of PVT. | 0.004 | Weighted average from Mofenson 2018 systematic review update |
| WHO 2006 Dual ARV regimen  (monthly) | Mother utilized two ARV regimens for PVT. | 0.0018 | Weighted average of studies that reported extended infant prophylaxis from Mofenson 2018 systematic review update |
| Option A  (monthly) | Mothers with CD4 > 350 used AZT from week 14 of gestation and single dose nevirapine at the onset of labor. Daily AZT/ 3TC used through 7 days postpartum. | 0.002 | Median of studies in the Rollins *et al.* 2012 systematic review. |
| Option B  (monthly) | Mothers with CD4 > 350 used triple ARVs starting at week 14 and continued through breastfeeding cessation. | 0.0013 | It is unclear where this VT probability came from, the Mofenson 2018 systematic review had a weighted average of 0.0011. |
| On ART <4 weeks before delivery  (monthly) | Triple ARVs were initiated <4 weeks before delivery and continued for life. | 0.002 | Expert opinion from Rollins *et al.* 2012 systematic review |
| On ART >4 weeks before delivery  (monthly) | Triple ARVs were initiated >4 weeks before delivery (but after conception) and continued for life. | 0.0011 | Weighted average of studies where mother started ART at any time during pregnancy from Mofenson 2018 systematic review update |
| On ART preconception  (monthly) | Mother was on triple ARVs at conception and continued for life. | 0.0002 | Weighted average of studies from Mofenson 2018 systematic review update |

### 2. 2024 systematic review

#### 2.2. PRISMA checklist

**Table 2.2.1.** PRISMA checklist

| Topic | No. | Summary | Location |
| --- | --- | --- | --- |
| **Title** | 1 | Identify the report as a systematic review. | Title |
| **Abstract** | 2 | See the PRISMA 2020 for Abstracts checklist | Included in Table 2.2.2 |
| **INTRODUCTION** | | | |
| **Rationale** | 3 | Describe the rationale for the review in the context of existing knowledge. | Introduction |
| **Objectives** | 4 | Provide an explicit statement of the objective(s) or question(s) the review addresses. | Introduction |
| **METHODS** | | | |
| **Eligibility criteria** | 5 | Specify the inclusion and exclusion criteria for the review and how studies were grouped for the syntheses. | Methods |
| **Information sources** | 6 | Specify all databases, registers, websites, organisations, reference lists and other sources searched or consulted to identify studies. Specify the date when each source was last searched or consulted. | Methods |
| **Search strategy** | 7 | Present the full search strategies for all databases, registers and websites, including any filters and limits used. | Methods and Appendix 2.3 |
| **Selection process** | 8 | Specify the methods used to decide whether a study met the inclusion criteria of the review, including how many reviewers screened each record and each report retrieved, whether they worked independently, and if applicable, details of automation tools used in the process. | Methods and Figure 1 |
| **Data collection process** | 9 | Specify the methods used to collect data from reports, including how many reviewers collected data from each report, whether they worked independently, any processes for obtaining or confirming data from study investigators, and if applicable, details of automation tools used in the process. | Methods |
| **Data items** | 10a | List and define all outcomes for which data were sought. Specify whether all results that were compatible with each outcome domain in each study were sought (e.g. for all measures, time points, analyses), and if not, the methods used to decide which results to collect. | Methods and Appendix 2.4 |
|  | 10b | List and define all other variables for which data were sought (e.g. participant and intervention characteristics, funding sources). Describe any assumptions made about any missing or unclear information. | Methods and Appendix 2.4 |
| **Study risk of bias assessment** | 11 | Specify the methods used to assess risk of bias in the included studies, including details of the tool(s) used, how many reviewers assessed each study and whether they worked independently, and if applicable, details of automation tools used in the process. | Not applicable |
| **Effect measures** | 12 | Specify for each outcome the effect measure(s) (e.g. risk ratio, mean difference) used in the synthesis or presentation of results. | Methods and Appendix 2.5 and 2.6 |
| **Synthesis methods** | 13a | Describe the processes used to decide which studies were eligible for each synthesis (e.g. tabulating the study intervention characteristics and comparing against the planned groups for each synthesis (item 5)). | Methods |
|  | 13b | Describe any methods required to prepare the data for presentation or synthesis, such as handling of missing summary statistics, or data conversions. | Methods and Appendix 4 |
|  | 13c | Describe any methods used to tabulate or visually display results of individual studies and syntheses. | Methods and Appendix 4.2 |
|  | 13d | Describe any methods used to synthesize results and provide a rationale for the choice(s). If meta-analysis was performed, describe the model(s), method(s) to identify the presence and extent of statistical heterogeneity, and software package(s) used. | Methods |
|  | 13e | Describe any methods used to explore possible causes of heterogeneity among study results (e.g. subgroup analysis, meta-regression). | Methods and Appendix 5 |
|  | 13f | Describe any sensitivity analyses conducted to assess robustness of the synthesized results. | Methods and Appendix 5 |
| **Reporting bias assessment** | 14 | Describe any methods used to assess risk of bias due to missing results in a synthesis (arising from reporting biases). | Not applicable |
| **Certainty assessment** | 15 | Describe any methods used to assess certainty (or confidence) in the body of evidence for an outcome. | Methods |
| **RESULTS** | | | |
| **Study selection** | 16a | Describe the results of the search and selection process, from the number of records identified in the search to the number of studies included in the review, ideally using a flow diagram. | Results and Figure 1 |
|  | 16b | Cite studies that might appear to meet the inclusion criteria, but which were excluded, and explain why they were excluded. | Results and Figure 1 |
| **Study characteristics** | 17 | Cite each included study and present its characteristics. | Appendix 3 |
| **Risk of bias in studies** | 18 | Present assessments of risk of bias for each included study. | Not applicable |
| **Results of individual studies** | 19 | For all outcomes, present, for each study: (a) summary statistics for each group (where appropriate) and (b) an effect estimate and its precision (e.g. confidence/credible interval), ideally using structured tables or plots. | Appendix 4.2 |
| **Results of syntheses** | 20a | For each synthesis, briefly summarise the characteristics and risk of bias among contributing studies. | Not applicable |
|  | 20b | Present results of all statistical syntheses conducted. If meta-analysis was done, present for each the summary estimate and its precision (e.g. confidence/credible interval) and measures of statistical heterogeneity. If comparing groups, describe the direction of the effect. | Results and Appendix 4 |
|  | 20c | Present results of all investigations of possible causes of heterogeneity among study results. | Results and Appendix 4 and 5 |
|  | 20d | Present results of all sensitivity analyses conducted to assess the robustness of the synthesized results. | Results and Appendix 5 |
| **Reporting biases** | 21 | Present assessments of risk of bias due to missing results (arising from reporting biases) for each synthesis assessed. | Not applicable |
| **Certainty of evidence** | 22 | Present assessments of certainty (or confidence) in the body of evidence for each outcome assessed. | Results |
| **DISCUSSION** | | | |
| **Discussion** | 23a | Provide a general interpretation of the results in the context of other evidence. | Discussion |
|  | 23b | Discuss any limitations of the evidence included in the review. | Discussion |
|  | 23c | Discuss any limitations of the review processes used. | Discussion |
|  | 23d | Discuss implications of the results for practice, policy, and future research. | Discussion |
| **OTHER INFORMATION** | | | |
| **Registration and protocol** | 24a | Provide registration information for the review, including register name and registration number, or state that the review was not registered. | Methods |
|  | 24b | Indicate where the review protocol can be accessed, or state that a protocol was not prepared. | Methods |
|  | 24c | Describe and explain any amendments to information provided at registration or in the protocol. | Not applicable |
| **Support** | 25 | Describe sources of financial or non-financial support for the review, and the role of the funders or sponsors in the review. | Abstract |
| **Competing interests** | 26 | Declare any competing interests of review authors. | Abstract |
| **Availability of data, code and other materials** | 27 | Report which of the following are publicly available and where they can be found: template data collection forms; data extracted from included studies; data used for all analyses; analytic code; any other materials used in the review. | Methods |

**Table 2.2.2.** PRISMA abstract checklist

| Topic | No. | Summary | Reported? |
| --- | --- | --- | --- |
| **Title** | 1 | Identify the report as a systematic review. | Yes |
| **BACKGROUND** | | | |
| **Objectives** | 2 | Provide an explicit statement of the main objective(s) or question(s) the review addresses. | Yes |
| **METHODS** | | | |
| **Eligibility criteria** | 3 | Specify the inclusion and exclusion criteria for the review. | Yes |
| **Information sources** | 4 | Specify the information sources (e.g. databases, registers) used to identify studies and the date when each was last searched. | Yes |
| **Risk of bias** | 5 | Specify the methods used to assess risk of bias in the included studies. | Yes |
| **Synthesis of results** | 6 | Specify the methods used to present and synthesize results. | Yes |
| **RESULTS** | | | |
| **Included studies** | 7 | Give the total number of included studies and participants and summarise relevant characteristics of studies. | Yes |
| **Synthesis of results** | 8 | Present results for main outcomes, preferably indicating the number of included studies and participants for each. If meta-analysis was done, report the summary estimate and confidence/credible interval. If comparing groups, indicate the direction of the effect (i.e. which group is favoured). | Yes |
| **DISCUSSION** | | | |
| **Limitations of evidence** | 9 | Provide a brief summary of the limitations of the evidence included in the review (e.g. study risk of bias, inconsistency and imprecision). | Yes |
| **Interpretation** | 10 | Provide a general interpretation of the results and important implications. | Yes |
| **OTHER** | | | |
| **Funding** | 11 | Specify the primary source of funding for the review. | Yes |
| **Registration** | 12 | Provide the register name and registration number. | Yes |

#### 2.3. Search strategy by data source

**Table 2.3.** Search strategy by data source used in the 2024 updated systematic review

| Data source | Search strategy |
| --- | --- |
| PubMed (National  Center for  Biotechnology  Information) | (({"HIV lnfections"[Mesh] OR "HIV"[Mesh] OR "Acquired Immunodeficiency  Syndrome"[Mesh] OR "Antiretroviral Therapy, Highly Active"[Mesh] OR "Anti­  HIV Agents"[Mesh] OR "Anti-Retroviral Agents"[Mesh] OR "human  immunodeficiency virus"[ti] OR "human immunedeficiency virus"[ti] OR  "human immuno deficiency virus"[ti] OR "human immune deficiency virus"[ti]  OR HIV[ti] OR HIVl[ti] OR HIV2[ti] OR "acquired immunodeficiency  syndrome"[ti] OR "acquired immunedeficiency syndrome"[ti] OR "acquired  immunodeficiency syndrome"[ti] OR "acquired immune deficiency  syndrome"[ti] OR antiretroviral*[ti] OR "anti retroviral"[ti]) **AND**  {"Pregnancy"[Mesh] OR "Pregnancy Complications, lnfectious"[Mesh] OR  "Pregnant Women"[Mesh] OR "Delivery, Obstetric"[Mesh] OR "Peripartum  Period"[Mesh] OR "Postpartum Period"[Mesh] OR "Breast Feeding"[Mesh] OR  "Infectious Disease Transmission, Vertical"[Mesh] OR PMTCT[tiab] OR  MTCT[tiab] OR "mother to child"[tiab] OR "parent to child"[tiab] OR  vertical*[tiab] OR intrauterine[tiab] OR "intra uterine"[tiab] OR  intrapartum[tiab] OR "intra partum"[tiab] OR pregnant[tiab] OR  pregnancy[tiab] OR prenatal*(tiab] OR "pre natal*"(tiab] OR antenatal*(tiab]  OR "ante natal*"(tiab] OR perinatal*(tiab] OR "peri natal*"(tiab] OR  puerperium[tiab] OR postnatal*[tiab] OR "post natal*"[tiab] OR  postpartum[tiab] OR "post partum"[tiab] OR peripartum[tiab] OR "peri  partum"[tiab] OR "in utero"[tiab] OR fetomaternal*[tiab] OR "feto  maternal*"[tiab] OR "maternal fetal"[tiab] OR fetus*[tiab] OR foetus*[tiab]  OR fetal*[tiab] OR foetal*[tiab] OR neonat*[tiab] OR breastfeed*[tiab] OR  "breast feeding"[tiab] OR "breast fed"[tiab] OR breastmilk[tiab] OR "breast  milk"[tiab] OR delivery[tiab] OR birth[tiab]) **AND** {"lnfant"[Mesh] OR "Infant,  Newborn"[Mesh] OR "Child"[Mesh] OR infant[tiab] OR infants[tiab] OR  infancy[tiab] OR newborn*(tiab] OR "new born"[tiab] OR neonat*(tiab] OR  child*(tiab] OR baby[tiab] OR babies[tiab]) **AND** {"Infectious Disease  Transmission, Vertical"[Mesh] OR transmit*[tiab] OR transmission*[tiab] OR  infection* OR infected)) **OR** (("HIV lnfections"[Mesh] OR "HIV  infection*"[tiab] OR HIV[ti]) **AND** {"Pregnancy Complications,  lnfectious"[Mesh] OR "Infectious Disease Transmission, Vertical"[Mesh] OR "vertical transmission"[tiab:~3] OR "vertical infection"[tiab:~3] OR "vertical  infections"[tiab:~3] OR "mother to child" OR "parent to child" OR MTCT OR  PMTCT OR "perinatal transmission"[tiab:~3] OR "perinatal infection"[tiab:~3]  OR "perinatally acquired"[tiab:~3]))) **AND** {"2018"[Date - Publication] :  "3000"[Date - Publication]) **AND** "English"[la] **NOT** (("Animals"[Mesh] OR macaque*[tiab]) NOT "Humans"[Mesh]) **NOT** (("Cross-Sectional Studies"[Mesh] OR "editorial"[Publication Type] OR "letter"[Publication Type] OR "comment"[Publication Type] OR "news"[Publication Type] OR "Case Reports" [Publication Type] OR "Case Reports as Topic"[Mesh]) NOT ("Systematic Review" [Publication Type] OR "Meta-Analysis" [Publication  Type])) |
| Embase (Elsevier) | ((('Human immunodeficiency virus infection'/de OR 'acquired immune deficiency syndrome'/de OR 'AIDS related complex'/de OR 'acute HIV infection'/de OR 'Human immunodeficiency virus 1 infection'/de OR 'Human immunodeficiency virus 2 infection'/de OR 'AIDS related complex'/de OR 'human immunodeficiency virus'/exp OR 'highly active antiretroviral therapy'/exp OR 'anti human immunodeficiency virus agent'/de OR 'antiretrovirus agent'/de OR 'human immunodeficiency virus':ti OR 'human immunedeficiency virus':ti OR 'human immunodeficiency virus':ti OR 'human immune deficiency virus':ti OR 'hiv':ti OR 'hivl':ti OR 'hiv2':ti OR 'acquired immunodeficiency syndrome':ti OR 'acquired immunedeficiency syndrome':ti OR 'acquired immuno deficiency syndrome':ti OR 'acquired immune deficiency syndrome':ti OR 'antiretroviral*':ti OR 'anti retroviral':ti) **AND** ('pregnancy'/exp OR 'infectious pregnancy complications'/exp OR 'pregnant woman'/exp OR 'obstetric delivery'/exp OR 'perinatal period'/exp OR 'puerperium'/exp OR 'breast feeding'/exp OR 'vertical transmission'/exp OR 'pmtct':ti,ab,kw OR 'mtct':ti,ab,kw OR 'mother to child':ti,ab,kw OR 'parent to child':ti,ab,kw OR 'vertical*':ti,ab,kw OR 'intrauterine':ti,ab,kw OR 'intra uterine':ti,ab,kw OR 'intrapartum':ti,ab,kw OR 'intra partum':ti,ab,kw OR 'pregnant':ti,ab,kw OR 'pregnancy':ti,ab,kw OR 'prenatal*':ti,ab,kw OR 'pre natal*':ti,ab,kw OR 'antenatal*':ti,ab,kw OR 'ante natal*':ti,ab,kw OR 'perinatal*':ti,ab,kw OR 'peri natal*':ti,ab,kw OR 'puerperium':ti,ab,kw OR 'postnatal*':ti,ab,kw OR 'post natal*':ti,ab,kw OR 'postpartum':ti,ab,kw OR 'post partum':ti,ab,kw OR 'peripartum':ti,ab,kw OR 'peri partum':ti,ab,kw OR 'in utero':ti,ab,kw OR 'fetomaternal*':ti,ab,kw OR 'feto maternal*':ti,ab,kw OR 'maternal fetal':ti,ab,kw OR 'fetus*':ti,ab,kw OR 'foetus*':ti,ab,kw OR 'fetal*':ti,ab,kw OR 'foetal*':ti,ab,kw OR 'neonat*':ti,ab,kw OR 'breastfeed*':ti,ab,kw OR 'breast feeding':ti,ab,kw OR 'breast fed':ti,ab,kw OR 'breastmilk':ti,ab,kw OR 'breast milk':ti,ab,kw OR 'delivery':ti,ab,kw OR 'birth':ti,ab,kw) **AND** ('infant'/exp OR 'newborn'/exp OR 'child'/exp OR 'infant':ti,ab,kw OR 'infants':ti,ab,kw OR 'infancy':ti,ab,kw OR 'newborn*':ti,ab,kw OR 'new born':ti,ab,kw OR 'neonat*':ti,ab,kw OR 'child*':ti,ab,kw OR 'baby':ti,ab,kw OR 'babies':ti,ab,kw) **AND** ('vertical transmission'/exp OR 'transmit*':ti,ab,kw OR 'transmission*':ti,ab,kw OR 'infection*' OR 'infected')) **OR** (('human immunodeficiency virus infection'/exp OR 'HIV infection*':ti,ab OR hiv:ti) **AND** ('infectious pregnancy complications'/exp OR 'vertical transmission'/exp OR (vertical NEAR/3 (transmission OR infection*)):ti,ab,kw OR 'mother to child':ti,ab,kw OR 'parent to child':ti,ab,kw OR MTCT OR PMTCT OR (perinatal* NEAR/3 (transmission OR infection OR acquired)):ti,ab,kw))) **AND** [english]/lim **AND** (2018-2024]/py **NOT** (('animal'/exp OR 'macaque*':ti,ab,kw) NOT  'human'/exp) **NOT** (('cross-sectional study'/exp OR 'editorial'/exp OR 'letter'/exp OR 'note'/exp OR 'case study'/exp OR 'case study':ti OR 'case report*':ti OR 'cross sectional':ti,ab) NOT ('systematic review'/exp OR 'systematic review (topic)'/exp OR 'meta analysis'/exp OR 'meta analysis (topic)'/exp)) **NOT** 'conference abstract'/exp |
| CINAHL Complete  (EBSCO) | (((MH "HIV Infections" OR MH "HIV Seropositivity" OR MH "Acquired Immunodeficiency Syndrome" OR MH "Human Immunodeficiency Virus+" OR MH "HIV-Positive Persons+" OR MH "Antiretroviral Therapy, Highly Active" OR MH "Anti-HIV Agents+" OR MH "Anti-Retroviral Agents+" OR Tl ("human immunodeficiency virus" OR "human immunedeficiency virus" OR "human immunodeficiency virus" OR "human immune deficiency virus" OR HIV OR HIVl OR HIV2 OR "acquired immunodeficiency syndrome" OR "acquired immunedeficiency syndrome" OR "acquired immunodeficiency syndrome" OR "acquired immune deficiency syndrome" OR antiretroviral* OR "anti retroviral")) **AND** (MH "Pregnancy+" OR MH "Childbirth+" OR MH "Pregnancy Complications, Infectious+" OR MH "Expectant Mothers" OR MH "Perinatal Period" OR MH "Postnatal Period+" OR MH "Breast Feeding+" OR MH  "Disease Transmission, Vertical" OR Tl (PMTCT OR MTCT OR "mother to child"  OR "parent to child" OR vertical* OR intrauterine OR "intra uterine" OR intrapartum OR "intra partum" OR pregnant OR pregnancy OR prenatal* OR "pre natal*" OR antenatal* OR "ante natal*" OR perinatal* OR "peri natal*" OR puerperium OR postnatal* OR "post natal*" OR postpartum OR "post partum" OR peripartum OR "peri partum" OR "in utero" OR fetomaternal* OR "feto maternal*" OR "maternal fetal" OR fetus* OR foetus* OR fetal* OR foetal* OR neonat* OR breastfeed* OR "breast feeding" OR "breast fed" OR breastmilk OR "breast milk" OR delivery OR birth) OR AB (PMTCT OR MTCT OR "mother to child" OR "parent to child" OR vertical* OR intrauterine OR "intra uterine" OR intrapartum OR "intra partum" OR pregnant OR pregnancy OR prenatal* OR "pre natal*" OR antenatal* OR "ante natal*" OR perinatal* OR "peri natal*" OR puerperium OR postnatal* OR "post natal*" OR postpartum OR "postpartum" OR peripartum OR "peri partum" OR "in utero" OR fetomaternal* OR "feto maternal*" OR "maternal fetal" OR fetus* OR foetus* OR fetal* OR foetal* OR neonat* OR breastfeed* OR "breast feeding" OR "breast fed" OR breastmilk OR "breast milk" OR delivery OR birth)) **AND** (MH "Infant+" OR MH "Infant, Newborn+" OR MH "Child+" OR Tl (infant OR infants OR infancy OR newborn* OR "new born" OR neonat* OR child* OR baby OR babies) OR AB (infant OR infants OR infancy OR newborn* OR "new born" OR neonat* OR child* OR baby OR babies)) **AND** (MH "Disease Transmission, Vertical" OR Tl (transmit* OR transmission* OR infection* OR infected) OR AB (transmit* OR transmission*))) **OR** ((MH "HIV Infections" OR Tl("HIV infection*" OR HIV) OR AB("HIV infection*")) **AND** (MH "Pregnancy Complications, Infectious+" OR MH "Disease Transmission, Vertical" OR (Vertical N3 (transmission OR infection*)) OR "mother to child" OR "parent to child" OR (perinatal N3 (transmission OR infection*)) OR (perinatally N3 acquired) OR MTCT OR PMTCT))) **AND** PY 2018-2024 **AND** LA "English" **NOT**  ((MH "Animals") NOT (MH "Human")) **NOT** ((MH "Cross Sectional Studies" OR  MH "Case Studies" OR Tl ("case report" OR "case reports" OR "case series" OR "cross sectional") OR PT (commentary OR editorial OR letter)) NOT PT ("systematic review" OR "meta analysis")) NOT PT abstract |
| Global Health  (EBSCO) | Limit: Publication Year 2018-2024  ((((DE "HIV infections" OR DE "HIV-1 infections" OR DE "HIV-2 infections" OR DE "human immunodeficiency viruses" OR DE "Human immunodeficiency virus 1" OR DE "Human immunodeficiency virus 2" OR DE "acquired immune deficiency syndrome" OR DE "antiretroviral agents" OR DE "reverse transcriptase inhibitors" OR Tl ("human immunodeficiency virus" OR "human immunedeficiency virus" OR "human immunodeficiency virus" OR "human immune deficiency virus" OR HIV OR HIVl OR HIV2 OR "acquired immunodeficiency syndrome" OR "acquired immunedeficiency syndrome" OR "acquired immunodeficiency syndrome" OR "acquired immune deficiency syndrome" OR antiretroviral* OR "anti retroviral")) **AND** (DE "pregnancy" OR DE "birth" OR DE "childbirth" OR DE "postpartum period" OR DE "pregnancy complications" OR DE "parturition" OR DE "prenatal period" OR DE "prepartum period" OR DE "puerperium" OR DE "fetus" OR DE "breast feeding" OR DE "human milk" OR DE "vertical transmission" OR DE "maternal transmission" OR Tl (PMTCT OR MTCT OR "mother to child" OR "parent to child" OR vertical* OR intrauterine OR "intra uterine" OR intrapartum OR "intra partum" OR pregnant OR pregnancy OR prenatal* OR "pre natal*" OR antenatal* OR "ante natal*" OR perinatal* OR "perinatal*" OR puerperium OR postnatal* OR "post natal*" OR postpartum OR "postpartum" OR peripartum OR "peri partum" OR "in utero" OR fetomaternal* OR "feto maternal*" OR "maternal fetal" OR fetus* OR foetus* OR fetal* OR foetal* OR neonat* OR breastfeed* OR "breast feeding" OR "breast fed" OR breastmilk OR "breast milk" OR delivery OR birth) OR AB (PMTCT OR MTCT OR "mother to child" OR "parent to child" OR vertical* OR intrauterine OR "intra uterine" OR intrapartum OR "intra partum" OR pregnant OR pregnancy OR prenatal* OR "pre natal*" OR antenatal* OR "ante natal*" OR perinatal* OR "peri natal*" OR puerperium OR postnatal* OR "post natal*" OR postpartum OR "postpartum" OR peripartum OR "peri partum" OR "in utero" OR fetomaternal* OR "feto maternal*" OR "maternal fetal" OR fetus* OR foetus* OR fetal* OR foetal* OR neonat* OR breastfeed* OR "breast feeding" OR "breast fed" OR breastmilk OR "breast milk" OR delivery OR birth)) **AND** (DE "children" OR DE "preschool children" OR DE "school children" OR DE "infants" OR DE "neonates" OR DE "neonates" OR Tl (infant OR infants OR infancy OR newborn* OR "new born" OR neonat* OR child* OR baby OR babies) OR AB (infant OR infants OR infancy OR newborn* OR "new born" OR neonat* OR child* OR baby OR babies)) **AND** (DE "vertical transmission" OR DE "maternal transmission" OR Tl (transmit* OR transmission* OR infection* OR infected) OR AB (transmit* OR transmission*))) **OR** ((DE "HIV infections" OR DE "HIV-1 infections" OR DE "HIV-2 infections" OR Tl("HIV infection*" OR HIV) OR AB("HIV infection*")) **AND** (DE "vertical transmission" OR DE "maternal transmission" OR (Vertical N3 (transmission OR infection)) OR "mother to child" OR "parent to child" OR (perinatal N3 (transmission OR  infection)) OR (perinatally N3 acquired) OR MTCT OR PMTCT))) **AND** LA "English" **NOT** ((DE "Animals" OR DE "Laboratory Animals") NOT DE "Hominidae") **NOT** ((Tl "cross sectional" OR AB "cross sectional" OR ZT "editorial" OR ZT "letter" OR DE "case reports" OR Tl "case report" OR Tl "case reports" OR Tl "case series") NOT (ZU "systematic reviews" OR ZU  "meta-analysis")) |
| Cochrane CENTRAL  (Wiley) | Limit - language: English  ID Search Hits  #1 MeSH descriptor: [HIV Infections] explode all trees 17667  #2 MeSH descriptor: [HIV] explode all trees 4211  #3 MeSH descriptor: [Anti-Retroviral Agents] explode all trees 6112  #4 MeSH descriptor: [Anti-HIV Agents] explode all trees 5128  #5 MeSH descriptor: [Antiretroviral Therapy, Highly Active] explode all  trees 1626  #6 (HIV NEXT infection*) OR "human immunodeficiency virus" OR  "human immunedeficiency virus" OR "human immune deficiency virus" OR  "human immunodeficiency virus" OR HIV OR HIVl OR HIV2 OR "acquired  immunodeficiency syndrome" OR "acquired immunedeficiency syndrome" OR  "acquired immune deficiency syndrome" OR "acquired immunodeficiency  syndrome" OR antiretroviral* OR "anti retroviral" 34501  #7 MeSH descriptor: [Pregnancy] explode all trees 33699  #8 MeSH descriptor: [Pregnancy Complications, Infectious] explode all  trees 1616  #9 MeSH descriptor: [Pregnant Women] explode all trees 988  #10 MeSH descriptor: [Delivery, Obstetric) explode all trees 7441  #11 MeSH descriptor: [Peripartum Period] explode all trees 47  #12 MeSH descriptor: [Postpartum Period) explode all trees 2754  #13 MeSH descriptor: [Breast Feeding] explode all trees 2878  #14 MeSH descriptor: [Infectious Disease Transmission, Vertical] explode  all trees 842  #15 PMTCT OR MTCT OR "mother to child" OR "parent to child" OR  vertical* OR intrauterine OR "intra uterine" OR intrapartum OR "intra  partum" OR pregnant OR pregnancy OR prenatal* OR (pre NEXT natal*) OR  antenatal* OR (ante NEXT natal*) OR perinatal* OR (peri NEXT natal*) OR  puerperium OR postnatal* OR (post NEXT natal*) OR postpartum OR "post  partum" OR peripartum OR "peri partum" OR "in utero" OR fetomaternal* OR  (feto NEXT maternal*) OR "maternal fetal" OR fetus* OR foetus* OR fetal* OR  foetal* OR neonat* OR breastfeed* OR "breast feeding" OR "breast fed" OR  breastmilk OR "breast milk" OR delivery OR birth 186277  #16 MeSH descriptor: [Infant] explode all trees 45750  #17 MeSH descriptor: [Child) explode all trees 81197  #18 infant OR infants OR infancy OR newborn* OR "new born" OR  neonat* OR child* OR baby OR babies 265502  #19 transmit* OR transmission* OR infection* OR infected 167225  #20 (#1 OR #2 OR #3 OR #4 OR #5 OR #6) AND (#7 OR #8 OR #9 OR #10 OR  #11 OR #12 OR #13 OR #14 OR #15) AND (#16 OR #17 OR #18) AND (#14 OR  #19) 2943  #21 (HIV NEXT infection*) OR HIV 33237  #22 (vertical NEAR/3 transmission) OR "mother to child" OR "parent to child" OR MTCT OR PMTCT OR (perinatal NEAR/3 transmission) OR (perinatal NEAR/3 infection*) OR (perinatally NEAR/3 acquired) 1845  #23 (#1 OR #21) AND #22 1327  #24 #20 OR #23 with Publication Year from 2018 to 2024, in Trials 773 |
| Global Index Medicus  (World Health  Organization) | (tw:(hiv OR "human immunodeficiency virus"))  AND  (tw:((pregnan* OR childbirth OR breast* OR perinatal* OR prenatal* OR utero  OR vertical* OR "mother to child")))  AND  (tw:(transmit* OR transmission))  AND  La:("en")  AND  Year_cluster:[2018 to 2024] |

#### 2.4. Variables extracted in systematic review

The following variables were extracted from all included studies:

*Study details:*

- Author name(s)
- Study title
- Journal
- Publication year
- Geographic region(s) covered (i.e. country or countries)
- Dates of data collection
- Study population(s)
- Total population size

*Vertical transmission details:*

- Total N of pregnant women or breastfeeding mothers living with HIV
- Total N of infants born to mothers living with HIV, who were tested for HIV
- N of infants who tested HIV-positive
- Events of vertical HIV transmission
- Timing of transmission (perinatal or during breastfeeding)
- Age of child at testing
- Maternal prophylaxis

*When available:*

- Maternal ART regimens
- Timing of maternal ART initiation
- Infant treatment and prophylaxis
- Maternal viral load or viral suppression information
  a. Timing of viral load test
  b. In viral load suppression, threshold for viral load suppression
- Maternal CD4 count information at baseline
- Infant feeding patterns (breastfed or formula fed, including duration in months)

#### 2.5. Perinatal transmission probability definition

Perinatal transmission was defined as transmission that occurs before six weeks (1.5 months) after birth. We included studies with both breastfeeding and formula-feeding populations. To model perinatal VT probability, we extracted the number of infections identified before six weeks postpartum and the number of HIV exposed infants.

$$PVT= \frac{{HPI}_{1.5 months}}{HEI}$$

Equation 2.2

Perinatal VT (*PVT*) probability was then calculated as the ratio of HIV positive infants (${HPI}_{1.5 months}$) to HIV exposed infants (*HEI*) as shown in Equation 2.2.

#### 2.6. Monthly breastfeeding transmission probability definition

Breastfeeding transmission was calculated as a monthly transmission probability and considered any vertical transmission that occurred in breastfeeding populations for infants after six weeks of age. The monthly breastfeeding transmission probability (*BFVT*) was then calculated according to Equation 2.3.

$$BFVT= \frac{{HPI}_{{BF}_{End}}- {HPI}_{1.5 months}}{(HEI- {HPI}_{1.5 months})*({BF}_{End}-1.5)}$$

Equation 2.3

The numerator represents the number of infections that occurred between the end of breastfeeding and the end of the perinatal period as the difference between the number of HIV positive infants at the end of breastfeeding (${HPI}_{{BF}_{End}}$) and at 1.5 months (${HPI}_{1.5 months}$). The denominator represents the number of HIV exposed (*HEI*) but uninfected infants during this period (minus ${HPI}_{1.5 months}$) and the number of months between the perinatal period (1.5) and the end of breastfeeding (${BF}_{End}$).

### 3. Meta-regression model estimates

#### 3.1 Regression tables for all models

**Table 3.1.1.** Regression table for models one, two, three, and four used to estimate VT probabilities compatible with Spectrum-AIM

| *Model one: VT probability among women not receiving PVT* | | | | | |
| --- | --- | --- | --- | --- | --- |
| Covariate | | **Estimate (logit)** | **95% confidence interval** | **Estimate**  **(odds ratio)** | **95% confidence interval** |
| Intercept | | -1.61 | (-1.82, -1.41) | 0.20 | (0.16, 0.24) |
| CD4 midpoint  (per 100 cells increase, centered on CD4 = 500 mm^3^) | | -0.23 | (-0.28, -0.18) | 0.80 | (0.75, 0.84) |
| Perinatal transmission  (Reference) | | 0.00 | (Reference) | 1.00 | (Reference) |
| Breastfeeding transmission | | -3.23 | (-3.84, -2.61) | 0.04 | (0.02, 0.07) |
| Interaction between CD4 midpoint and breastfeeding transmission | | 0.16 | (-0.22, 0.54) | 1.18 | (0.80, 1.72) |
| *Model two: VT probability among maternal seroconversion or short-course PVT* | | | | | |
| Covariate | | **Estimate (logit)** | **95% confidence interval** | **Estimate (odds)** | **95% confidence interval** |
| Transmission timing | **Category** |  |  |  |  |
| Perinatal | **Infection** | -1.51 | (-1.92, -1.09) | 0.221 | (0.147, 0.337) |
|  | **SDNVP** | -2.40 | (-2.76, -2.06) | 0.091 | (0.063, 0.128) |
|  | **Dual ARV** | -3.44 | (-3.85, -3.01) | 0.032 | (0.021, 0.049) |
|  | **Option A** | -3.43 | (-3.73, -3.13) | 0.032 | (0.024, 0.044) |
|  | **Option B** | -4.02 | (-4.27, -3.77) | 0.018 | (0.014, 0.023) |
| Breastfeeding (monthly) | **Infection** | -0.93 | (-1.34, -0.52) | 0.395 | (0.263, 0.592) |
|  | **SDNVP, <350** | -4.71 | (-6.43, -3.07) | 0.009 | (0.002, 0.047) |
|  | **SDNVP, >350** | -5.64 | (-7.40, -3.92) | 0.004 | (0.001, 0.020) |
|  | **Dual ARV** | -6.21 | (-7.49, -4.89) | 0.002 | (0.001, 0.007) |
|  | **Option A** | -6.18 | (-7.37, -5.07) | 0.002 | (0.001, 0.006) |
|  | **Option B** | -6.58 | (-7.32, -5.81) | 0.001 | (0.001, 0.003) |
| *Model three: Perinatal transmission probability among women receiving ART by timing of initiation* | | | | | |
| Covariate | | **Estimate (logit)** | **95% confidence interval** | **Estimate**  **(odds ratio)** | **95% confidence interval** |
| Intercept | | -4.55 | (-4.79, -4.32) | 0.011 | (0.008, 0.013) |
| Weeks on ART before delivery  (centered on 20 weeks) | | -0.06 | (-0.07, -0.04) | 0.944 | (0.930, 0.958) |
| Late ART initiation  (<4 weeks before delivery) | | 0.68 | (-0.05, 1.45) | 1.974 | (0.952, 4.265) |
| *Model four: Monthly breastfeeding transmission among women receiving lifelong ART* | | | | | |
| Covariate | | **Estimate (logit)** | **95% confidence interval** | **Estimate**  **(odds ratio)** | **95% confidence interval** |
| Intercept  (on ART preconception) | | -8.70 | (-10.13, -7.40) | 0.000 | (0.000, 0.001) |
| ART started during pregnancy | | 2.06 | (0.63, 3.56) | 7.813 | (1.878, 35.245) |

**Table 3.1.2.** Regression table for model three with fixed effects for ART regimen class

| Covariate | Estimate (logit) | | 95% confidence interval | | Estimate (odds ratio) | | 95% confidence interval |
| --- | --- | --- | --- | --- | --- | --- | --- |
| Intercept | -4.45 | (-4.71, -4.19) | | 0.012 | | (0.009, 0.015) | |
| Weeks on ART before delivery  (centered on 20 weeks) | -0.06 | (-0.07, -0.04) | | 0.946 | | (0.932, 0.961) | |
| Late ART initiation  (<4 weeks before delivery) | 0.72 | (-0.05, 1.49) | | 2.050 | | (0.951, 4.418) | |
| ART class |  |  | |  | |  | |
| NNRTI (reference) | 0.00 | (Reference) | | 1.000 | | (Reference) | |
| INSTI | -1.01 | (-1.95, -0.07) | | 0.364 | | (0.142, 0.935) | |
| PI | -0.12 | (-0.56, 0.32) | | 0.885 | | (0.570, 1.374) | |
| Miscellaneous regimens | -0.04 | (-0.67, 0.59) | | 0.957 | | (0.510, 1.795) | |

**Table 4.1.3.** Regression table for model three with fixed effects for ART regimen class and region

| Covariate | Estimate (logit) | | 95% confidence interval | | Estimate (odds ratio) | | 95% confidence interval |
| --- | --- | --- | --- | --- | --- | --- | --- |
| Intercept | -4.31 | (-357, -4.05) | | 0.013 | | (0.010, 0.017) | |
| Weeks on ART before delivery  (centered on 20 weeks) | -0.06 | (-0.07, -0.04) | | 0.946 | | (0.932, 0.961) | |
| Late ART initiation  (<4 weeks before delivery) | 1.05 | (0.31, 1.79) | | 2.865 | | (1.370, 5.992) | |
| ART class |  |  | |  | |  | |
| NNRTI (reference) | 0.00 | (Reference) | | 1.000 | | (Reference) | |
| INSTI | -0.74 | (-1.75, 0.26) | | 0.475 | | (0.174, 1.300) | |
| PI | 0.07 | (-0.37, 0.51) | | 1.076 | | (0.694, 1.670) | |
| Miscellaneous regimens | 0.22 | (-0.29, 0.72) | | 1.242 | | (0.748, 2.062) | |
| Region |  |  | |  | |  | |
| Sub-Saharan Africa (reference) | 0.00 | (Reference) | | 1.000 | | (Reference) | |
| Non Sub-Saharan Africa | -0.58 | (-0.95, -0.21) | | 0.560 | | (0.386, 0.813) | |
| Mixed regions | -0.18 | (-1.07, 0.72) | | 0.838 | | (0.343, 2.048) | |

**Table 3.1.4.** Regression table for viral load suppression model by ART regimen class and timing of ART initiation

| Covariate | | Estimate (logit) | | 95% confidence interval | | Estimate (odds ratio) | | 95% confidence interval |
| --- | --- | --- | --- | --- | --- | --- | --- | --- |
| Intercept | 2.26 | | (1.42, 3.04) | | 9.59 | | (4.16, 21.01) | |
| ART class |  | |  | |  | |  | |
| NNRTI (reference) | 0.00 | | (Reference) | | 1.00 | | (Reference) | |
| INSTI | 0.86 | | (-0.72, 2.51) | | 2.37 | | (0.48, 12.28) | |
| PI | -0.92 | | (-1.73, -0.10) | | 0.40 | | (0.18, 0.91) | |
| Miscellaneous regimens | -0.06 | | (-0.89, 0.76) | | 0.94 | | (0.41, 2.14) | |
| Later ART initiation  (After 1^st^ trimester) | -0.98 | | (-1.58, -0.38) | | 0.37 | | (0.21, 0.68) | |
| Interaction with INSTI | -0.18 | | (-1.99, 1.64) | | 0.84 | | (0.14, 5.14) | |
| Interaction with PI | 0.05 | | (-0.83, 0.94) | | 1.05 | | (0.44, 2.56) | |
| Interaction with miscellaneous regimens | -0.16 | | (-2.51, 2.19) | | 0.85 | | (0.08, 8.98) | |

#### 3.2 Study-level and pooled estimates of vertical transmission probability

Figures 3.2.1 through 3.2.15 presented extracted systematic review data as forest plots, stratified according to the VT probability categories defined by Spectrum-AIM. Data are summarised as forest plots for each stratum.

Data from studies newly identified in the 2024 systematic review are shown in red; studies whereas studies that were in the previous systematic reviews and used in this review are in black, and studies used to calculate the former VT probabilities but were excluded in this analysis are shown in grey.

Studies are summarised as weighted average of the studies used to produce the former VT used in Spectrum-AIM (‘Weighted average: former VT’) and estimates from models two and four fit to the pre-2024 systematic review VT studies (‘Modelled: former VT’). When using the data used to estimate the former VT probabilities, there was insufficient data to fit model one and model three. For each category, we give reasons for excluding studies that had been used in the former VT probabilities but are not used in this analysis.

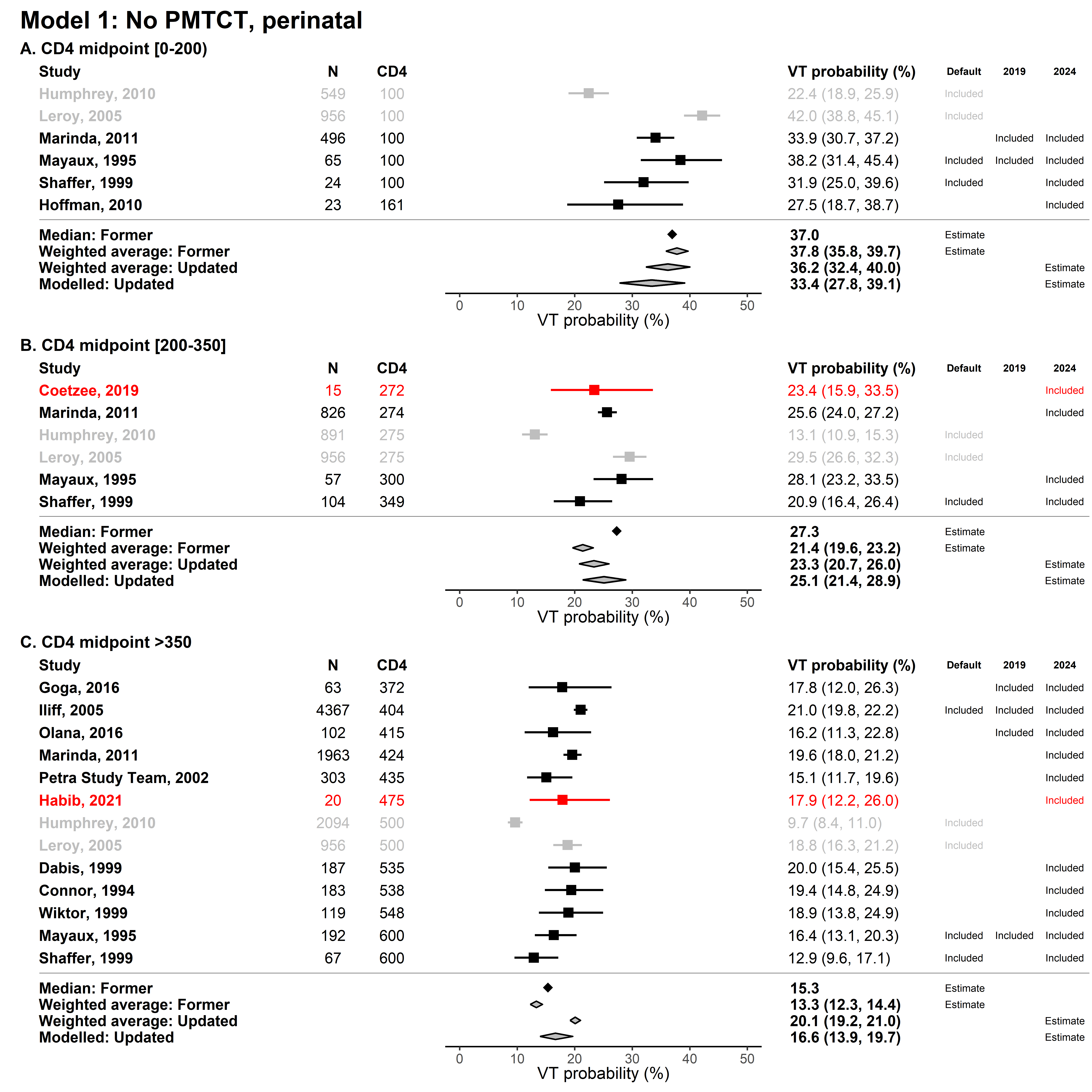

**Figure 3.2.1 Pooled estimates of perinatal VT among women not receiving PVT.** Results are stratified by the Spectrum-AIM defined CD4 ranges. The following pooled estimates are presented: median of studies included in the former VT probabilities (‘Median: former VT’), the weighted average of studies included in the former VT probabilities (‘Weighted average: former VT’), the weighted average of studies included in this analysis (‘Weighted average: Updated’), and the results of the model one (‘Modelled: Updated‘). Studies added in the 2024 review are shown in red and studies excluded from the meta-regression are shown in grey. Humphrey 2010 was excluded as it was not a peer-reviewed study and Leroy 2005 was excluded as it was a pooled analysis using data from other included studies.

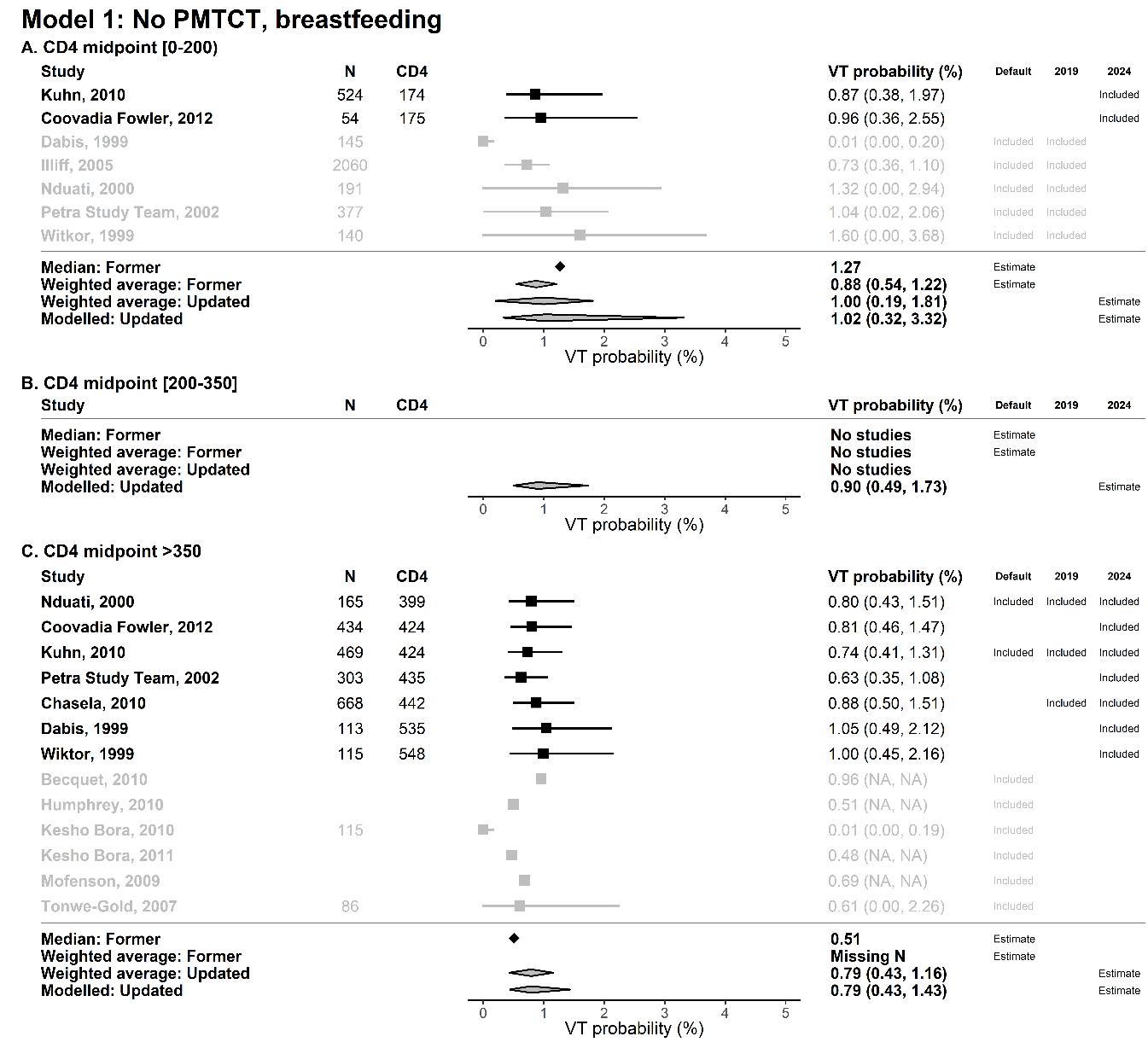

**Figure 3.2.2 Pooled estimates of breastfeeding VT among women not receiving PVT.** Results are stratified by the Spectrum-AIM defined CD4 ranges. The following pooled estimates are presented: median of studies included in the former VT probabilities (‘Median: former VT’), the weighted average of studies included in the former VT probabilities (‘Weighted average: former VT’), the weighted average of studies included in this analysis (‘Weighted average: Updated’), and the results of the model one (‘Modelled: Updated‘). Studies excluded from the meta-regression are shown in grey. Kesho Bora 2010, Kesho Bora 2011, and Tonwe-Gold 2007 were excluded as mothers received AZT. Becquet 2010, Humphrey 2010, and Mofenson 2009 were excluded as they were not peer-reviewed studies. Iliff 2005 was excluded as it was a pooled analysis using data from other included studies.

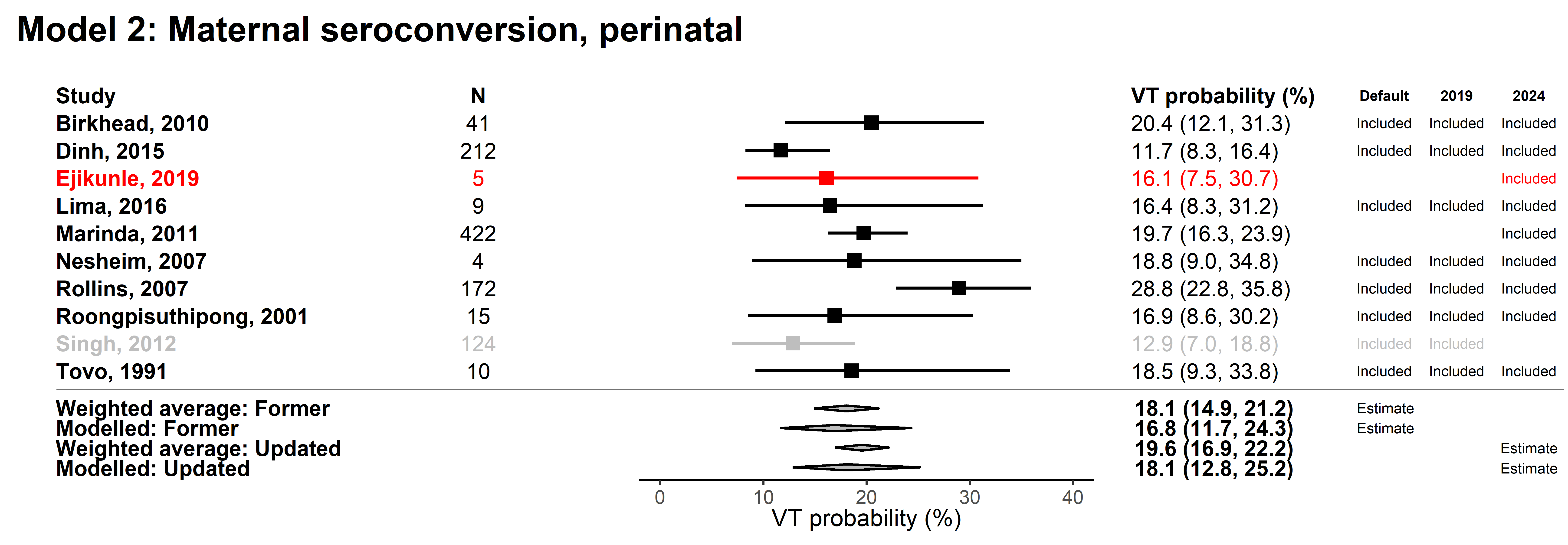

**Figure 3.2.3 Pooled estimates of VT among women who seroconverted during pregnancy.** The following pooled estimates are presented: the weighted average of studies included in the former VT probabilities (‘Weighted average: former VT’), estimates from model two fit to the pre-2024 systematic review studies (‘Modelled: former VT’), the weighted average of studies included in this analysis (‘Weighted average: Updated’), and the results of the model two (‘Modelled: Updated‘). Studies added in the 2024 review are shown in red and studies excluded from the meta-regression are shown in grey. Singh 2012 was excluded as it was not a peer-reviewed study.

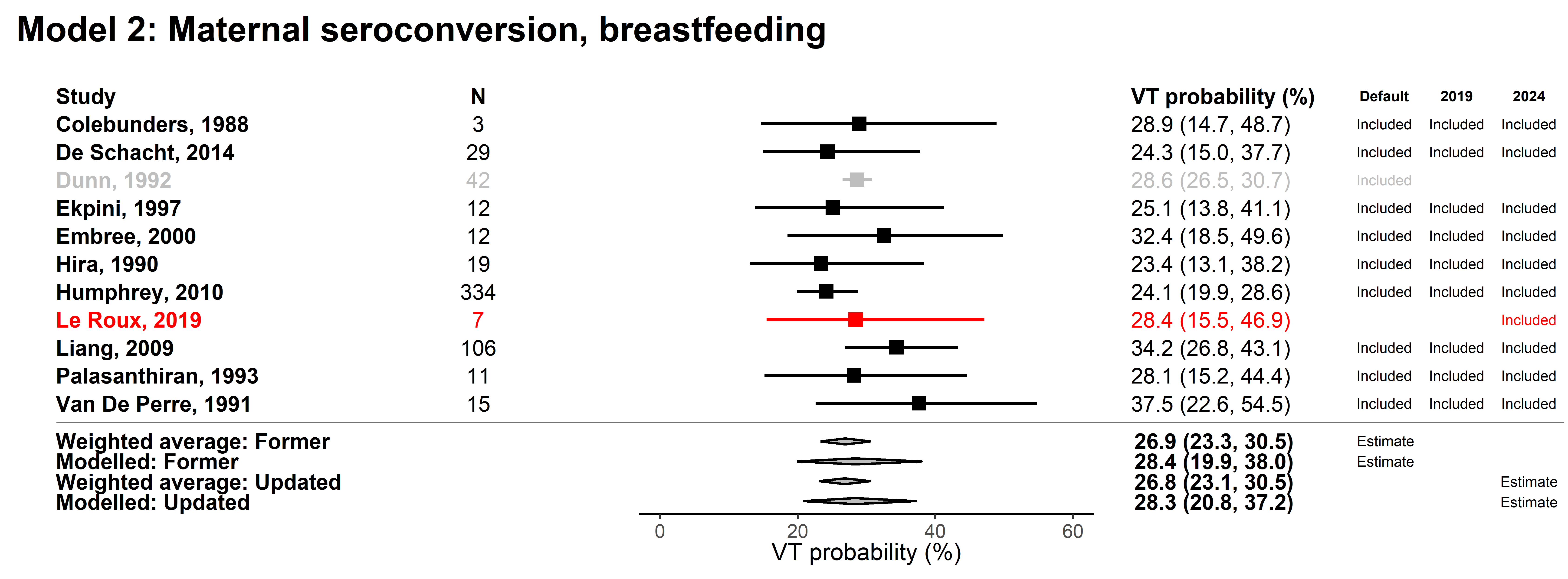

**Figure 3.2.4 Pooled estimates of VT among women who seroconverted during breastfeeding.** The following pooled estimates are presented: the weighted average of studies included in the former VT probabilities (‘Weighted average: former VT’), estimates from model two fit to the pre-2024 systematic review studies (‘Modelled: former VT’), the weighted average of studies included in this analysis (‘Weighted average: Updated’), and the results of the model two (‘Modelled: Updated‘). Studies added in the 2024 review are shown in red and studies excluded from the meta-regression are shown in grey. Singh 2012 was excluded as it was not a peer-reviewed study. Dunn 1992 was excluded as it was a pooled analysis using data from other included studies.

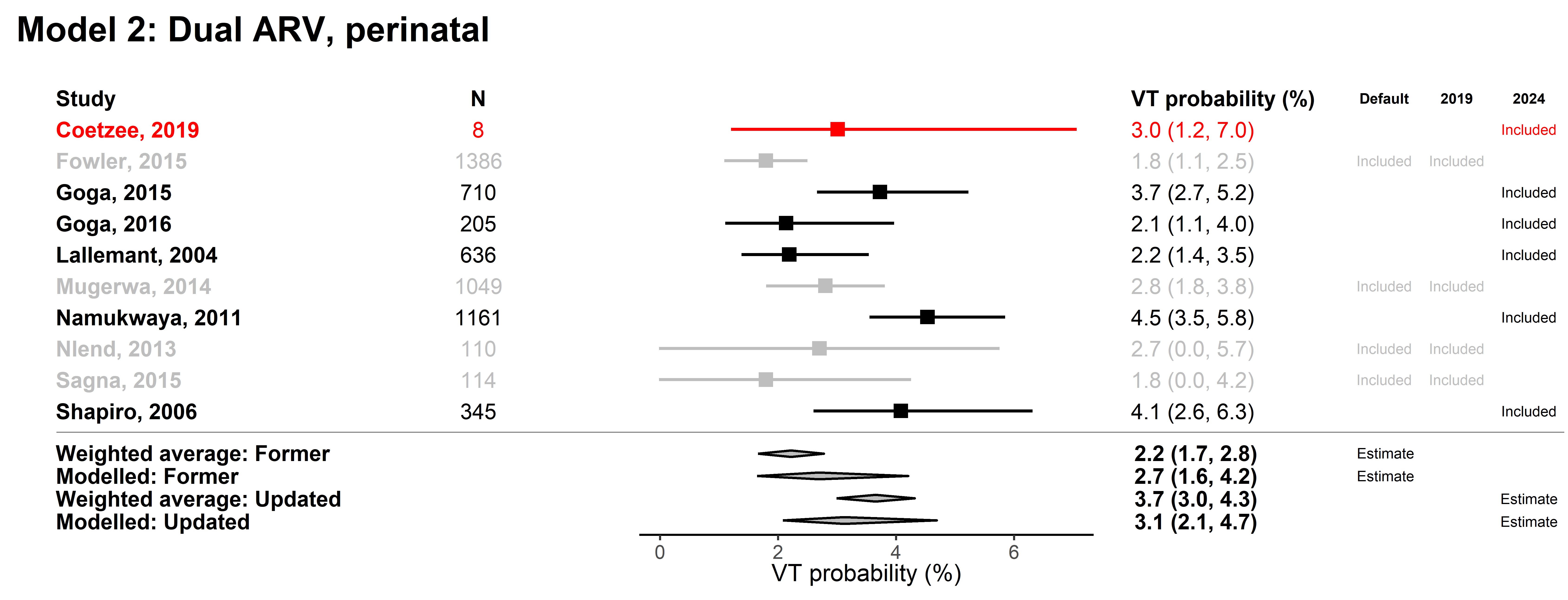

**Figure 3.2.5 Pooled estimates of perinatal VT among women receiving dual ARV.** The following pooled estimates are presented: the weighted average of studies included in the former VT probabilities (‘Weighted average: former VT’), estimates from model two fit to the pre-2024 systematic review studies (‘Modelled: former VT’), the weighted average of studies included in this analysis (‘Weighted average: Updated’), and the results of the model two (‘Modelled: Updated‘). Studies added in the 2024 review are shown in red and studies excluded from the meta-regression are shown in grey. Fowler 2015 and Mugerwa 2014 were excluded as they were not peer-reviewed studies and Nlend 2013 and Sagna 2015 were excluded as they excluded WLHIV with CD4 <350.

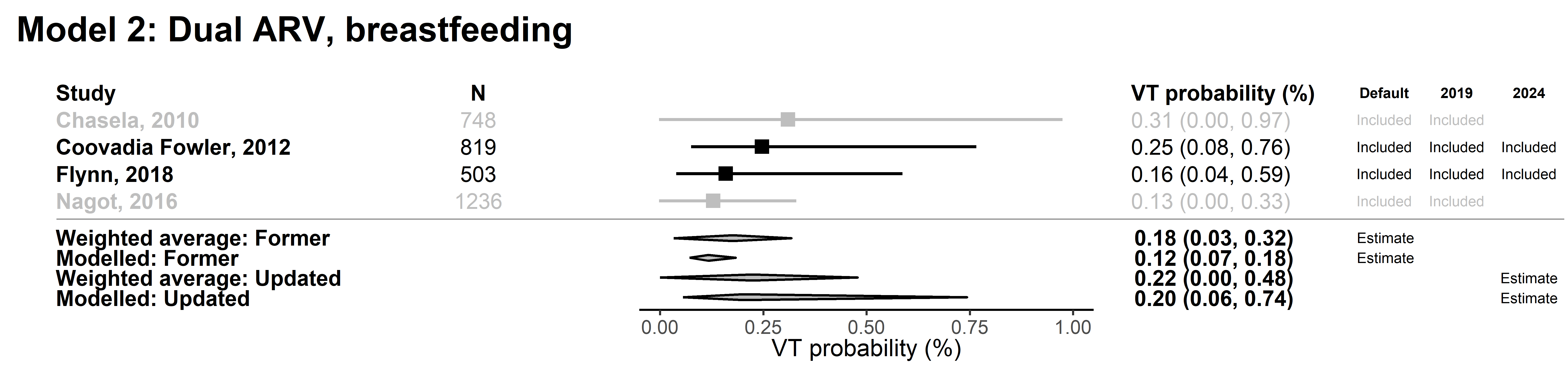
**Figure 3.2.6 Pooled estimates of breastfeeding VT among women receiving dual ARV.** The following pooled estimates are presented: the weighted average of studies included in the former VT probabilities (‘Weighted average: former VT’), estimates from model two fit to the pre-2024 systematic review studies (‘Modelled: former VT’), the weighted average of studies included in this analysis (‘Weighted average: Updated’), and the results of the model two (‘Modelled: Updated‘). Studies excluded from the meta-regression are shown in grey. Chasela 2010 and Nagot 2016 was excluded as they excluded WLHIV with CD4 < 350.

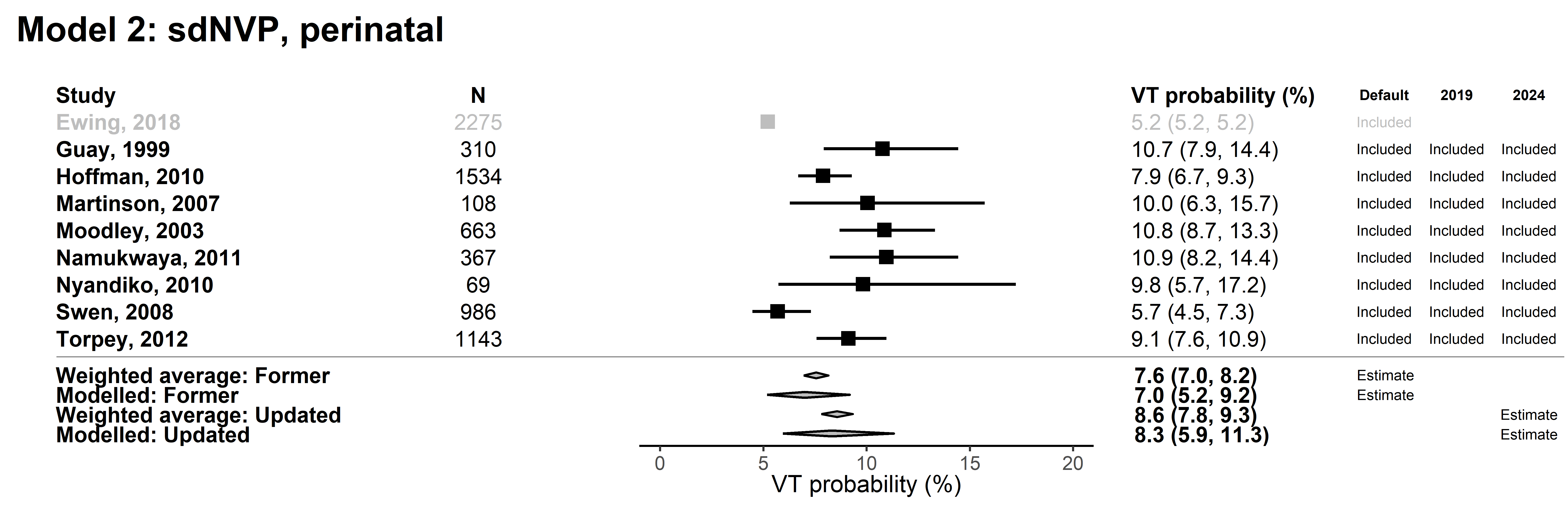

**Figure 3.2.7 Pooled estimates of perinatal VT among women receiving single dose nevirapine.** The following pooled estimates are presented: the weighted average of studies included in the former VT probabilities (‘Weighted average: former VT’), estimates from model two fit to the pre-2024 systematic review studies (‘Modelled: former VT’), the weighted average of studies included in this analysis (‘Weighted average: Updated’), and the results of the model two (‘Modelled: Updated‘). Studies excluded from the meta-regression are shown in grey. Ewing 2018 was excluded as it was not a peer-reviewed study.

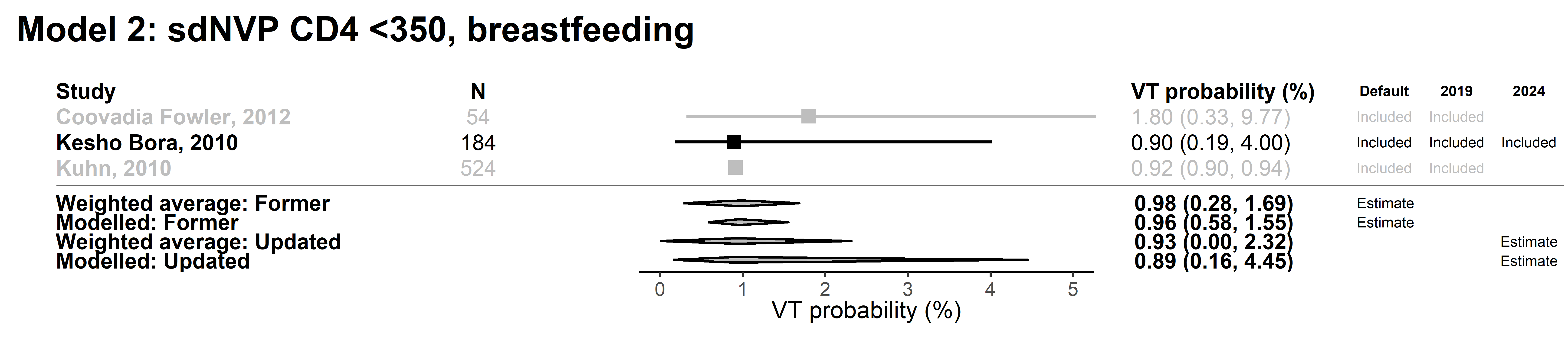

**Figure 3.2.8 Pooled estimates of breastfeeding VT among women with CD4 < 350 receiving single dose nevirapine.** The following pooled estimates are presented: the weighted average of studies included in the former VT probabilities (‘Weighted average: former VT’), estimates from model two fit to the pre-2024 systematic review studies (‘Modelled: former VT’), the weighted average of studies included in this analysis (‘Weighted average: Updated’), and the results of the model two (‘Modelled: Updated‘). Studies excluded from the meta-regression are shown in grey. Kuhn 2010 was excluded as WLHIV were receiving ART.

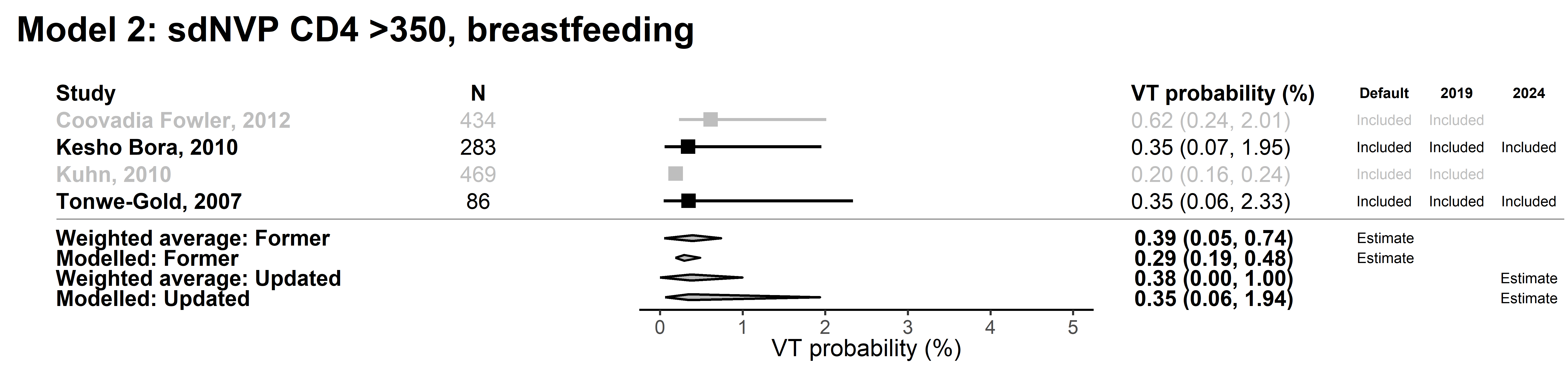

**Figure 3.2.9 Pooled estimates of breastfeeding VT among women with CD4 >350 receiving single dose nevirapine.** The following pooled estimates are presented: the weighted average of studies included in the former VT probabilities (‘Weighted average: former VT’), estimates from model two fit to the pre-2024 systematic review studies (‘Modelled: former VT’), the weighted average of studies included in this analysis (‘Weighted average: Updated’), and the results of the model two (‘Modelled: Updated‘). Studies excluded from the meta-regression are shown in grey. Kuhn 2010 was excluded as WLHIV were receiving ART.

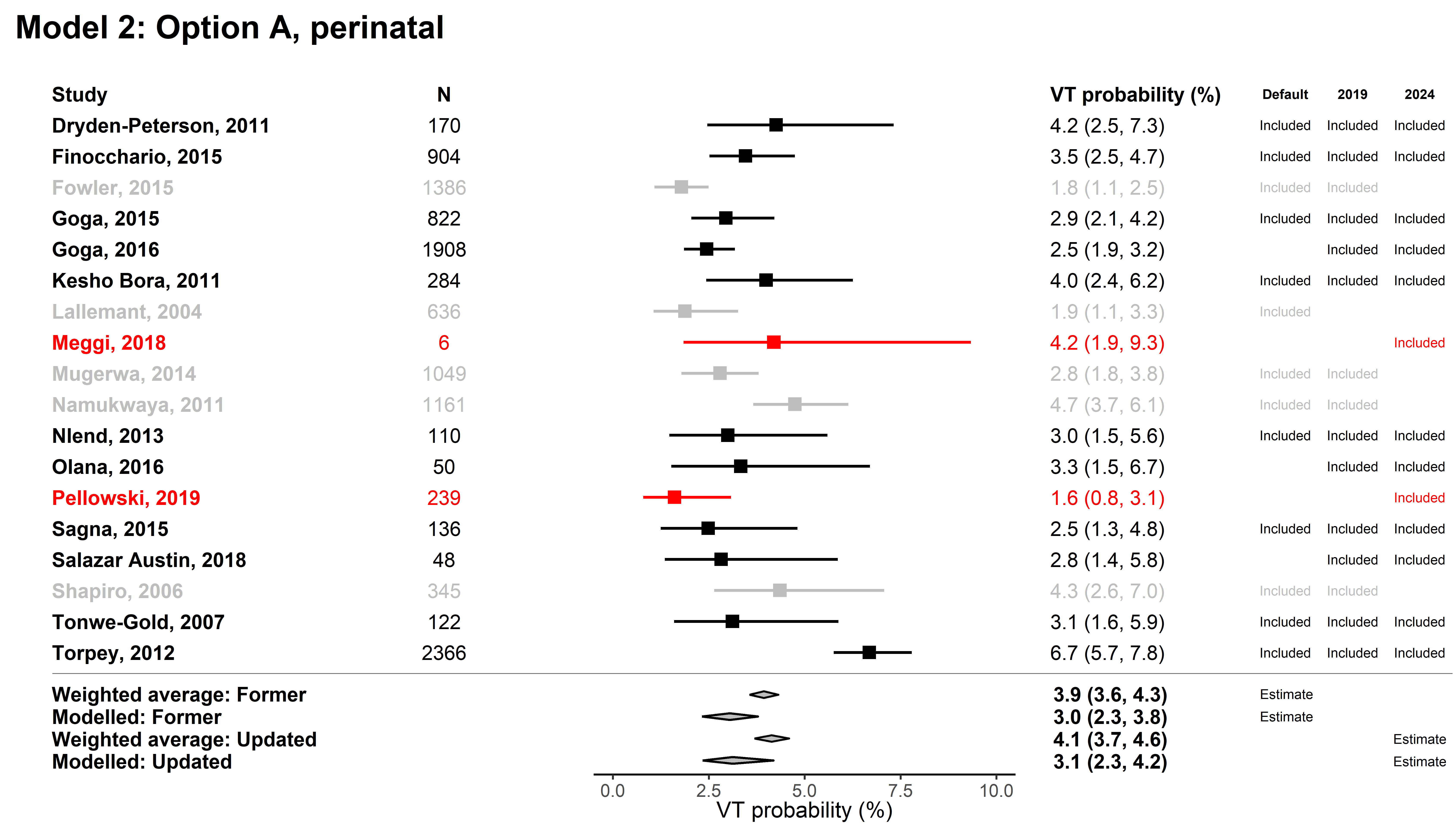

**Figure 3.2.10 Pooled estimates of perinatal VT among women receiving Option A.** The following pooled estimates are presented: the weighted average of studies included in the former VT probabilities (‘Weighted average: former VT’), estimates from model two fit to the pre-2024 systematic review studies (‘Modelled: former VT’), the weighted average of studies included in this analysis (‘Weighted average: Updated’), and the results of the model two (‘Modelled: Updated‘). Studies added in the 2024 review are shown in red and studies excluded from the meta-regression are shown in grey. Fowler 2015 and Mugerwa 2014 were excluded as they were not peer-reviewed studies, all other studies that were excluded had women who initiated AZT at 28 weeks rather than 14 weeks.

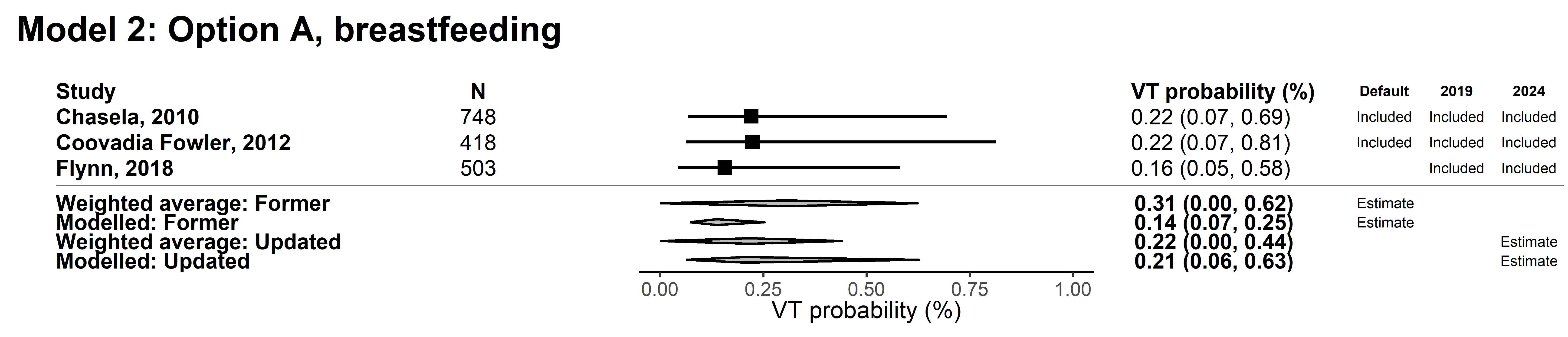

**Figure 3.2.11 Pooled estimates of breastfeeding VT among women receiving Option A.** The following pooled estimates are presented: the weighted average of studies included in the former VT probabilities (‘Weighted average: former VT’), estimates from model two fit to the pre-2024 systematic review studies (‘Modelled: former VT’), the weighted average of studies included in this analysis (‘Weighted average: Updated’), and the results of the model two (‘Modelled: Updated‘).

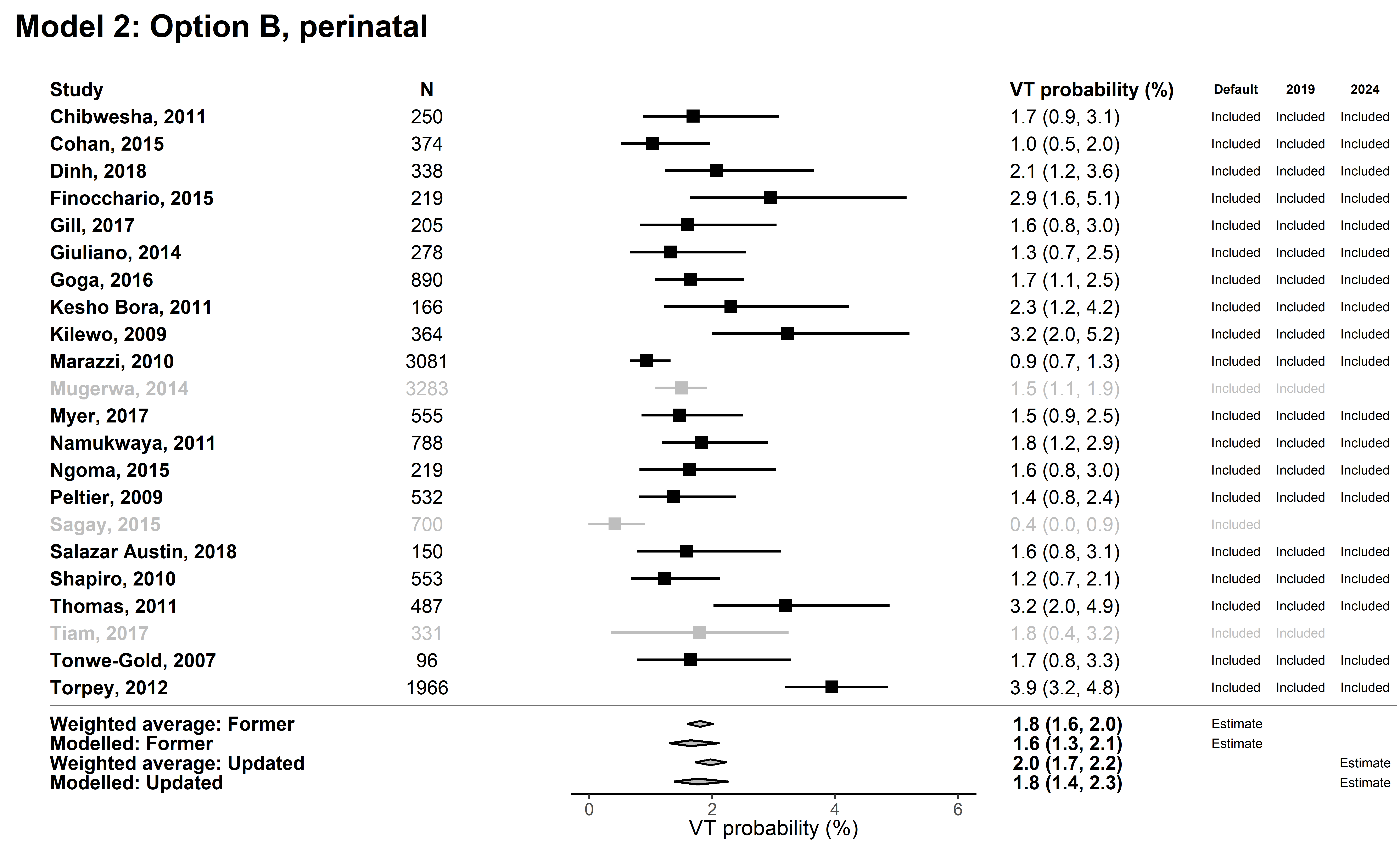

**Figure 3.2.12 Pooled estimates of perinatal VT among women receiving Option B.** The following pooled estimates are presented: the weighted average of studies included in the former VT probabilities (‘Weighted average: former VT’), estimates from model two fit to the pre-2024 systematic review studies (‘Modelled: former VT’), the weighted average of studies included in this analysis (‘Weighted average: Updated’), and the results of the model two (‘Modelled: Updated‘). Studies excluded from the meta-regression are shown in grey. Mugerwa 2014 and Tiam 2017 were excluded as they were not peer-reviewed studies. Sagay 2015 was excluded as we could not determine the timing of the HIV test.

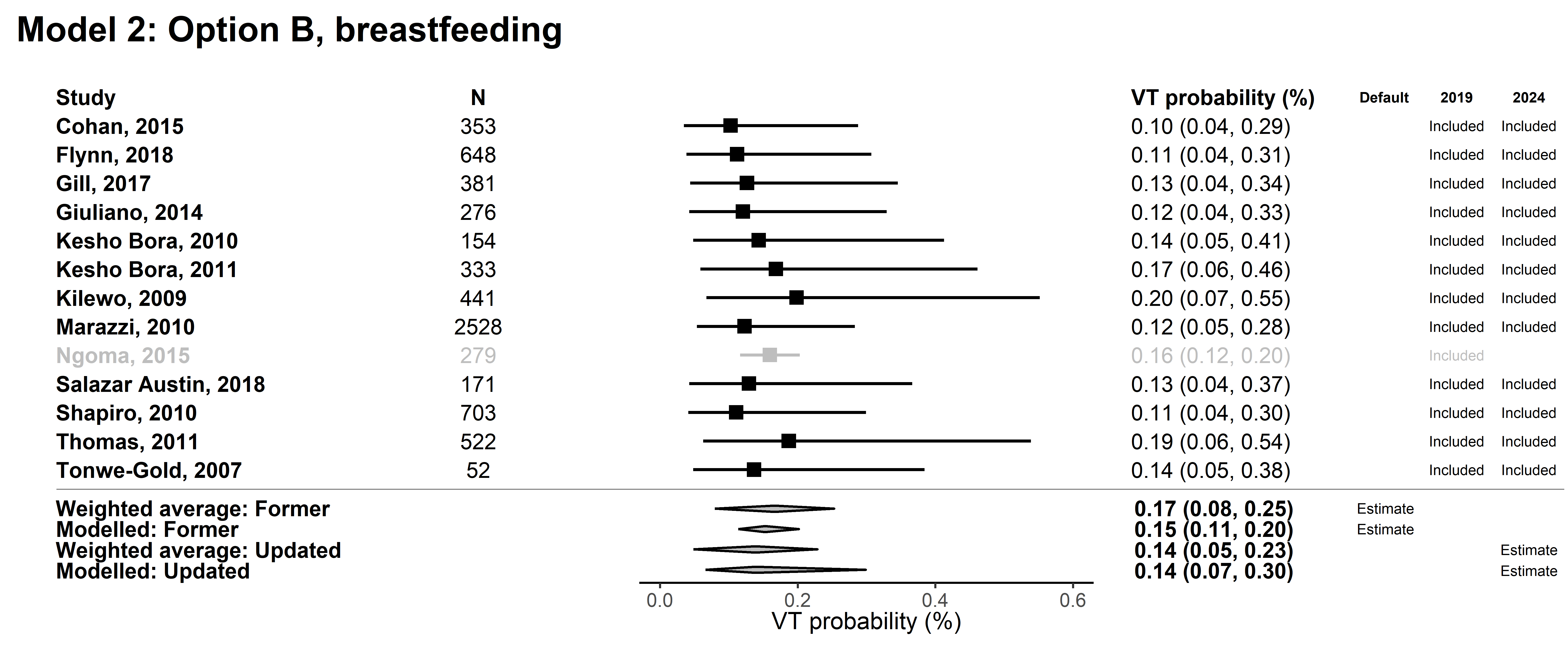

**Figure 3.2.13 Pooled estimates of breastfeeding VT among women receiving Option B.** The following pooled estimates are presented: the weighted average of studies included in the former VT probabilities (‘Weighted average: former VT’), estimates from model two fit to the pre-2024 systematic review studies (‘Modelled: former VT’), the weighted average of studies included in this analysis (‘Weighted average: Updated’), and the results of the model two (‘Modelled: Updated‘). Studies excluded from the meta-regression are shown in grey. Ngoma 2015 was excluded as it did not restrict to women with CD4 >350.

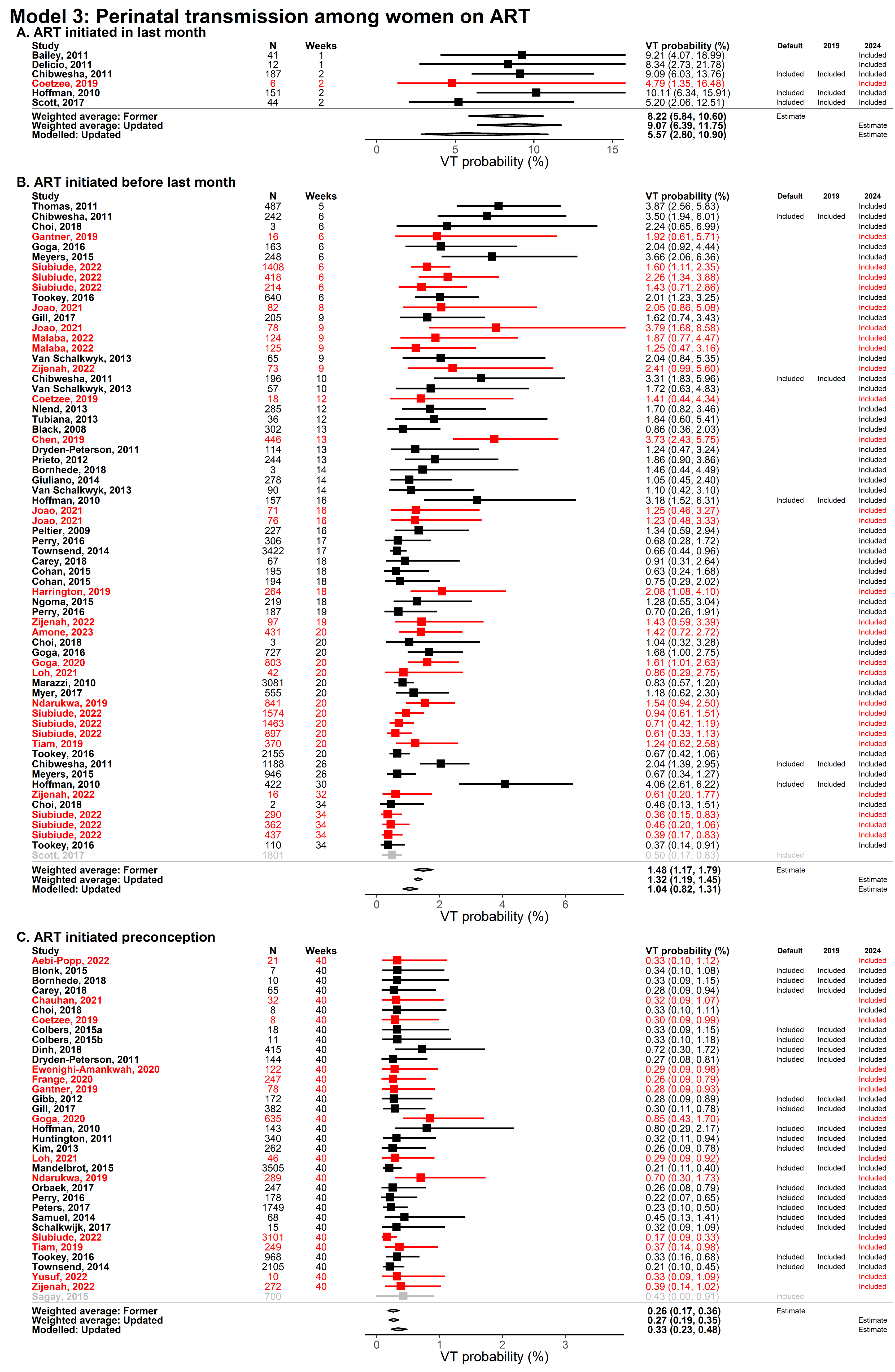

**Figure 3.2.14 Pooled estimates of perinatal VT among women receiving ART.** The following pooled estimates are presented: the weighted average of studies included in the former VT probabilities (‘Weighted average: former VT’), the weighted average of studies included in this analysis (‘Weighted average: Updated’), and the results of the model three (‘Modelled: Updated‘). Studies added in the 2024 review are shown in red and studies excluded from the meta-regression are shown in grey. Scott 2017 was excluded as we could not determine whether ART initiation occurred before or during pregnancy and Sagay 2015 was excluded as we could not determine paediatric test timing.

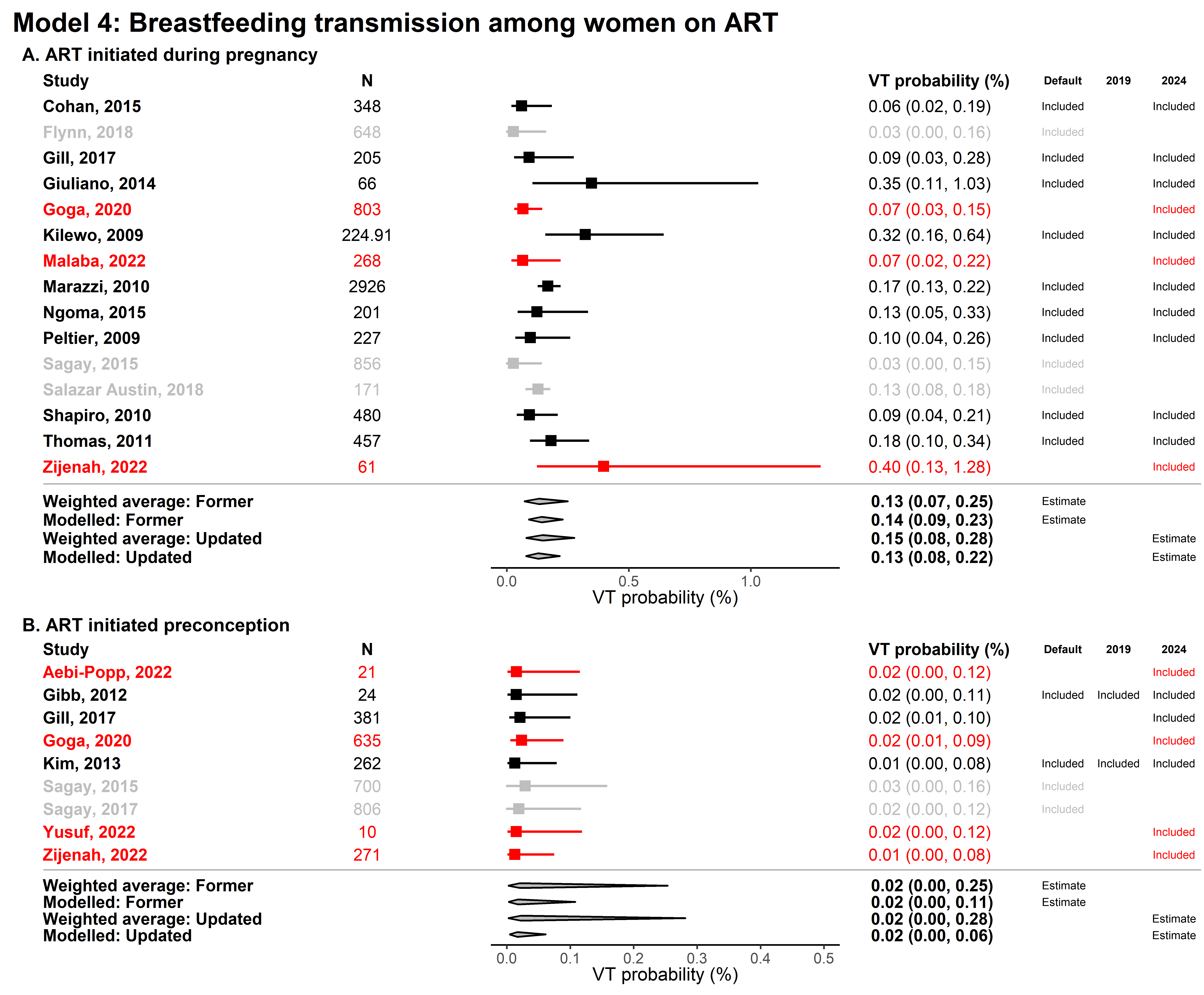

**Figure 3.2.15 Pooled estimates of breastfeeding VT among women receiving ART.** The following pooled estimates are presented: the weighted average of studies included in the former VT probabilities (‘Weighted average: former VT’), estimates from model four fit to the pre-2024 systematic review studies (‘Modelled: former VT’), the weighted average of studies included in this analysis (‘Weighted average: Updated’), and the results of the model four (‘Modelled: Updated‘).Studies added in the 2024 review are shown in red and studies excluded from the meta-regression are shown in grey. Flynn 2018, Sagay 2017, and Salazar Austin 2018 were excluded as we could not determine whether ART initiation occurred before or during pregnancy and Sagay 2015 was excluded as we could not determine paediatric test timing.

### 4. Sensitivity analyses on meta-regression model assumptions

#### 4.1. Model one: VT probability from women not receiving PVT

Model one included data from 16 studies that reported data about transmissions among women who did not receive PVT stratified by CD4 count. This including two new studies published since the 2018 review (Supplementary material 3.1). For the continuous CD4 covariate in model 1, we summarized the CD4 distribution among the study population by the approximate midpoint CD4. Here we assess (1) the sensitivity of model estimates to how the CD4 midpoint was defined from each study and (2) the sensitivity of estimates at different CD4 midpoints.

##### 4.1.1. Sensitivity to CD4 midpoint calculation

Data included in model one reported CD4 among the study population in one of two formats: (1) women with CD4 in a given range (e.g. 200 – 350 mm^3^) or (2) median CD4 of women who did not receive PVT care at baseline. If a study reported CD4 as a range, we took the median of the upper and lower bound (e.g. a range of 200-350 was extracted as 275). If a study reported CD4 using the second method (median CD4), the median was extracted.

Studies that stratified participates by CD4 range categories often reported vertical transmission probability data stratified by CD4 range, meaning that one study often reported multiple observations. Among studies that reported perinatal transmission among women not receiving PVT, four studies reported transmission by CD4 range. Three of the four studies reported transmission among three CD4 ranges. Nine studies reported perinatal transmission among women not on PVT that were not stratified by CD4 range, but did report a median CD4 among the study population. Among studies that reported breastfeeding transmission among women not receiving PVT, two studies reported transmission by CD4 range. One of the two studies reported transmission among multiple CD4 ranges. Four studies reported breastfeeding transmission among women not on PVT that were not stratified by CD4 range, but did report a median CD4 among the study population. Studies used in model one and method of CD4 reporting are listed in Table 4.1.1.1.

**Table 4.1.1.1.** Studies used in model one and method of CD4 reporting

| Study | Location | Study years | Transmission type | CD4 | VT | N | Method of CD4 reporting |
| --- | --- | --- | --- | --- | --- | --- | --- |
| Hoffman, 2010 | South Africa | 2004 - 2008 | Perinatal | 161 | 0.17 | 23 | Median |
| Coetzee, 2019 | South Africa | 2010 - 2010 | Perinatal | 272 | 0.13 | 15 | Median |
| Goga, 2016 | South Africa | 2011 - 2013 | Perinatal | 372 | 0.13 | 63 | Median |
| Olana, 2016 | Ethiopia | 2006 - 2014 | Perinatal | 415 | 0.13 | 102 | Median |
| Petra Study Team, 2002 | Tanzania, South Africa, Uganda | 2000 - 2000 | Perinatal | 435 | 0.13 | 303 | Median |
| Habib, 2021 | Iran | 2015 - 2017 | Perinatal | 475 | 0.20 | 20 | Median |
| Dabis, 1999 | Ivory Coast, Burkina Faso | 1995 - 1998 | Perinatal | 535 | 0.29 | 145 | Median |
| Connor, 1994 | USA and France | 1991 - 1993 | Perinatal | 538 | 0.22 | 183 | Median |
| Wiktor, 1999 | Ivory Coast | 1996 - 1998 | Perinatal | 548 | 0.22 | 119 | Median |
| Marinda, 2011 | Zimbabwe | 1997 - 2000 | Perinatal | 99.5 | 0.36 | 496 | CD4 range |
| Mayaux, 1995 | France | 1986 - 1994 | Perinatal | 99.5 | 0.43 | 65 | CD4 range |
| Shaffer, 1999 | Thailand | 1996 - 1997 | Perinatal | 99.5 | 0.38 | 24 | CD4 range |
| Marinda, 2011 | Zimbabwe | 1997 - 2000 | Perinatal | 274.5 | 0.24 | 826 | CD4 range |
| Mayaux, 1995 | France | 1986 - 1994 | Perinatal | 300 | 0.26 | 57 | CD4 range |
| Shaffer, 1999 | Thailand | 1996 - 1997 | Perinatal | 349.5 | 0.18 | 104 | CD4 range |
| Iliff, 2005 | Zimbabwe | 1997 - 2000 | Perinatal | 404 | 0.21 | 4367 | CD4 range |
| Marinda, 2011 | Zimbabwe | 1997 - 2000 | Perinatal | 424.5 | 0.20 | 1963 | CD4 range |
| Mayaux, 1995 | France | 1986 - 1994 | Perinatal | 600 | 0.17 | 192 | CD4 range |
| Shaffer, 1999 | Thailand | 1996 - 1997 | Perinatal | 600 | 0.13 | 67 | CD4 range |
| Petra Study Team, 2002 | Tanzania, South Africa, Uganda | 1996 - 2000 | BF (monthly) | 435 | 0.01 | 303 | Median |
| Chasela, 2010 | Malawi | 2004 - 2010 | BF (monthly) | 442 | 0.01 | 668 | Median |
| Dabis, 1999 | Ivory Coast, Burkina Faso | 1995 - 1998 | BF (monthly) | 535 | 0.01 | 113 | Median |
| Wiktor, 1999 | Ivory Coast | 1996 - 1998 | BF (monthly) | 548 | 0.02 | 115 | Median |
| Kuhn, 2010 | Zambia |  | BF (monthly) | 174.5 | 0.01 | 524 | CD4 range |
| Coovadia Fowler, 2012 | South Africa, Tanzania, Uganda, Zimbabwe | 2008 - 2011 | BF (monthly) | 424.5 | 0.01 | 434 | CD4 range |
| Kuhn, 2010 | Zambia |  | BF (monthly) | 424.5 | 0.00 | 469 | CD4 range |

As these two formats (CD4 range and median CD4) are not directly comparable, we conducted a subgroup analysis to assess the sensitivity of model one results to the difference in method of CD4 reporting. The subgroups we analysed were (1) studies that reported a CD4 range (“CD4 range” model) and (2) studies that reported a CD4 median (“median CD4” model). We re-fit model one to these different subgroups using Equation 4.1. Model one’s fit to all data as presented in the main text is also copied below (“all data” model).

$$\mathrm{logit}\left( {VT}_{BF, CD4, S, Obs} \right)=\beta_{0, BF=0}+\beta_{1, BF=1}+\beta_{2}*{CD4}_{Midpoint}+\beta_{3,BF=1}*{CD4}_{Midpoint}+\mu_{S}+\mu_{Obs}$$

Equation 4.1

The following description of Equation 4.1 is copied from the main text: “*Model one included fixed effects for perinatal (*$\beta_{0}$*) and monthly breastfeeding VT (*$\beta_{1}$*), CD4 midpoint of the study population (*$\beta_{2},$ *per 100 mm^3^, centred at 500 mm^3^), and the interaction between CD4 midpoint and breastfeeding timing (*$\beta_{3}$*). CD4 midpoint was extracted as the median CD4 of WLHIV not receiving ARVs or the midpoint of relevant CD4 range if studies reported VT by CD4 categories… Random effects were included for study and observation (*$\mu_{0}$ *and* $\mu_{1}$*, respectively).*”

**Table 4.1.1.2.** Sensitivity analysis is of model 1 results to CD4 midpoint assumption for data used in model one

| Covariate | Estimates of the “all data” model (logit)  (n = 12,021) | | Estimates of the  “CD4 range” model  (logit)  (n = 9642) | | Estimates of the “median CD4” model  (logit)  (n = 2379) | |
| --- | --- | --- | --- | --- | --- | --- |
| Intercept | -1.61 | (-1.81, -1.41) | -1.57 | (-1.65, -1.48) | -1.47 | (-1.63, -1.30) |
| CD4 midpoint  (per 100 cells increase, centered on CD4 = 500 mm^3^) | -0.23 | (-0.28, -0.17) | -0.24 | (-0.29, -0.18) | 0.33 | (0.10, 0.56) |
| Perinatal transmission  (Reference) | 0.00 | (Reference) | 0.00 | (Reference) | 0.00 | (Reference) |
| Breastfeeding transmission | -3.22 | (-3.84, -2.60) | -4.05 | (-5.33, -2.77) | -2.88 | (-3.58, -2.19) |
| Interaction between CD4 midpoint and breastfeeding transmission | 0.17 | (-0.21, 0.54) | -0.08 | (-0.59, 0.43) | 0.20 | (-0.92, 1.32) |

The covariates of the “all data” model one and “CD4 range” model one fit had overlapping confidence intervals (Table 4.1.1.2). The “all data” model one and “median CD4” model one fit were significantly different from each other. While the “all data” model one had a negative estimate for the effect of CD4 midpoint on VT probability (-0.2 (-0.3 - -0.2)), the “median CD4” model had a positive estimate for the effect of CD4 midpoint on VT probability (0.4 (0.1-0.7)).

The difference in effect of CD4 midpoint on VT probability among women not receiving PVT was driven by three studies that reported both high CD4 medians and transmission rates (Dabis 1999, Connor 1994, and Wiktor 1999, Table 4.1.1.1). Excluding these three studies resulted in a negative CD4 midpoint fixed effect. These studies collected data in the early stage of the HIV epidemic (1991 to 1998 across the three studies), reported high transmission rates and CD4 medians. This may may reflect high incidence among women of childbearing age. In acute HIV infection, both CD4 and viral load are high, and so women with high CD4 can have higher risk of vertical transmission than women no longer in the acute infection stage.^4^ Dabis 1999 describes data from the placebo group of the DITRAME trial which had high average CD4 and viral load; among women who transmitted the mean CD4 was 358 and the mean viral load was approximately 55,000.^5^ Similar data was not available Connor 1994 and Wiktor 1999.

The observations from these three studies (Dabis, Connor, and Wiktor) represent 31% of the available data in the “median CD4” model and are the only observations with CD4 midpoints above 500. The results of the “median CD4” model are confounded by studies that reflect an earlier HIV epidemic, where transmission probability may be high among high CD4 values due to high incidence rates. When all data is used, the effect of these studies is diluted as there is more data about transmission among women with high CD4 values. The meta-regression framework then imposes larger study and observation level random effects on observations from Dabis 1999, Connor 1994, and Wiktor 1999. Although subgroup analysis shows that model one results are sensitive to the format of CD4 midpoint reporting, the difference in transmission may be confounded by other study characteristics (specifically study year and epidemic stage) rather than due to reporting a CD4 median itself.

##### 4.1.2 Sensitivity to CD4 midpoint used to produce estimates for Spectrum

For women not receiving PVT, Spectrum-AIM stratifies transmission probabilities by CD4 range. These are CD4 <200, 200-350, and >350. To produce model based estimates of VT probabilities compatible with Spectrum-AIM’s stratification, we used CD4 midpoints of 100, 275, and 500 (“Main text value” in Table 4.1.2). We assessed the sensitivity of our estimates to this choice by considering alternate uses of model one that could be used to approximate the Spectrum-AIM CD4 categories. Alternate approaches included: the mean of all transmission probabilities for CD4 midpoints within the Spectrum-AIM ranges (“Mean across range” in Table 4.1.2), the lowest CD4 of the Spectrum-AIM ranges (“Lowest CD4” in Table 4.1.2), and the highest CD4 of the Spectrum-AIM range (“Highest CD4” in Table 4.1.2). For the >350 category, the highest CD4 we considered was 650 mm^3^.

**Table 4.1.2** Sensitivity of model one VT estimates to CD4 midpoint chosen to produce Spectrum-AIM compatible estimates

|  | CD4 range | Main text value | | Mean across range | | Lowest CD4 | | Highest CD4 | |
| --- | --- | --- | --- | --- | --- | --- | --- | --- | --- |
| Perinatal | <200 | 33.4 | (27.8, 39.1) | 33.1 | (26.0, 41.8) | 38.8 | (32.0, 45.7) | 28.6 | (24.0, 32.7) |
|  | 200-350 | 25.1 | (21.4, 28.9) | 25.1 | (20.0, 30.8) | 28.6 | (24.0, 32.6) | 22.1 | (18.8, 25.4) |
|  | >350 | 16.6 | (13.9, 19.7) | 16.8 | (11.4, 23.2) | 22.1 | (18.8, 25.2) | 12.5 | (9.9, 15.7) |
| BF (monthly) | <200 | 1.0 | (0.3, 3.3) | 1.1 | (0.4 3.5) | 1.2 | (0.3, 4.8) | 1.0 | (0.5, 2.2) |
|  | 200-350 | 0.9 | (0.5, 1.7) | 0.9 | (0.5, 1.8) | 1.0 | (0.5, 2.2) | 0.9 | (0.5, 1.5) |
|  | >350 | 0.8 | (0.4, 1.4) | 0.8 | (0.4, 1.5) | 0.9 | (0.5, 1.5) | 0.7 | (0.3, 1.9) |

Perinatal transmission rates were within a -4.7% to 5.6% absolute difference range of the main text values estimated using the CD4 midpoints of 100, 275, and 500. Breastfeeding transmission rates were within a -0.1% to 0.1% absolute difference range of the main text values. The largest difference was among perinatal transmission with CD4 >350 (Table 4.1.2). The midpoint of 500 produced a transmission probability of 16.7% (13.8–20.0%). The highest CD4 in that range (CD4 = 650) had a transmission rate of 12.5% (9.9%–15.7%) while the lowest CD4 in that range (CD4 = 351) had a transmission rate of 22.1% (18.8–25.2%). If CD4 testing data shows that most women have high CD4 values (> 500), the VT probability used in Spectrum may be overestimating VT. However, given high PVT coverage, most women who are exposed to the VT probabilities for women not receiving PVT will have been those who were not retained on care. Treatment interruption is associated with rapid decline in CD4 count and increase in viral load after interruption, indicating that VT probability in this group is unlikely to be underestimated.^6^

#### 4.2. Model three: perinatal transmission probability from women receiving ART by timing of initiation

Model three included data from 57 studies (Supplementary material 3.3). Here we assess the sensitivity of model three’s fit to the calculation used to extract a time on ART midpoint from each study.

##### 4.2.1 Sensitivity to time on ART data extraction

Publications reported time on ART before delivery in one of four ways: (1) median or mean number of weeks on ART before delivery, (2) median or mean gestational week of ART initiation, (3) a range of gestational weeks when women initiated ART (either reported as a trimester or through exclusion criteria), and (4) women who were on ART pre-conception.

To maximize the number of studies used, we used all reporting types and attributed a number of “weeks on ART midpoint” for studies where women used lifelong ART during pregnancy. The minimum value accepted was 0.5, indicating that the women initiated ART one week before delivery and the maximum value accepted was 40, indicating that the women initiated ART preconception. We preferentially extracted median (or mean) weeks on ART before delivery or median (mean) gestational week of ART initiation. As described in Supplementary Material 2, for countries that reported time on ART using the third method (a range of weeks), we contacted authors to request a median number of weeks on ART before delivery for the cohort. For the remaining studies where median time on ART was not available and the only information was a range of weeks when women initiated ART, we used the median of this range to determine the weeks on ART midpoint. The method used to derive the weeks on ART is shown for each study used in model three in Table 4.2.1.1.

**Table 4.2.1.1.** Data used to estimate effect of time on ART on VT estimates

| Study | Location | Study years | Weeks on ART midpoint | VT events | N | Method to derive number of weeks on ART^A^ |
| --- | --- | --- | --- | --- | --- | --- |
| Bailey, 2011 | Europe | 2000 - 2009 | 1 | 5 | 41 | 3 |
| Delicio, 2011 | Brazil | 2000 - 2009 | 1 | 2 | 12 | 3 |
| Chibwesha, 2011 | Zambia | 2007 - 2010 | 2 | 17 | 187 | 3 |
| Coetzee, 2019^B^ | South Africa | 2010 - 2010 | 2 | 0 | 6 | 2 |
| Hoffman, 2010 | South Africa | 2004 - 2008 | 2 | 14 | 151 | 3 |
| Scott, 2017 | USA | 2002 - 2009 | 2 | 2 | 44 | 3 |
| Tubiana, 2013 | France | 2007 - 2010 | 3.2 | 1 | 36 | 1 |
| Thomas, 2011 | Kenya | 2003 - 2009 | 5 | 20 | 487 | 3 |
| Choi, 2018 | Korea | 2005 - 2017 | 6 | 0 | 3 | 3 |
| Gantner, 2019 | France | 2008 - 2014 | 6 | 0 | 16 | 3 |
| Goga, 2016 | South Africa | 2011 - 2013 | 6 | 2 | 163 | 3 |
| Meyers, 2015 | China | 2010 - 2012 | 6 | 11 | 248 | 3 |
| Siubiude, 2017 | France | 2000 - 2023 | 6 | 36 | 2152 | 3 |
| Tookey, 2016 | UK | 2003 - 2013 | 6 | 13 | 640 | 3 |
| Malaba, 2018 ^B^ | South Africa And Uganda | 2022 - 2018 | 9 | 3 | 135 | 2 |
| Malaba, 2018 ^B^ | South Africa And Uganda | 2022 - 2018 | 9 | 0 | 133 | 2 |
| Van Schalkwyk, 2013^B^ | South Africa | 2008 - 2010 | 9 | 2 | 65 | 2 |
| Zijenah, 2022 ^B^ | Zimbabwe | 2017 - 2018 | 9 | 2 | 73 | 2 |
| Gill, 2017 | Rwanda | 2013 - 2014 | 9.6 | 3 | 205 | 1 |
| Van Schalkwyk, 2013^B^ | South Africa | 2008 - 2010 | 10 | 1 | 57 | 2 |
| Joao, 2021 | Argentina, Brazil, South Africa, Tanzania, Thailand, USA | 2013 - 2018 | 11.5 | 7 | 393 | 3 |
| Nlend, 2013 | Cameroon | 2012 - 2008 | 12 | 5 | 285 | 1 |
| Black, 2008 | South Africa | 2004 - 2007 | 13 | 1 | 302 | 2 |
| Chen, 2019 | China | 2007 - 2015 | 13 | 19 | 446 | 3 |
| Prieto, 2012 | Spain | 2000 - 2007 | 13 | 5 | 244 | 3 |
| Dryden-Peterson, 2011 | Botswana | 2009 - 2010 | 13.1 | 1 | 114 | 2 |
| Bornhede, 2018 | Sweden | 2014 - 2017 | 14 | 0 | 3 | 3 |
| Giuliano, 2014 | Malawi | 2008 - 2009 | 14 | 2 | 278 | 2 |
| Van Schalkwyk, 2013^B^ | South Africa | 2008 - 2010 | 14.6 | 0 | 90 | 2 |
| Townsend, 2014 | UK and Ireland | 2007 - 2011 | 17.3 | 21 | 3422 | 2 |
| Perry, 2016 | UK | 2007 - 2012 | 17.9 | 1 | 306 | 2 |
| Carey, 2018 | UK | 2008 - 2014 | 18 | 0 | 67 | 2 |
| Harrington, 2019 | Malawi | 2015 - 2016 | 18 | 7 | 264 | 2 |
| Ngoma, 2015 | Zambia | 2008 - 2009 | 18 | 3 | 219 | 3 |
| Cohan, 2015 | Uganda | 2009 - 2013 | 18.8 | 1 | 374 | 2 |
| Perry, 2016 | UK | 2007 - 2012 | 19.6 | 1 | 187 | 2 |
| Zijenah, 2022^B^ | Zimbabwe | 2017 - 2018 | 19 | 2 | 97 | 2 |
| Amone, 2016 | Uganda | 2017 - 2023 | 20 | 7 | 431 | 3 |
| Choi, 2018 | Korea | 2005 - 2017 | 20 | 0 | 3 | 3 |
| Goga, 2016 | South Africa | 2011 - 2013 | 20 | 14 | 727 | 3 |
| Loh, 2021 | Singapore | 2008 - 2015 | 20 | 0 | 42 | 3 |
| Myer, 2017 | South Africa | 2013 - 2014 | 20 | 7 | 555 | 2 |
| Ndarukwa, 2019 | Zimbabwe | 2014 - 2016 | 20 | 13 | 841 | 3 |
| Siubiude, 2017 | France | 2000 - 2023 | 20 | 31 | 4147 | 3 |
| Tiam, 2019 | Lesotho | 2014 - 2016 | 20 | 5 | 370 | 3 |
| Tookey, 2016 | UK | 2003 - 2013 | 20 | 13 | 2155 | 3 |
| Marazzi, 2010 | Malawi and Mozambique | 2005 - 2009 | 20.1 | 25 | 3081 | 2 |
| Chibwesha, 2011 | Zambia | 2007 - 2010 | 22 | 42 | 1626 | 3 |
| Hoffman, 2010 | South Africa | 2004 - 2008 | 22 | 28 | 579 | 3 |
| Van Schalkwyk, 2013^B^ | South Africa | 2008 - 2010 | 24 | 1 | 127 | 3 |
| Meyers, 2015 | China | 2010 - 2012 | 26 | 5 | 946 | 3 |
| Coetzee, 2019^B^ | South Africa | 2010 - 2010 | 28 | 0 | 18 | 2 |
| Zijenah, 2022^B^ | Zimbabwe | 2017 - 2018 | 32 | 0 | 16 | 2 |
| Choi, 2018 | Korea | 2005 - 2017 | 34 | 0 | 2 | 3 |
| Siubiude, 2017 | France | 2000 - 2023 | 34 | 6 | 1149 | 3 |
| Tookey, 2016 | UK | 2003 - 2013 | 34 | 0 | 110 | 3 |
| Aebi-Popp, 2022 | Switzerland | 2019 - 2021 | 40 | 0 | 17 | 4 |
| Blonk, 2015 | Europe | 2010 - 2014 | 40 | 0 | 7 | 4 |
| Bornhede, 2018 | Sweden | 2014 - 2017 | 40 | 0 | 10 | 4 |
| Carey, 2018 | UK | 2008 - 2014 | 40 | 0 | 65 | 4 |
| Chauhan, 2021 | India | 2016 - 2018 | 40 | 0 | 32 | 4 |
| Choi, 2018 | Korea | 2005 - 2017 | 40 | 0 | 8 | 4 |
| Coetzee, 2019^B^ | South Africa | 2010 - 2010 | 40 | 0 | 8 | 4 |
| Colbers, 2015a | Europe |  | 40 | 0 | 18 | 4 |
| Colbers, 2015 | Europe | 2009 - 2014 | 40 | 0 | 11 | 4 |
| Dinh, 2018 | Zimbabwe | 2013 - 2013 | 40 | 5 | 415 | 4 |
| Dryden-Peterson, 2011 | Botswana | 2009 - 2010 | 40 | 0 | 144 | 4 |
| Ewenighi-Amankwah, 2020 | Nigeria |  | 40 | 0 | 122 | 4 |
| Frange, 2020 | France | 2010 - 2018 | 40 | 0 | 247 | 4 |
| Gantner, 2019 | France | 2008 - 2014 | 40 | 0 | 78 | 4 |
| Gibb, 2012 | Uganda, Zimbabwe | 2003 - 2009 | 40 | 0 | 172 | 4 |
| Gill, 2017 | Rwanda | 2014 - 2016 | 40 | 1 | 382 | 4 |
| Goga, 2020 | South Africa | 2012 - 2014 | 40 | 7 | 635 | 4 |
| Hoffman, 2010 | South Africa | 2004 - 2008 | 40 | 1 | 143 | 4 |
| Huntington, 2011 | UK | 1996 - 2009 | 40 | 1 | 340 | 4 |
| Kim, 2013 | Malawi | 2009 - 2011 | 40 | 0 | 262 | 4 |
| Loh, 2021 | Singapore | 2008 - 2015 | 40 | 0 | 46 | 4 |
| Mandelbrot, 2015 | France | 2000 - 2011 | 40 | 6 | 3505 | 4 |
| Ndarukwa, 2019 | Zimbabwe | 2014 - 2016 | 40 | 4 | 289 | 4 |
| Orbaek, 2017 | Denmark | 2002 - 2014 | 40 | 0 | 247 | 4 |
| Perry, 2016 | UK | 2007 - 2012 | 40 | 0 | 178 | 4 |
| Peters, 2017 | UK | 2012 - 2014 | 40 | 3 | 1749 | 4 |
| Sagay, 2015 | Nigeria | 2010 - 2012 | 40 | 3 | 700 | 4 |
| Samuel, 2014 | UK | 2004 - 2010 | 40 | 1 | 68 | 4 |
| Schalkwijk, 2017 | Europe | - | 40 | 0 | 15 | 4 |
| Siubiude, 2017 | France | 2000 - 2023 | 40 | 9 | 6606 | 4 |
| Tiam, 2019 | Lesotho | 2014 - 2016 | 40 | 1 | 249 | 4 |
| Tookey, 2016 | UK | 2003 - 2013 | 40 | 4 | 968 | 4 |
| Townsend, 2014 | UK and Ireland | 2007 - 2011 | 40 | 4 | 2105 | 4 |
| Yusuf, 2022 | USA | - | 40 | 0 | 1 | 4 |
| Zijenah, 2022^B^ | Zimbabwe | 2017 - 2018 | 40 | 1 | 272 | 4 |

^A^ (1) Reported median or mean weeks on ART before delivery

(2) Reported median or mean gestational week of ART initiation

(3) Reported a range of gestational weeks during which women initiated ART

(4) Reported women who were on ART pre-conception

^B^ Median gestational weeks at ART initiation was not reported in text, however corresponding author provided this information upon request.

As these two formats are not directly comparable, we conducted a subgroup analysis to assess the sensitivity of model three results to the difference in method for extracting the “weeks on ART midpoint”. The subgroups we analysed were (1) studies that reported a median number of weeks on ART before delivery, studies that reported ART was initiated in the last four weeks of pregnancy, and transmission among women on ART preconception (methods one, two, and four above, “Median weeks reported” below) and (2) studies that reported a range of gestational weeks when women initiated ART or transmission among women on ART preconception (methods three and four above, “Range of weeks reported” below). We re-fit model three to these different subgroups using Equation 4.2. Model three’s fit to all data as presented in the main text is also copied below (“All data” model).

$$logit({VT}_{BF=0,Weeks, late, S,Obs})=\beta_{0}+\beta_{1}*T_{weeks}+\beta_{2}*late initation+\mu_{S}+\mu_{Obs}$$

Equation 4.2

The following description of Equation 4.2 is copied from the main text: “*Model three included a fixed effect (*$\beta_{1}$*) for weeks on ART before delivery (T_Weeks_, centred on ART initiated 20 weeks before delivery; specified as 40 weeks for women already on ART at conception), and a fixed effect for ART initiation less than four weeks before delivery (*$\beta_{2}$*, ‘late initiation’). The ‘late initiation’ effect accounted for increased risk of unsuppressed VL at delivery when ART is initiated late in pregnancy. T_Weeks_ was extracted as the median weeks on ART before delivery, where available. For studies that reported ART initiation as a range of gestational weeks, we extracted the range midpoint. We contacted corresponding authors of studies that reported a range of gestational weeks when ART was initiated to request the median gestational week ART was initiated... Random effects were included for study and observation (*$\mu_{0}$ *and* $\mu_{1}$*, respectively).”*

**Table 4.2.1.2.** Sensitivity analysis is of model three results to time on ART midpoint assumption

| Covariate | Estimates of the  “All data”  model  (logit)  (n = 45,220) | | Estimates of the  “Median weeks  reported” model  (logit)  (n = 34,831) | | Estimates of the “Range of weeks reported” model  (logit)  (n = 26,311) | |
| --- | --- | --- | --- | --- | --- | --- |
| Intercept | -4.55 | (-4.79, -4.32) | -4.83 | (-5.14, -4.54) | -4.38 | (-4.65, -4.11) |
| Weeks on ART before delivery  (centered on 20 weeks) | -0.06 | (-0.07, -0.04) | -0.05 | (-0.07, -0.03) | -0.06 | (-0.08, -0.05) |
| Late ART initiation (<4 weeks before delivery) | 0.68 | (-0.05, 1.45) | 1.51 | (0.77, 2.30) | 0.57 | (-0.19, 1.38) |

The “all data” model three and “range of weeks reported” model three fit were not significantly different from each other (Table 4.2.1.2). The “all data” model three and “median weeks reported” model three fit were significantly different from each other. The late ART initiation covariate was significantly positive in the “median weeks reported” model, whereas in the “all data” model it was positive, but not significant.

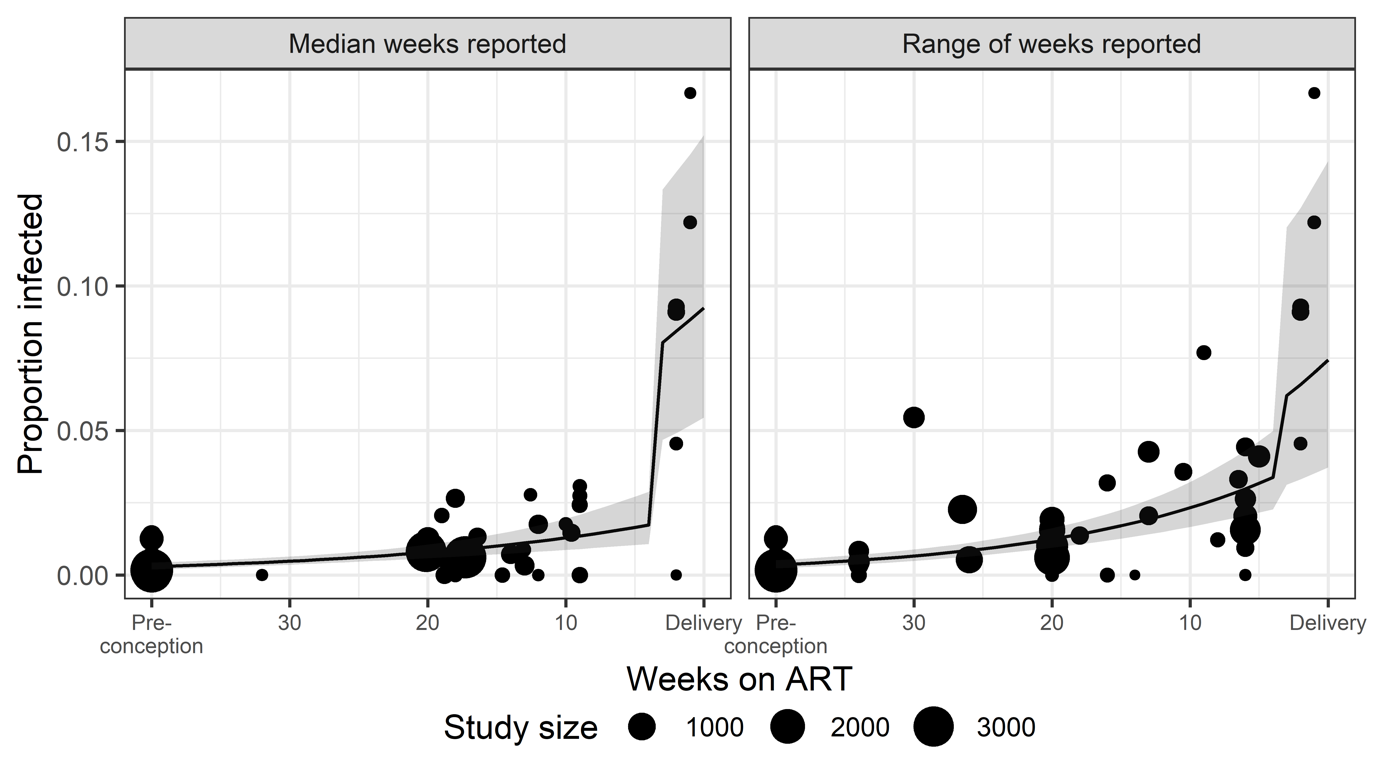

**Figure 4.2.1.** Effect of definition of weeks on ART midpoint on model three estimates of perinatal VT probability

The “all data” and “range of weeks reported” models included more studies where women initiated ART in the final ten weeks of pregnancy. These studies had VT probabilities ranging from 0% to approximately 7.5%, making it so that the studies with late ART initiation weren’t significantly higher than what was captured in the “weeks on ART” covariate.

##### 4.2.2 Sensitivity to assumption of 40 gestational weeks at delivery

For studies where we inferred the time on ART from the reported timing of ART initiation, we assumed that the average gestational age at delivery was 40 weeks (“full-term”). As not all women will deliver at full-term, we assessed the sensitivity of our results to this assumption. Margulis et al 2023^7^ published estimates of gestational age at birth from U.S. birth data from the Center for Disease Control. Margulis et al reported a median gestational age at birth of 39 weeks. Similar data at the required granularity wasn’t available for other regions, however, estimates of gestational age at birth from a cohort of South African women living with HIV were slightly lower, but similar (mean: 37.6 (range of 26-42)).^8^

For each study where we inferred the number of weeks on ART given delivery at full-term, we re-estimated the number of weeks on ART from the probability distribution of gestational age at delivery published by Margulis et al. Using the new dataset, we re-fit model 3 (Table 4.2.2.1). Model 3 fit to data that assumed full-term gestation and those that used the probability distributions published by Margulis et al produced similar results, with overlapping confidence intervals.

**Table 4.2.2.1.** Sensitivity analysis is of model three results to gestational age at delivery assumption

| Covariate | Estimates of the full-term model  (logit) | | Estimates of the weighted gestational age model  (logit) | |
| --- | --- | --- | --- | --- |
| Intercept | -4.55 | (-4.79, -4.32) | -4.63 | (-4.87, -4.40) |
| Weeks on ART before delivery  (centered on 20 weeks) | -0.06 | (-0.07, -0.04) | -0.05 | (-0.07, -0.04) |
| Late ART initiation (<4 weeks before delivery) | 0.68 | (-0.05, 1.45) | 0.75 | (0.11, 1.42) |

The estimates of VT probability for model three fit to the weighted gestational age model were lower for women who initiate ART <4 weeks before delivery (Table 4.2.2.2). This was despite a higher odds ratio for the coefficient for late ART initiation (<4 weeks before delivery) in the weighted gestational age model was significantly greater than one (OR 2.12 (1.12-4.14)). Given that only 1% of WLHIV globally fall into this transmission category, Spectrum-AIM’s estimates of vertical transmission aren’t sensitive to these results. More work should be done to characterize gestational age at delivery among WLHIV in diverse global settings to improve these estimates.

**Table 4.2.2.2.** Sensitivity of model three estimates of VT probability to gestational age at delivery assumption

| VT probability | Estimates of the full-term model  (logit) | | Estimates of the weighted gestational age model  (logit) | |
| --- | --- | --- | --- | --- |
| Option B+, on ART <4 weeks | 5.6 | (2.8 - 10.9) | 4.8 | (2.9 – 9.4) |
| Option B+, on ART 5-39 weeks | 1.0 | (0.8 - 1.3) | 1.0 | (0.8 - 1.2) |
| Option B+, on ART pre-conception | 0.33 | (0.23 - 0.48) | 0.36 | (0.19 - 0.55) |

#### 4.3 Geographic region as a confounder of ART class’s effect on VT probability

Because 9/12 studies that reported perinatal transmission among women receiving an INSTI-based regimen occurred in high-income countries, we assessed geographic region as a confounder of ART class’s effect on VT probability. To do so, we fit a modified version of model three, with fixed effects on ART class and geographic region. Geographic region was coded as: Sub-Saharan Africa (SSA, reference), non SSA, or multiple regions.

**Table 4.3.** Geographic region as a potential confounder for ART class’s effect on VT probability

|  | No geographic region fixed effect | | | | Geographic region fixed effect | | |
| --- | --- | --- | --- | --- | --- | --- | --- |
| Covariate | Estimate  (logit) | | 95% confidence interval | | Estimate (logit) | | 95% confidence interval |
| Intercept | -4.45 | (-4.71, -4.19) | | -4.31 | | (-4.57, -4.05) | |
| Weeks on ART before delivery  (centered on 20 weeks) | -0.06 | (-0.07, -0.04) | | -0.06 | | (-0.07, -0.04) | |
| Late ART initiation  (<4 weeks before delivery) | 0.72 | (-0.05, 1.49) | | 1.05 | | (0.31, 1.79) | |
| ART class |  |  | |  | |  | |
| NNRTI (reference) | 0.00 | (Reference) | | 0.00 | | (Reference) | |
| INSTI | -1.01 | (-1.95, -0.07) | | -0.74 | | (-1.75, 0.26) | |
| PI | -0.12 | (-0.56, 0.32) | | 0.07 | | (-0.37, 0.51) | |
| Miscellaneous regimens | -0.04 | (-0.67, 0.59) | | 0.22 | | (-0.29, 0.72) | |
| Geographic region | Not included | | |  | |  | |
| SSA |  |  |  | 0.00 | | (Reference) | |
| Non-SSA |  |  |  | -0.58 | | (-0.95, -0.21) | |
| Multiple regions |  |  |  | -0.18 | | (-1.07, 0.72) | |

In the model that included geographic region, the non-SSA region had the lowest VT probability when a NNRTI-based regimen was initiated 20 weeks before delivery, although the regions didn’t differ significantly (Table 4.3). While in the model without geographic region specified INSTI-based regimens had significantly lower VT when ART was started 20 weeks before delivery, including geographic region in the mode made this effect not significant. This suggests that the effects of ART regimen class on perinatal VT are confounded by the study geographic region.

### 5. Implications of estimated VT probabilities for Spectrum-AIM’s estimates of paediatric HIV infections

We used our estimates of VT probability in Spectrum-AIM model to assess the change in the number of perinatal and breastfeeding infections compared to those calculated using the former VT probabilities. For women not receiving any treatment, we used model one to estimated VT probability at the following CD4 midpoints: 100, 275, and 500. These were used to align with the Spectrum-AIM CD4 categories of <200, 200-350, and >350. For women receiving lifelong ART, used model three to estimate perinatal VT for weeks on ART of 2, 20, and 40 weeks to represent the Spectrum-AIM’s perinatal transmission categories of on ART of <4 weeks, for 4-39 weeks before delivery, and preconception. For breastfeeding transmission probabilities, we used model four to estimate a monthly breastfeeding VT probability for women who initiated ART preconception and women who initiated ART during pregnancy. Probabilities of VT for women who seroconverted during pregnancy or breastfeeding and women who received short-course PVT were estimated using model two. The values used in this analysis are listed in Table 1 and Table 2 of the main text.

Using the 2024 published publicly available Spectrum-AIM files for Malawi, Rwanda, Democratic Republic of the Congo, and Burkina Faso, we calculated the percent change in the number of perinatal infections (Equation 5.1), breastfeeding infections (Equation 5.2), and total paediatric infections (Equation 5.3) in the years 2000, 2010, 2015, and 2023. These countries were chosen as they represent a country in Southern, Eastern, Central, and Western Africa respectively, and these years represent a variety of PVT strategies and coverages. These results do not necessarily reflect the potential results of applying these changes to other or all countries.

$${Percent change}_{Perinatal}= \frac{({Infections}_{Perinatal,MR}- {Infections}_{Perinatal,Former})}{{Infections}_{Perinatal,Former}}*100$$

Equation 5.1

In Equation 5.1, ${Infections}_{Perinatal,MR}$ represented the number of perinatal infections that resulted from using the VT probabilities estimated in the meta-regression analysis, ${Infections}_{Perinatal,Former}$ represented the number of perinatal infections that resulted from using the former Spectrum-AIM VT probabilities, and ${Percent change}_{Perinatal}$ represented the percent change in perinatal infections.

$${Percent change}_{BF}= \frac{({Infections}_{BF,MR}- {Infections}_{BF,Default})}{{Infections}_{BF,Former}}*100$$

Equation 5.2

In Equation 5.2, ${Infections}_{BF,MR}$ represented the number of breastfeeding infections that resulted from using the VT probabilities estimated in the meta-regression analysis, ${Infections}_{BF,Former}$ represented the number of breastfeeding infections that resulted from using the former Spectrum-AIM VT probabilities, and ${Percent change}_{BF}$ represented the percent change in breastfeeding infections.

$${Percent change}_{Total}= \frac{({Infections}_{Total,MR}- {Infections}_{Total,Former})}{{Infections}_{Total,Former}}*100$$

Equation 5.3

In Equation 5.3, ${Infections}_{Total,MR}$ represented the number of vertical infections that resulted from using the VT probabilities estimated in the meta-regression analysis, ${Infections}_{Total,Former}$ represented the number of vertical infections that resulted from using the former Spectrum-AIM VT probabilities, and ${Percent change}_{Total}$ represented the percent change in vertical infections.

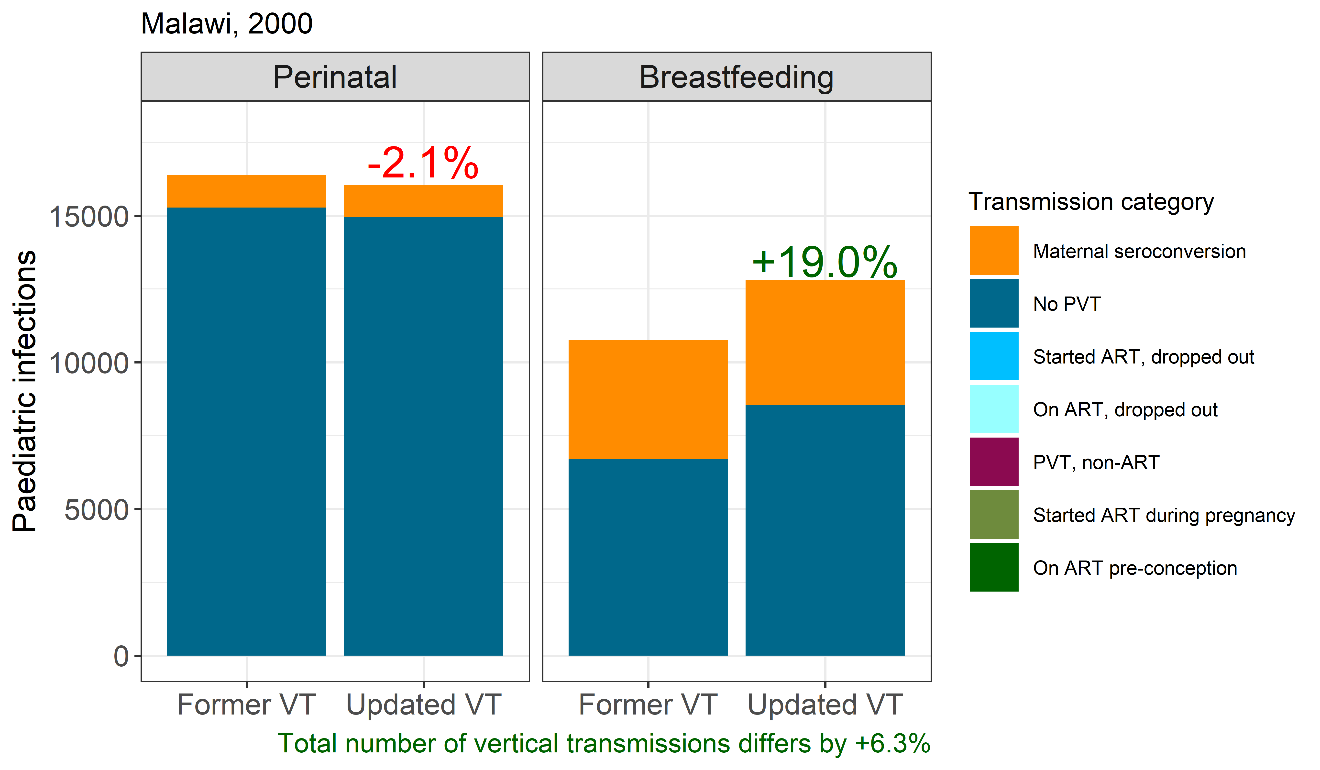

**Figure 5.1.** Change in vertical infections due to estimated vertical transmission probabilities by infection timing, Malawi 2000

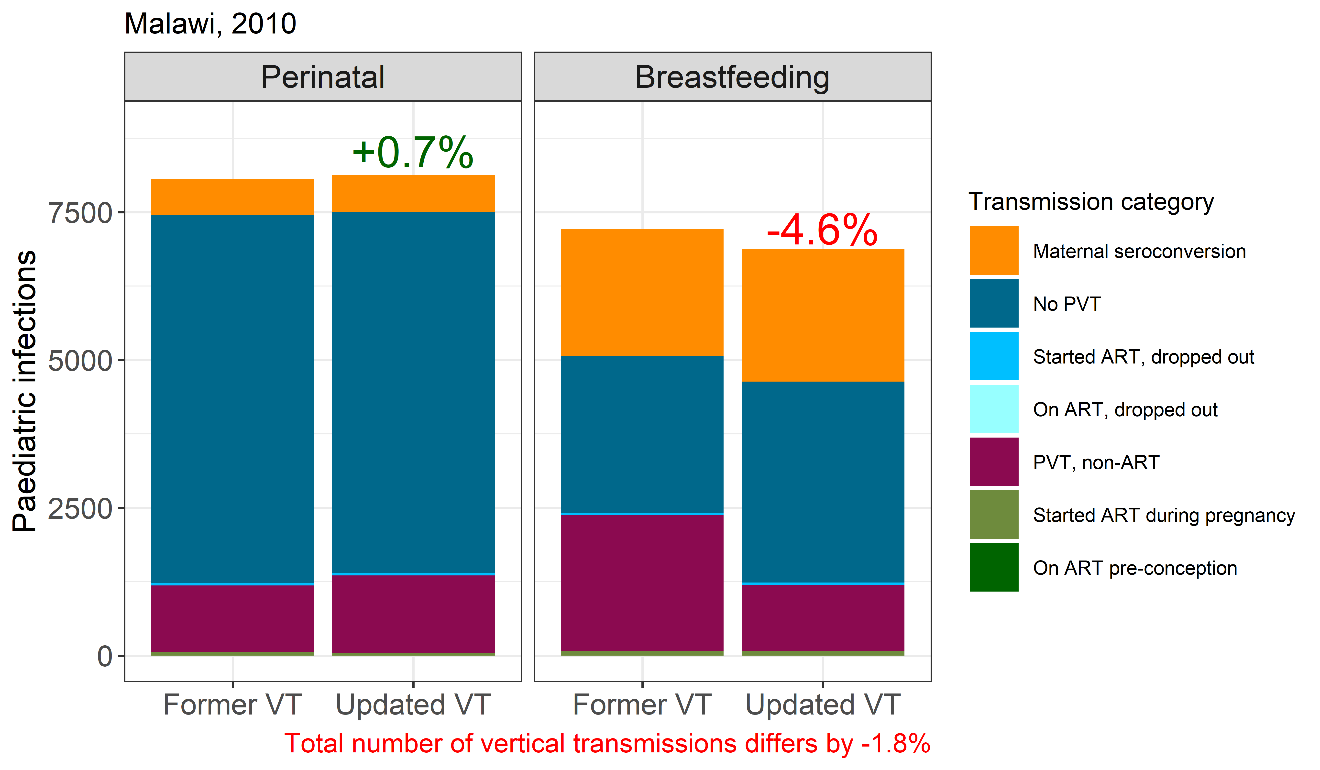

**Figure 5.2.** Change in vertical infections due to estimated vertical transmission probabilities by infection timing, Malawi 2010

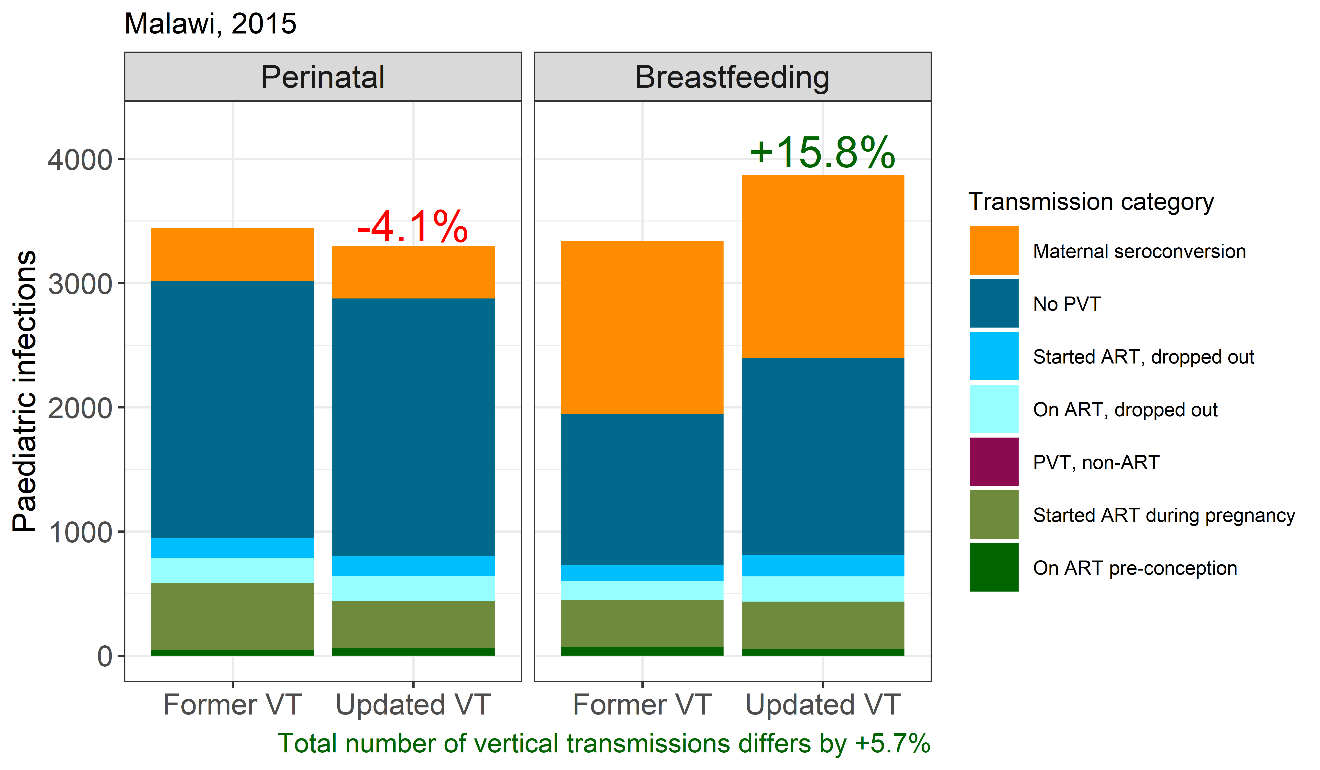

**Figure 5.3.** Change in vertical infections due to estimated vertical transmission probabilities by infection timing, Malawi 2015

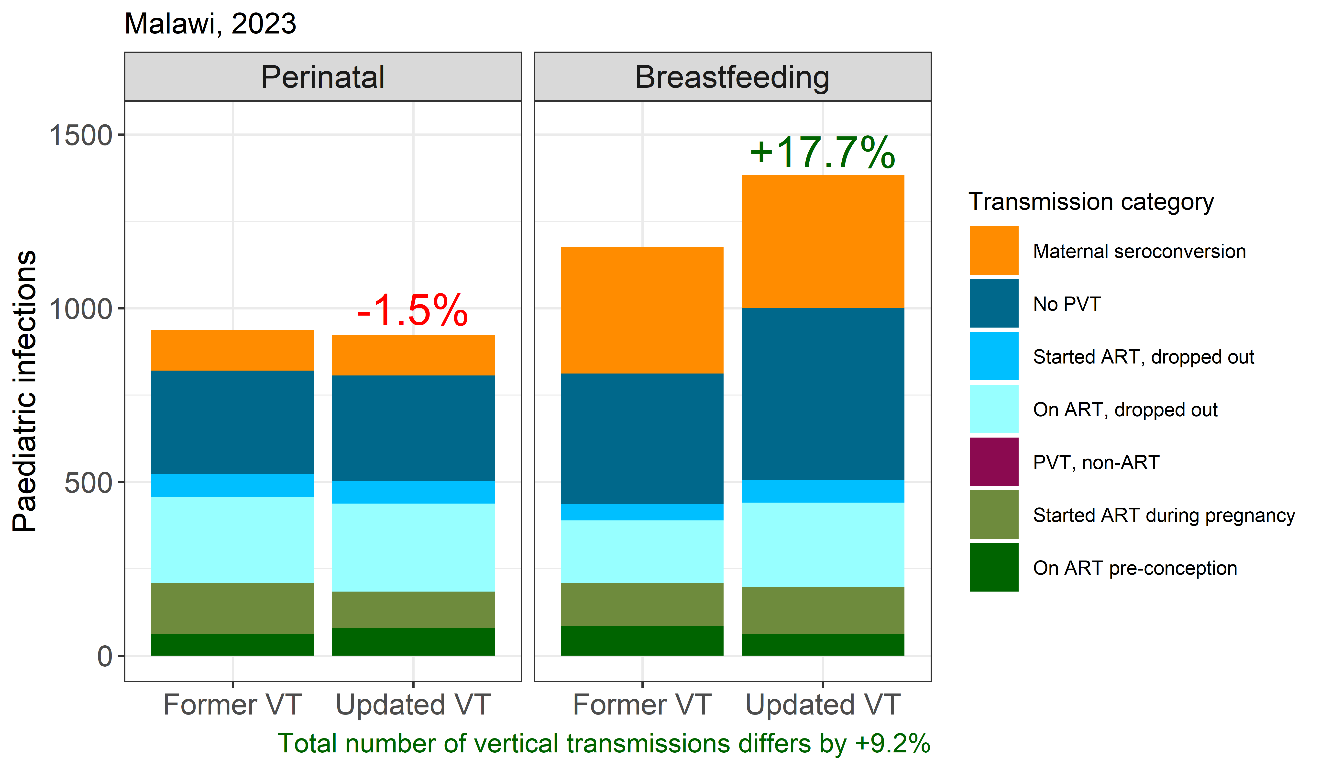

**Figure 5.4.** Change in vertical infections due to estimated vertical transmission probabilities by infection timing, Malawi 2023

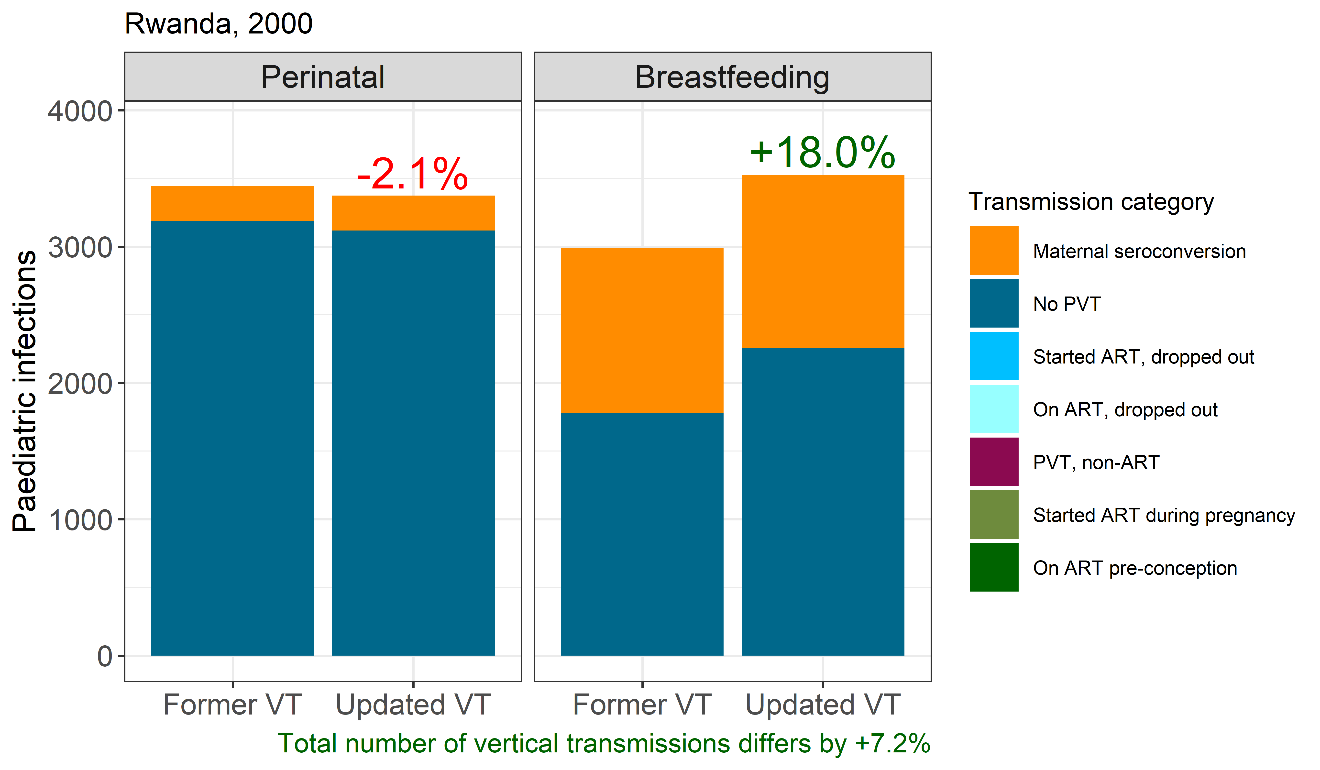

**Figure 5.5.** Change in vertical infections due to estimated vertical transmission probabilities by infection timing, Rwanda 2000

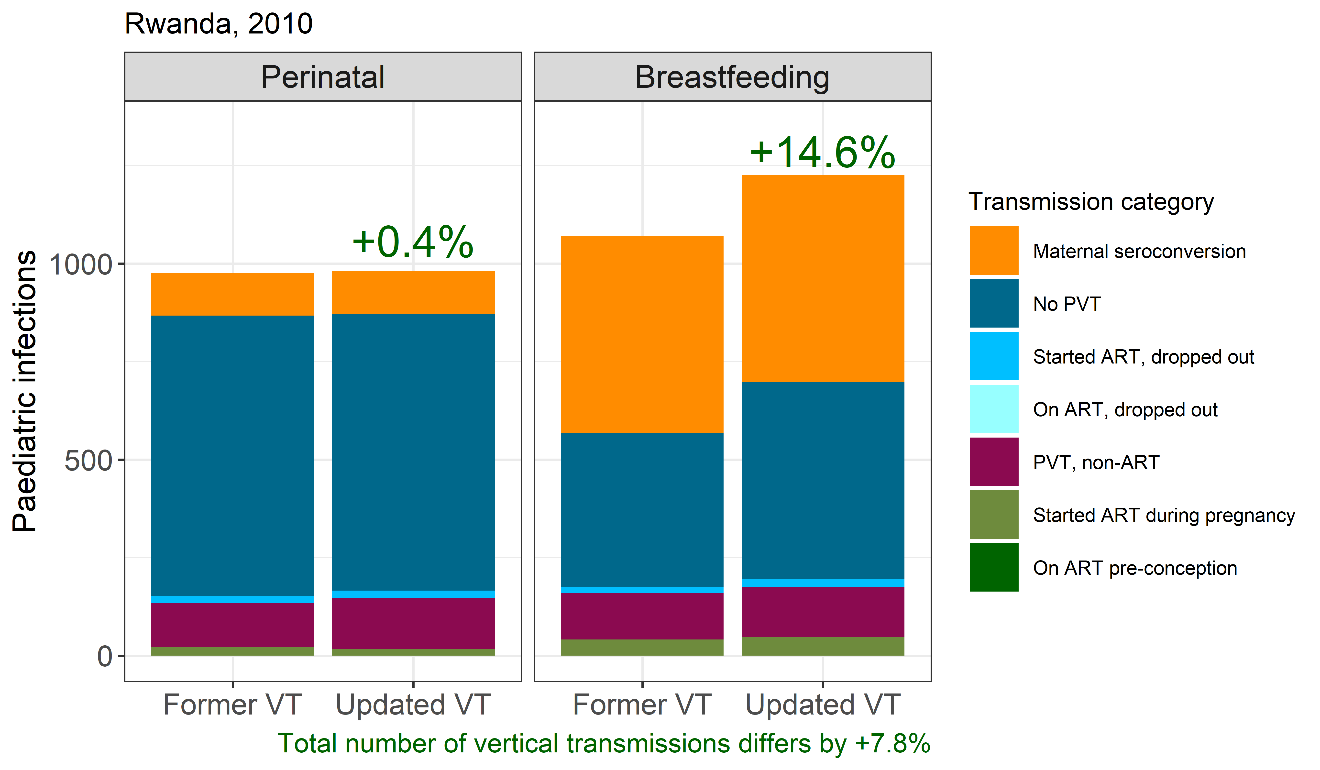

**Figure 5.5.** Change in vertical infections due to estimated vertical transmission probabilities by infection timing, Rwanda 2010

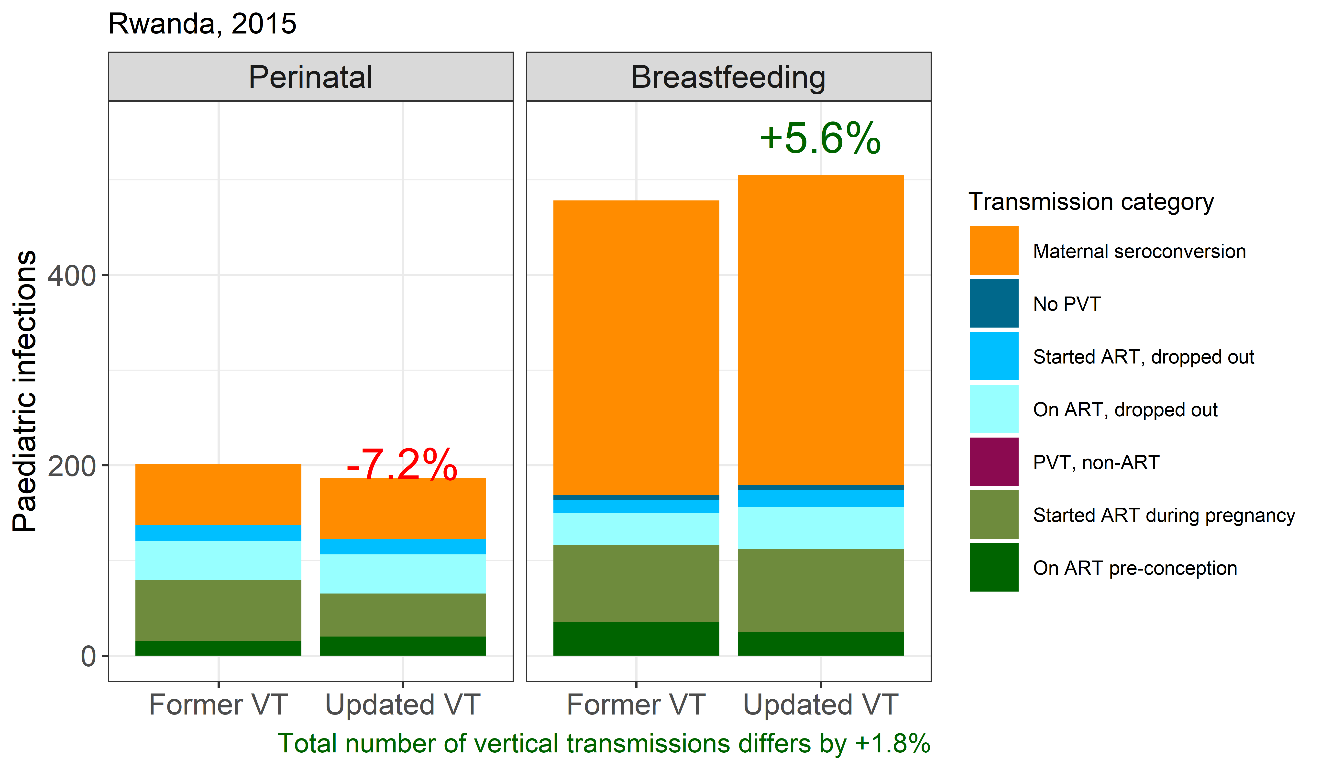

**Figure 5.7.** Change in vertical infections due to estimated vertical transmission probabilities by infection timing, Rwanda 2015

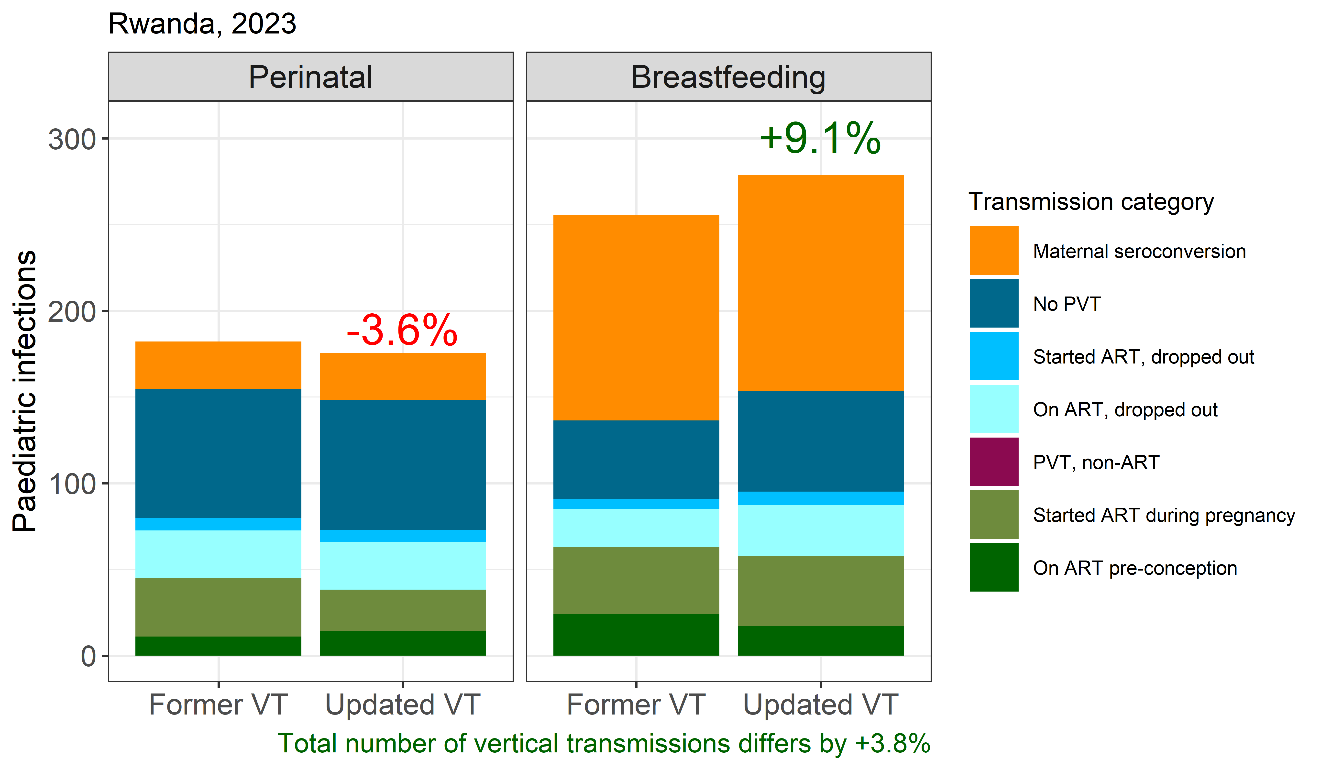

**Figure 5.8.** Change in vertical infections due to estimated vertical transmission probabilities by infection timing, Rwanda 2023

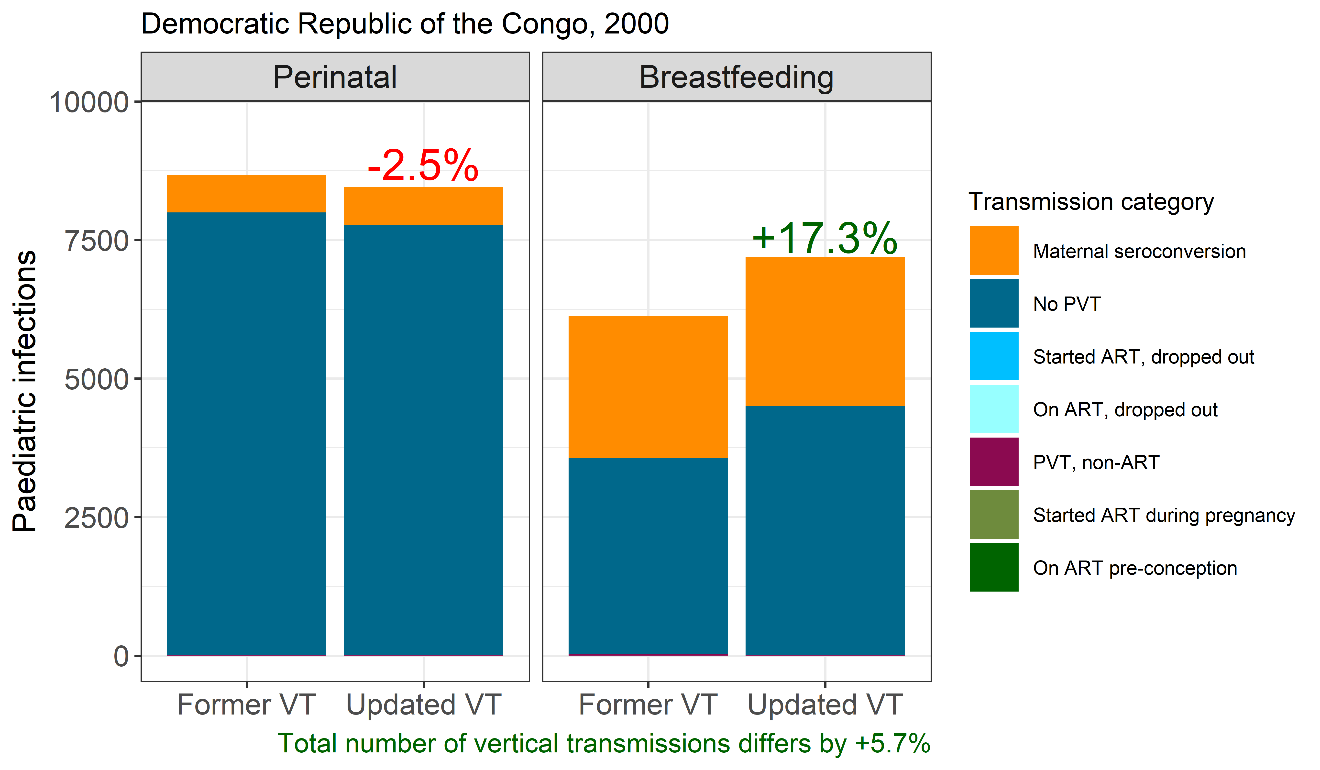

**Figure 5.9.** Change in vertical infections due to estimated vertical transmission probabilities by infection timing, Democratic Republic of the Congo 2000

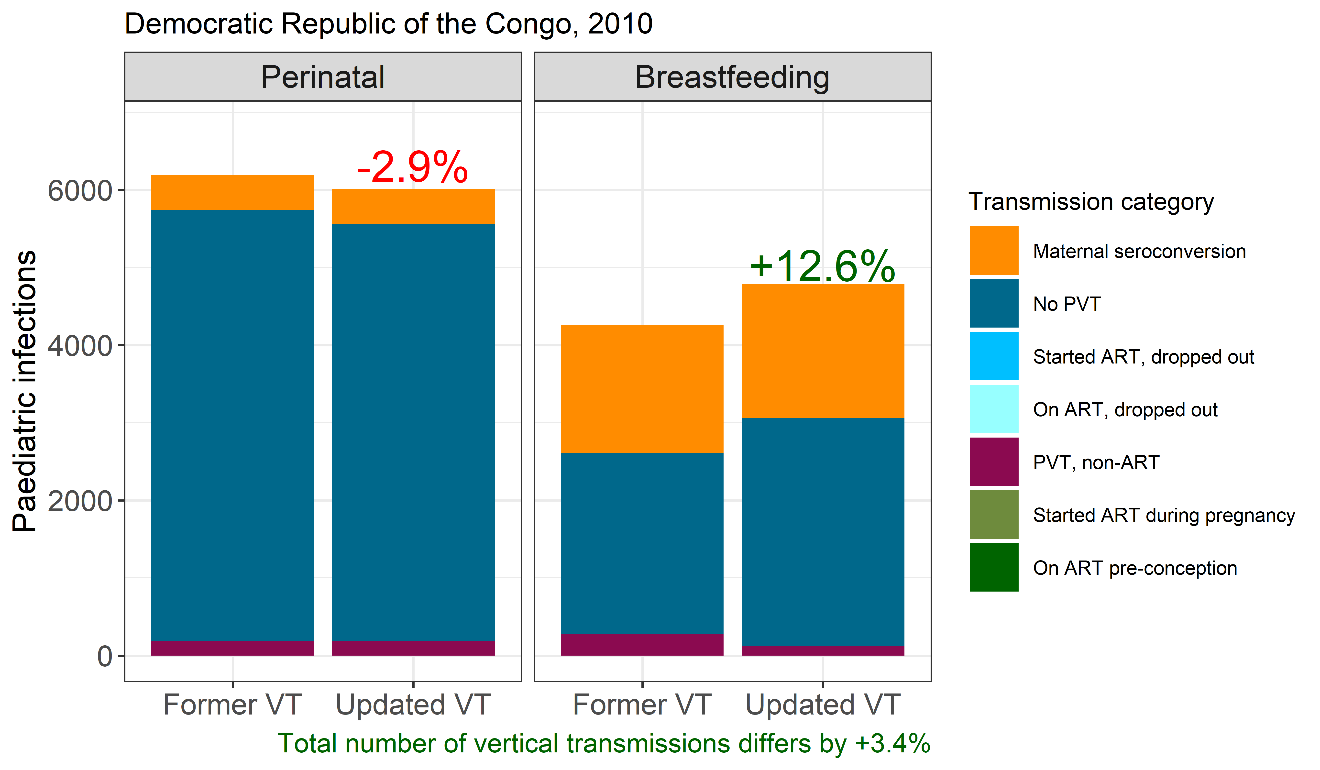

**Figure 5.10.** Change in vertical infections due to estimated vertical transmission probabilities by infection timing, Democratic Republic of the Congo 2010

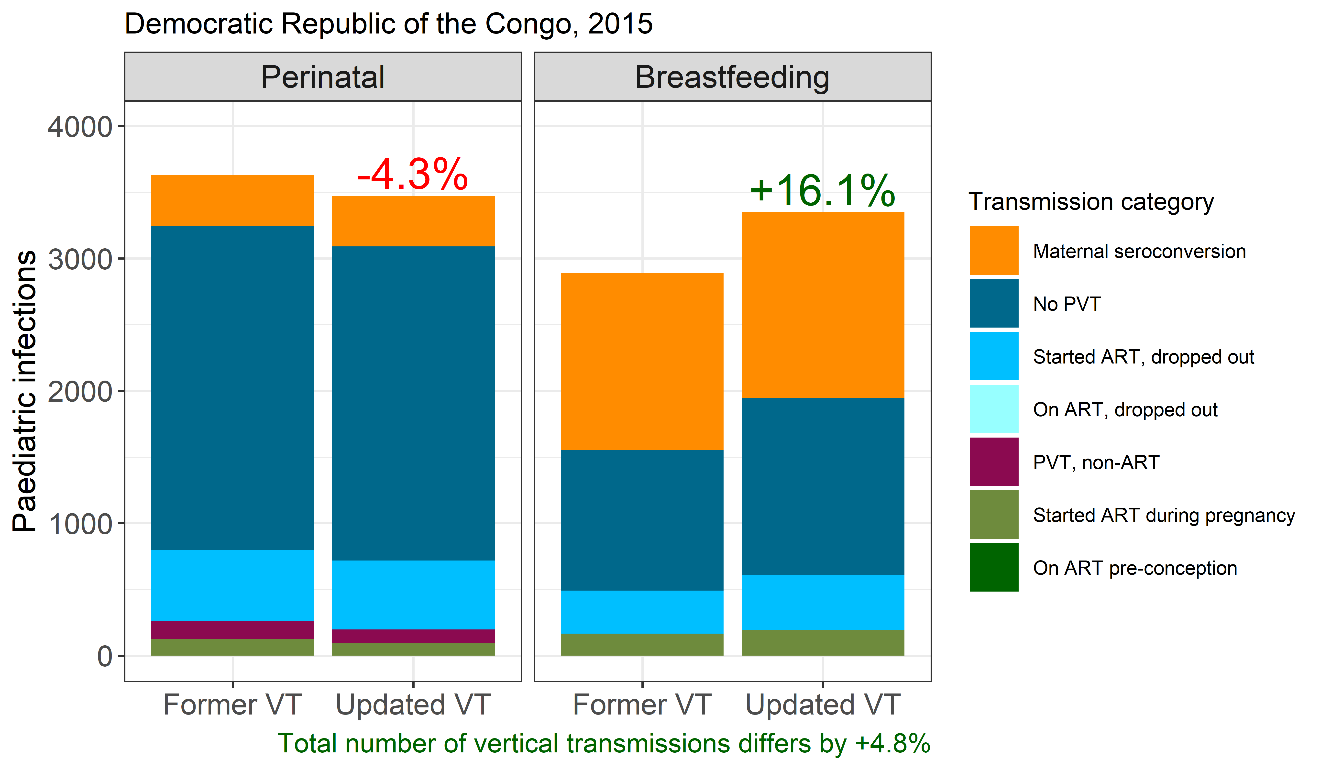

**Figure 5.11.** Change in vertical infections due to estimated vertical transmission probabilities by infection timing, Democratic Republic of the Congo 2015

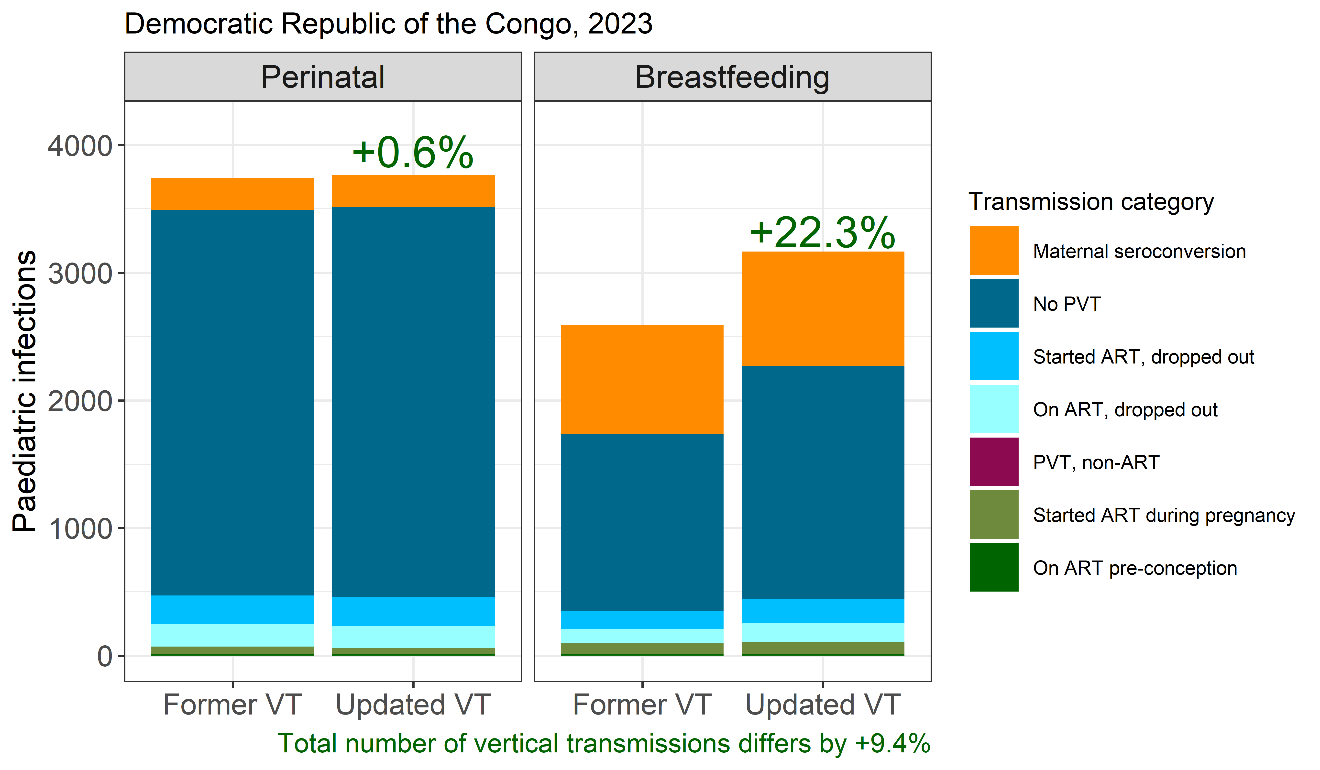

**Figure 5.12.** Change in vertical infections due to estimated vertical transmission probabilities by infection timing, Democratic Republic of the Congo 2023

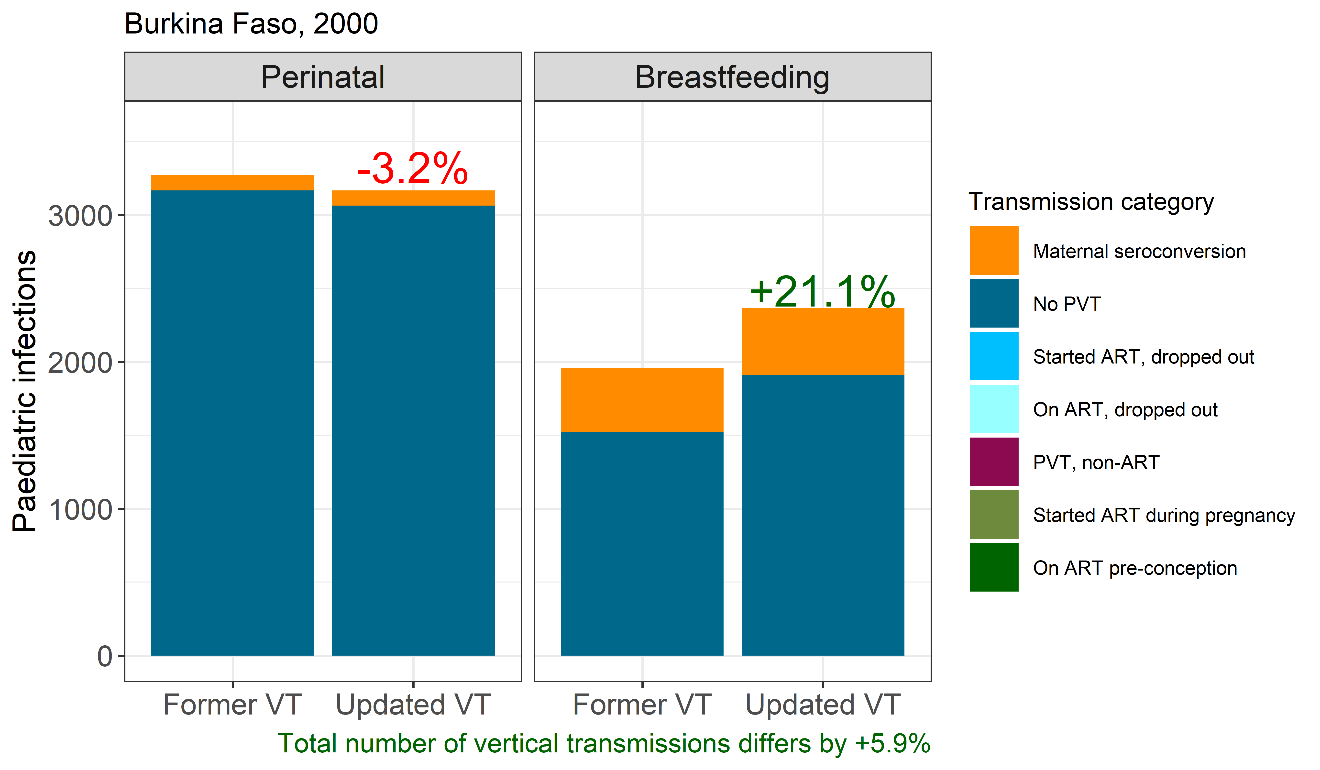

**Figure 5.13.** Change in vertical infections due to estimated vertical transmission probabilities by infection timing, Burkina Faso 2000

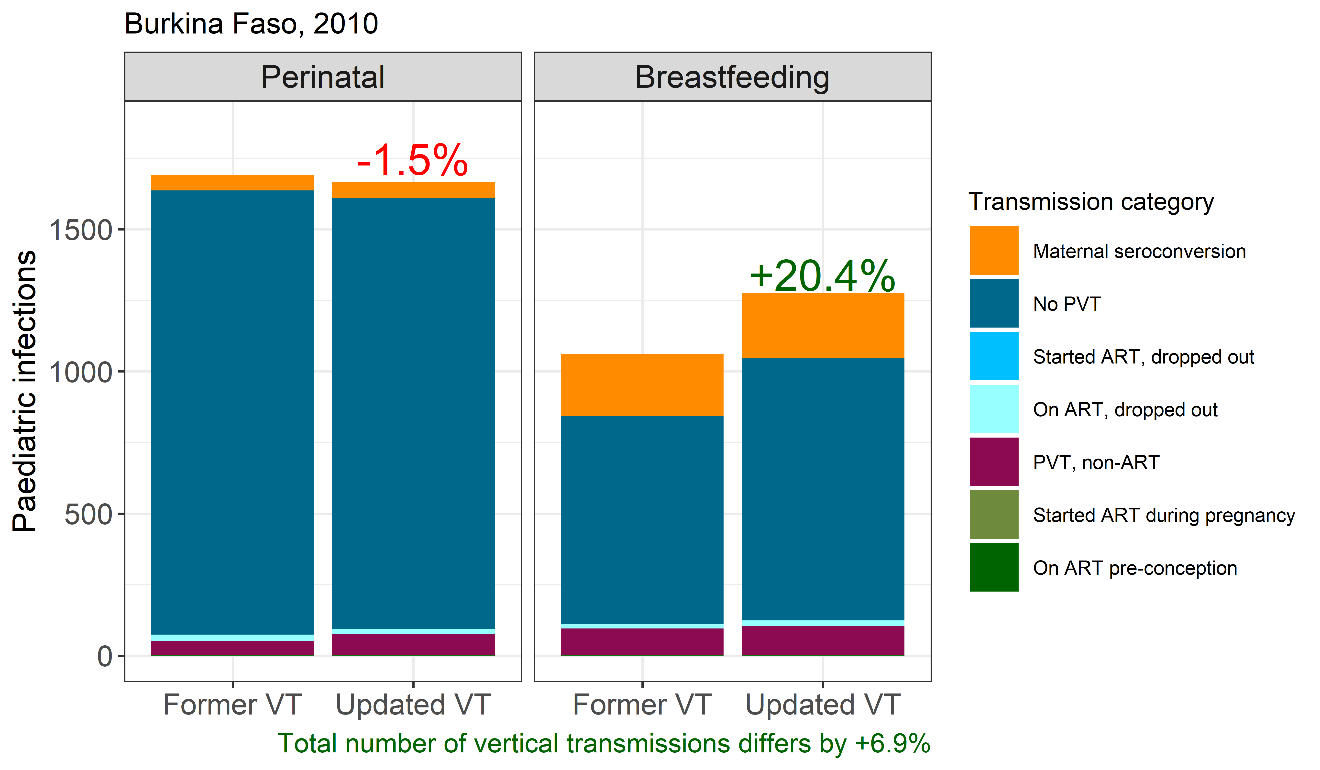

**Figure 5.14.** Change in vertical infections due to estimated vertical transmission probabilities by infection timing, Burkina Faso 2010

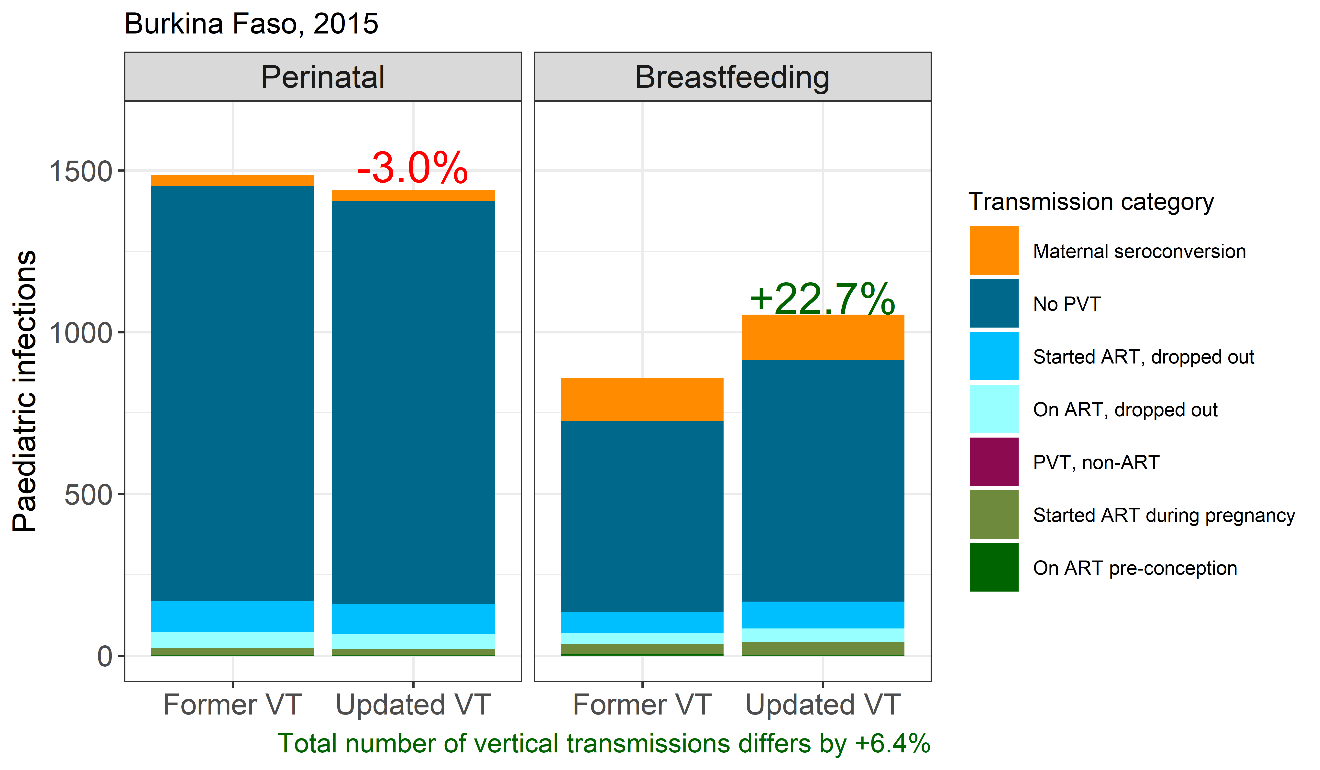

**Figure 5.15.** Change in vertical infections due to estimated vertical transmission probabilities by infection timing, Burkina Faso 2015

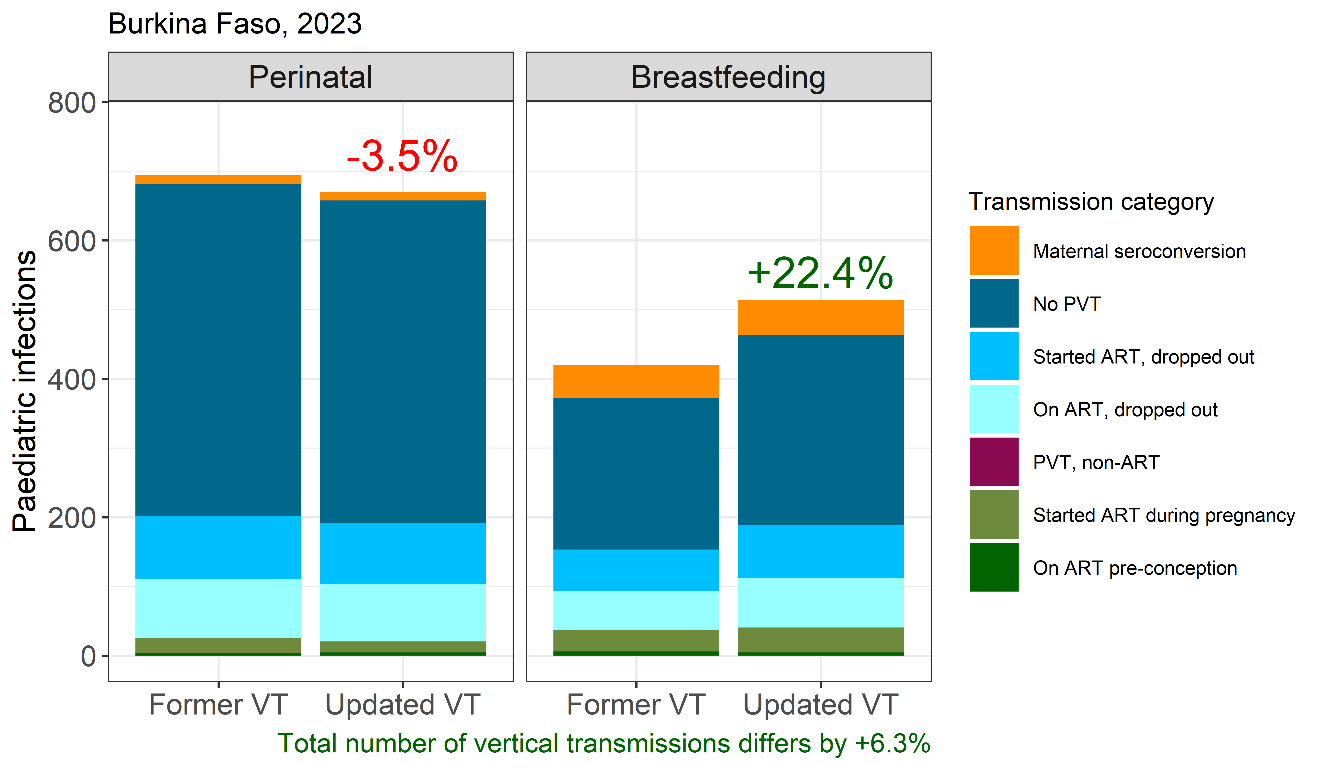

**Figure 5.15.** Change in vertical infections due to estimated vertical transmission probabilities by infection timing, Burkina Faso 2023

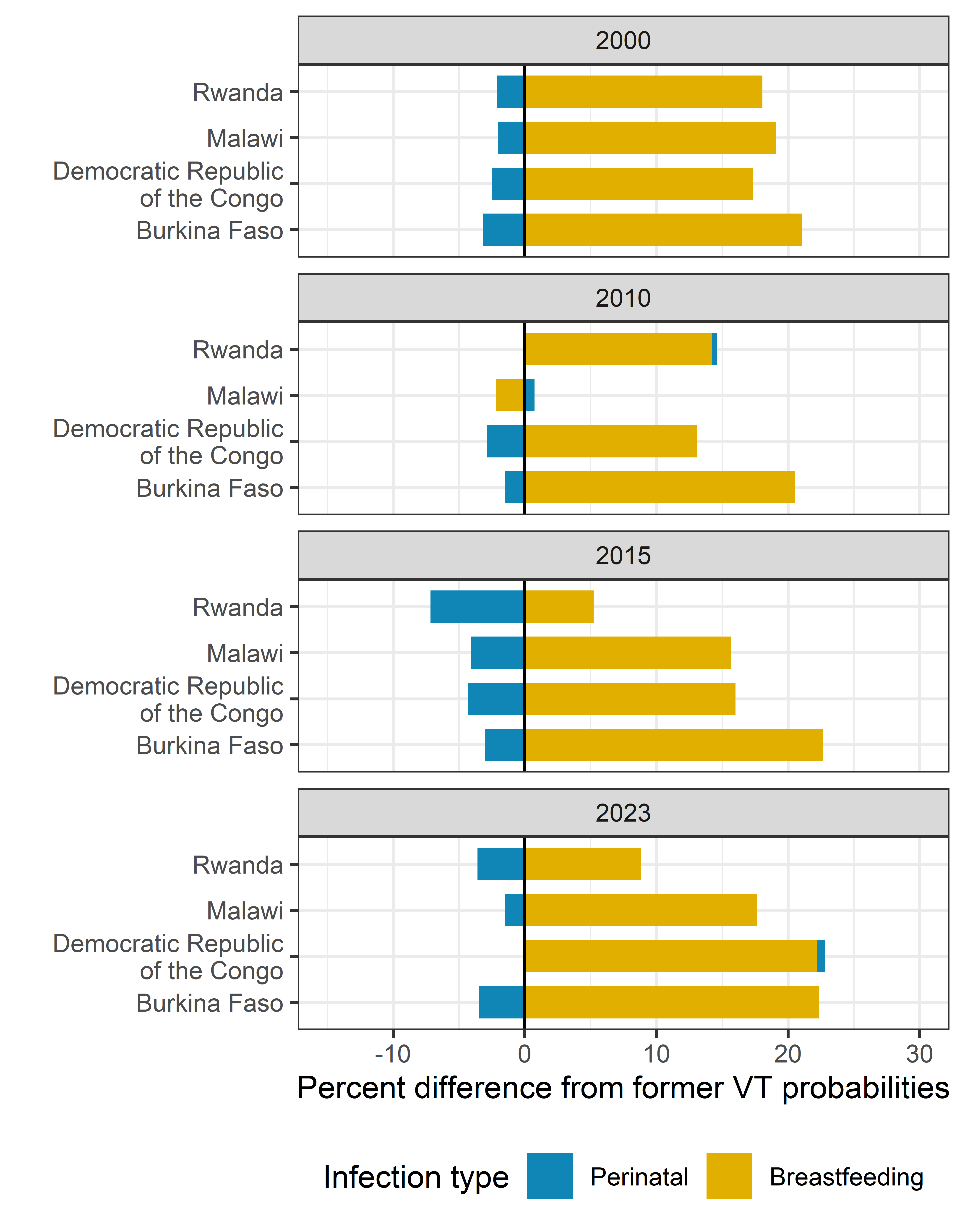

**Figure 5.17.** Summary of percent difference in the number of infections by infection type (perinatal or breastfeeding) between the former and updated vertical transmission probabilities for Rwanda, Malawi, Democratic Republic of the Congo, and Burkina Faso for the years 2000, 2010, 2015, and 2023.

### 6. Data included in meta-regression analysis

Table 6 summarises the studies that contributed data to the four meta-regression analyses on vertical transmission and to the analysis of viral load suppression at delivery. The table summarises key study characteristics (year, location, transmission timing[s] observed, preventive regimens studied, and sample size), annotates which model(s) for which each study contributed data, and whether the data were newly identified in the updated 2019-2024 search. All extracted data is also available in the supplementary file <https://github.com/mwalte10/hiv_vt_mr/blob/main/data/public_data.csv>.

**Table 6.** Study characteristics of studies included in models one through four and viral load suppression at delivery model

^1^Indicates whether the mother received PVT. If not, infection types are split into “Existing” or WLHIV who had HIV before the current pregnancy and “Infection” for mothers who seroconverted during pregnancy or breastfeeding.

^2^Yes indicates the paper was identified in the 2024 systematic review that searched for literature published between 2018-2024. No indicates the paper is from a previous review.

| ***ID*** | ***Study*** | ***Study years*** | ***Location*** | ***Maternal PVT regimen or infection type^1^*** | ***Number of HIV exposed infants*** | ***Transmission timing*** | ***Model 1:***  ***No PVT*** | ***Model 2: Maternal seroconversion and short-course PVT*** | ***Model 3: Perinatal transmission from women on ART*** | ***Model 4: Breastfeeding transmission from women on ART*** | ***Viral suppression at delivery model*** | ***Added in 2024 systematic review^2^*** |
| --- | --- | --- | --- | --- | --- | --- | --- | --- | --- | --- | --- | --- |
| **1** | **Aebi-Popp, 2022** | 2019-2021 | Switzerland | On ART | 21 | Perinatal |  |  | Included |  |  | Yes |
|  |  |  |  |  | 21 | Breastfeeding |  |  |  | Included |  |  |
| **2** | **Amone, 2023** | 2016-2017 | Uganda | Started ART | 431 | Perinatal |  |  | Included |  |  | No |
| **3** | **Bailey, 2011** | 2000-2009 | Europe | Started ART | 1,760 | Perinatal |  |  | Included |  |  | No |
| **4** | **Birkhead, 2010** | 2002-2006 | United States | Infection | 41 | Perinatal |  | Included |  |  |  | No |
| **5** | **Black, 2008** | 2004-2007 | South Africa | Started ART | 302 | Perinatal |  |  | Included |  |  | No |
| **6** | **Blonk, 2015** | 2010-2014 | Europe | On ART | 7 | Perinatal |  |  | Included |  | Included | No |
| **7** | **Bornhede, 2018** | 2014-2017 | Sweden | Started ART | 3 | Perinatal |  |  | Included |  | Included | No |
|  |  |  |  | On ART | 10 |  |  |  | Included |  | Included |  |
| **8** | **Carey, 2018** | 2008-2014 | United Kingdom | Started ART | 67 | Perinatal |  |  | Included |  |  | No |
|  |  |  |  | On ART | 65 |  |  |  | Included |  |  |  |
| **9** | **Chasela, 2010** | 2006-2008 | Malawi | Existing | 668 | Breastfeeding | Included |  |  |  |  | No |
|  |  |  |  | Option A | 748 |  |  | Included |  |  |  |  |
| **10** | **Chauhan, 2021** | 2016-2018 | India | On ART | 32 | Perinatal |  |  | Included |  |  | Yes |
| **11** | **Chen, 2019** | 2007-2015 | China | Started ART | 446 | Perinatal |  |  | Included |  |  | Yes |
| **12** | **Chibwesha, 2011** | 2007-2010 | Zambia | Option B | 250 | Perinatal |  | Included |  |  |  | No |
|  |  |  |  | Started ART | 1,813 |  |  |  | Included |  |  |  |
| **13** | **Choi, 2018** | 2005-2017 | Korea | Started ART | 8 | Perinatal |  |  | Included |  | Included | No |
|  |  |  |  | On ART | 8 |  |  |  | Included |  | Included |  |
| **14** | **Coetzee, 2019** | 2010 | South Africa | Existing | 15 | Perinatal | Included |  |  |  |  | Yes |
|  |  |  |  | Dual ARV | 8 |  |  | Included |  |  |  |  |
|  |  |  |  | Started ART | 44 |  |  |  | Included |  |  |  |
|  |  |  |  | On ART | 25 |  |  |  | Included |  |  |  |
| **15** | **Cohan, 2015** | 2009-2013 | Uganda | Option B | 353 | Breastfeeding |  | Included |  |  |  | No |
|  |  |  |  | Option B | 374 | Perinatal |  | Included |  |  |  | No |
|  |  |  |  | Started ART | 353 | Breastfeeding |  |  |  | Included |  | No |
|  |  |  |  | Started ART | 389 | Perinatal |  |  | Included |  |  | No |
| **16** | **Colbers, 2015** | Not reported | Europe | On ART | 18 | Perinatal |  |  | Included |  | Included | No |
| **17** | **Colbers, 2015** | Not reported | Europe | On ART | 11 | Perinatal |  |  | Included |  |  | No |
| **18** | **Colebunders, 1988** | 1986 | Democratic Republic of the Congo | Infection | 3 | Breastfeeding |  | Included |  |  |  | No |
| **19** | **Connor, 1994** | 1991-1993 | United States, France | Existing | 183 | Breastfeeding | Included |  |  |  |  | No |
| **20** | **Coovadia, 2013** | 2008-2011 | South Africa, Tanzania, Uganda, Zimbabwe | Existing | 434 | Breastfeeding | Included |  |  |  |  | No |
|  |  |  |  | Dual ARV | 819 |  |  | Included |  |  |  |  |
|  |  |  |  | Option A | 418 |  |  | Included |  |  |  |  |
|  |  |  |  | Single dose Nevirapine, CD4 >350 | 434 |  |  | Included |  |  |  |  |
|  |  |  |  | Single dose Nevirapine, CD4 <350 | 54 |  |  | Included |  |  |  |  |
| **21** | **Dabis, 1999** | 1995-1998 | Ivory Coast, Burkina Faso | Existing | 113 | Breastfeeding | Included |  |  |  |  | No |
|  |  |  |  |  | 145 | Perinatal | Included |  |  |  |  |  |
| **22** | **De Schacht, 2014** | 2008-2011 | Mozambique | Infection | 29 | Breastfeeding |  | Included |  |  |  | No |
| **23** | **Delicio, 2011** | 2000-2009 | Brazil | Started ART | 12 | Perinatal |  |  | Included |  |  | No |
| **24** | **Dinh, 2015** | 2011-2012 | South Africa | Infection | 212 | Perinatal |  | Included |  |  |  | No |
| **25** | **Dinh, 2018** | 2013 | Zimbabwe | Option B | 338 | Perinatal |  | Included |  |  |  | No |
|  |  |  |  | On ART | 415 |  |  |  | Included |  |  |  |
| **26** | **Dryden-Peterson, 2011** | 2009-2010 | Botswana | Option A | 170 | Perinatal |  | Included |  |  |  | No |
|  |  |  |  | Started ART | 114 |  |  |  | Included |  |  |  |
|  |  |  |  | On ART | 144 |  |  |  | Included |  |  |  |
| **27** | **Ejikunle, 2019** | 2015-2016 | Nigeria | Infection | 5 | Perinatal |  | Included |  |  |  | Yes |
| **28** | **Ekpini, 1997** | 1990-1994 | Ivory Coast | Infection | 12 | Breastfeeding |  | Included |  |  |  | No |
| **29** | **Embree, 2000** | 1986-1997 | Kenya | Infection | 12 | Breastfeeding |  | Included |  |  |  | No |
| **30** | **Ewenighi-Amankwah, 2020** | Not reported | Nigeria | On ART | 122 | Perinatal |  |  | Included |  |  | Yes |
| **31** | **Finocchario-Kessler, 2015** | 2010-2012 | Kenya | Dual ARV | 904 | Perinatal |  | Included |  |  |  | No |
|  |  |  |  | Option A | 904 |  |  | Included |  |  |  |  |
|  |  |  |  | Option B | 219 |  |  | Included |  |  |  |  |
| **32** | **Flynn, 2018** | 2011-2014 | Malawi, South Africa, Zimbabwe, Uganda, Zambia, Tanzania, India | Dual ARV | 503 | Breastfeeding |  | Included |  |  |  | No |
|  |  |  |  | Option A | 503 |  |  | Included |  |  |  |  |
|  |  |  |  | Option B | 648 |  |  | Included |  |  |  |  |
| **33** | **Frange, 2020** | 2010-2018 | France | On ART | 247 | Perinatal |  |  | Included |  | Included | Yes |
| **34** | **Ganter, 2019** | 2008-2014 | France | Started ART | 16 | Perinatal |  |  | Included |  |  | Yes |
|  |  |  |  | On ART | 78 |  |  |  | Included |  |  |  |
| **35** | **Gibb, 2012** | 2003-2009 | Uganda, Zimbabwe | On ART | 172 | Perinatal |  |  | Included |  |  | No |
|  |  |  |  |  | 172 | Breastfeeding |  |  |  | Included |  |  |
| **36** | **Gill, 2017** | 2013-2014 | Rwanda | Option B | 381 | Breastfeeding |  | Included |  |  |  | No |
|  |  |  |  | Started ART | 205 |  |  |  |  | Included |  |  |
|  |  |  |  | On ART | 381 |  |  |  |  | Included |  |  |
|  |  |  |  | Option B | 205 | Perinatal |  | Included |  |  |  |  |
|  |  |  |  | Started ART | 205 |  |  |  | Included |  |  |  |
|  |  |  |  | On ART | 381 |  |  |  | Included |  |  |  |
| **37** | **Giuliano, 2014** | 2008-2009 | Malawi | Option B | 276 | Breastfeeding |  | Included |  |  |  | No |
|  |  |  |  | Started ART | 276 |  |  |  |  | Included |  |  |
|  |  |  |  | Option B | 278 | Perinatal |  | Included |  |  |  |  |
|  |  |  |  | Started ART | 278 |  |  |  | Included |  |  |  |
| **38** | **Goga, 2015** | 2010 | South Africa | Dual ARV | 1532 | Perinatal |  | Included |  |  |  | No |
|  |  |  |  | Option A | 1532 |  |  | Included |  |  |  |  |
| **39** | **Goga, 2016** | 2011-2013 | South Africa | Dual ARV | 2113 | Perinatal |  | Included |  |  |  | No |
|  |  |  |  | Existing | 63 |  | Included |  |  |  |  |  |
|  |  |  |  | Option A | 2113 |  |  | Included |  |  |  |  |
|  |  |  |  | Option B | 890 |  |  | Included |  |  |  |  |
|  |  |  |  | Started ART | 890 |  |  |  | Included |  |  |  |
| **40** | **Goga, 2020** | 2012-2014 | South Africa | On ART | 635 | Perinatal |  |  | Included |  |  | Yes |
| **41** | **Guay, 1999** | 1997-1999 | Uganda | Single dose Nevirapine | 310 | Perinatal |  | Included |  |  |  | No |
| **42** | **Habib, 2021** | 2015-2017 | Iran | Existing | 20 | Perinatal | Included |  |  |  |  | Yes |
| **43** | **Harrington, 2019** | 2015-2016 | Malawi | Started ART | 264 | Perinatal |  |  | Included |  |  | Yes |
| **44** | **Hira, 1990** | 1985-1986 | Zambia | Infection | 19 | Breastfeeding |  | Included |  |  |  | No |
| **45** | **Hoffman, 2010** | 2004-2008 | South Africa | Existing | 23 | Perinatal | Included |  |  |  |  | No |
|  |  |  |  | Single dose Nevirapine | 1534 |  |  | Included |  |  |  |  |
|  |  |  |  | Started ART | 730 |  |  |  | Included |  |  |  |
|  |  |  |  | On ART | 143 |  |  |  | Included |  |  |  |
| **46** | **Humphrey, 2010** | 1997-2000 | Zimbabwe | Infection | 334 | Breastfeeding |  | Included |  |  |  | No |
| **47** | **Huntington, 2011** | 1996-2009 | United Kingdom | On ART | 340 | Perinatal |  |  | Included |  |  | No |
| **48** | **Iliff, 2005** | 1997-2000 | Zimbabwe | Existing | 4367 | Perinatal | Included |  |  |  |  | No |
| **49** | **João, 2012** | 2013-2018 | Argentina, Brazil, South Africa, Tanzania, Thailand, United States | Started ART | 307 | Perinatal |  |  | Included |  |  | Yes |
| **50** | **Kesho Bora, 2010** | 2005-2008 | Burkina Faso, Kenya, South Africa | Option B | 154 | Breastfeeding |  | Included |  |  |  | No |
|  |  |  |  | Single dose Nevirapine, CD4 >350 | 283 |  |  | Included |  |  |  |  |
|  |  |  |  | Single dose Nevirapine, CD4 <350 | 184 |  |  | Included |  |  |  |  |
| **51** | **Kesho Bora, 2011** | 2005-2008 | Burkina Faso, Kenya, South Africa | Option A | 284 | Perinatal |  | Included |  |  |  | No |
|  |  |  |  | Option B | 333 | Breastfeeding |  | Included |  |  |  |  |
|  |  |  |  |  | 166 | Perinatal |  | Included |  |  |  |  |
| **52** | **Kilweo, 2009** | 2004-2006 | Tanzania | Option B | 441 | Breastfeeding |  | Included |  |  |  | No |
|  |  |  |  | Started ART | 423 |  |  |  | Included |  |  |  |
|  |  |  |  | Option B | 364 | Perinatal |  | Included |  |  |  |  |
| **53** | **Kim, 2013** | 2009-2011 | Malawi | On ART | 262 | Perinatal |  |  | Included |  |  | No |
|  |  |  |  |  | 262 | Breastfeeding |  |  |  | Included |  | No |
| **54** | **Kuhn, 2010** | Not reported | Zambia | Existing | 993 | Breastfeeding | Included |  |  |  |  | No |
| **55** | **Lallemant, 2004** | 2001-2003 | Thailand | Dual ARV | 636 | Perinatal |  | Included |  |  |  | No |
|  |  |  |  | Option A | 508 |  |  | Included |  |  |  |  |
| **56** | **Le Roux, 2019** | Not reported | South Africa | Infection | 7 | Breastfeeding |  | Included |  |  |  | Yes |
| **57** | **Liang, 2009** | 2007 | China | Infection | 106 | Perinatal |  | Included |  |  |  | No |
| **58** | **Lima, 2016** | 2008-2013 | Brazil | Infection | 9 | Breastfeeding |  | Included |  |  |  | No |
| **59** | **Loh, 2021** | 2008-2015 | Singapore | Started ART | 42 | Perinatal |  |  | Included |  |  | Yes |
|  |  |  |  | On ART | 46 |  |  |  | Included |  |  |  |
| **60** | **Malaba, 2022** | 2018 | South Africa, Uganda | Started ART | 268 | Perinatal |  |  | Included |  |  | Yes |
|  |  |  |  |  | 268 | Breastfeeding |  |  |  | Included |  |  |
| **61** | **Mandelbrot, 2015** | 2000-2011 | France | Started ART | 4267 | Perinatal |  |  | Included |  | Included | No |
|  |  |  |  | On ART | 3505 |  |  |  | Included |  | Included |  |
| **62** | **Marazzi, 2010** | 2005-2009 | Malawi, Mozambique | Option B | 2528 | Breastfeeding |  | Included |  |  |  | No |
|  |  |  |  | Started ART | 2926 |  |  |  |  | Included |  |  |
|  |  |  |  | Option B | 3081 | Perinatal |  | Included |  |  |  |  |
|  |  |  |  | Started ART | 3081 |  |  |  | Included |  |  |  |
| **63** | **Marinda, 2011** | 1997-2000 | Zimbabwe | Existing | 3285 | Perinatal | Included |  |  |  |  | No |
|  |  |  |  | Infection | 422 |  |  | Included |  |  |  |  |
| **64** | **Martinson, 2007** | 2003-2005 | South Africa | Single dose Nevirapine | 108 | Perinatal |  | Included |  |  |  | No |
| **65** | **Mayaux, 1995** | 1986-1994 | France | Existing | 236 | Perinatal | Included |  |  |  |  | No |
| **66** | **Meggi, 2018** | 2014-2016 | Mozambique | Option A | 6 | Perinatal |  | Included |  |  |  | Yes |
| **67** | **Meyers, 2015** | 2010-2013 | China | Started ART | 1994 | Perinatal |  |  | Included |  |  | No |
| **68** | **Moodley, 2003** | 1999-2000 | South Africa | Single dose Nevirapine | 663 | Perinatal |  | Included |  |  |  | No |
| **69** | **Myer, 2017** | 2013-2014 | South Africa | Option B | 555 | Perinatal |  | Included |  |  | Included | No |
|  |  |  |  | Started ART | 555 |  |  |  | Included |  | Included |  |
| **70** | **Namukwaya, 2011** | 2007-2009 | Uganda | Dual ARV | 1161 | Perinatal |  | Included |  |  |  | No |
|  |  |  |  | Option A | 1161 |  |  | Included |  |  |  |  |
|  |  |  |  | Single dose Nevirapine | 367 |  |  | Included |  |  |  |  |
| **71** | **Ndarukwa, 2019** | 2014-2016 | Zimbabwe | Started ART | 841 | Perinatal |  |  | Included |  |  | Yes |
|  |  |  |  | On ART | 289 |  |  |  | Included |  |  |  |
| **72** | **Nduati, 2000** | 1992-1997 | Kenya | Existing | 165 | Breastfeeding | Included |  |  |  |  | No |
| **73** | **Nesheim, 2007** | 2001-2005 | United States | Infection | 4 | Perinatal |  | Included |  |  |  | No |
| **74** | **Ngoma, 2015** | 2008-2009 | Zambia | Option B | 219 | Perinatal |  | Included |  |  |  | No |
|  |  |  |  | Started ART | 219 |  |  |  | Included |  |  |  |
| **75** | **Nlend, 2013** | 2008-2012 | Cameroon | Option A | 110 | Perinatal |  | Included |  |  |  | No |
|  |  |  |  | Started ART | 285 |  |  |  | Included |  |  |  |
| **76** | **Nyandiko, 2010** | 2002-2007 | Kenya | Single dose Nevirapine | 69 | Perinatal |  | Included |  |  |  | No |
| **77** | **Olana, 2016** | 2006-2014 | Ethiopia | Existing | 102 | Perinatal | Included |  |  |  |  | No |
|  |  |  |  | Dual ARV | 50 |  |  | Included |  |  |  |  |
|  |  |  |  | Option A | 50 |  |  | Included |  |  |  |  |
| **78** | **Ørbæk, 2017** | 2002-2014 | Denmark | On ART | 247 | Perinatal |  |  | Included |  |  | No |
| **79** | **Palasanthiran, 1993** | 1980-1989 | Australia | Infection | 11 | Breastfeeding |  | Included |  |  |  | No |
| **80** | **Pellowski, 2019** | 2012-2015 | South African | Option A | 239 | Perinatal |  | Included |  |  |  | Yes |
| **81** | **Peltier, 2009** | 2005-2007 | Rwanda | Option B | 532 | Perinatal |  | Included |  |  |  | No |
|  |  |  |  | Started ART | 227 |  |  |  | Included |  |  |  |
|  |  |  |  |  | 227 | Breastfeeding |  |  |  | Included |  |  |
| **82** | **Perry, 2016** | 2007-2012 | United Kingdom | On ART | 178 | Perinatal |  |  | Included |  |  | No |
|  |  |  |  | Started ART | 493 |  |  |  | Included |  |  |  |
| **83** | **Peters, 2017** | 2012-2014 | United Kingdom | On ART | 1749 | Perinatal |  |  | Included |  |  | No |
| **84** | **PETRA Study Team, 2002** | 1996-2000 | Tanzania, South Africa, Uganda | Existing | 303 | Perinatal | Included |  |  |  |  | No |
|  |  |  |  |  | 303 | Breastfeeding | Included |  |  |  |  |  |
| **85** | **Prieto, 2012** | 2000-2007 | Spain | Existing | 68 | Perinatal | Included |  |  |  |  | No |
|  |  |  |  | Started ART | 244 |  |  |  | Included |  |  |  |
| **86** | **Rollins, 2007** | 2004-2005 | South Africa | Infection | 172 | Perinatal |  | Included |  |  |  | No |
| **87** | **Roongpisuthipong, 2001** | 1992-1994 | Thailand | Infection | 15 | Perinatal |  | Included |  |  |  | No |
| **88** | **Sagna, 2015** | 2009-2013 | Burkina Faso | Option A | 136 | Perinatal |  |  | Included |  |  | No |
| **89** | **Salazar-Austin, 2018** | 2011-2014 | South Africa | Dual ARV | 48 | Perinatal |  |  | Included |  |  | No |
|  |  |  |  | Option A | 48 |  |  |  | Included |  |  |  |
|  |  |  |  | Option B | 150 |  |  |  | Included |  |  |  |
|  |  |  |  | Option B | 171 | Breastfeeding |  |  | Included |  |  |  |
| **90** | **Samuel, 2014** | 2004-2010 | United Kingdom | On ART | 68 | Perinatal |  |  | Included |  | Included | No |
| **91** | **Schalkwijk, 2017** | Not reported | Europe | On ART | 15 | Perinatal |  |  | Included |  | Included | No |
| **92** | **Scott, 2017** | 2002-2009 | United States | Started ART | 44 | Perinatal |  |  | Included |  |  | No |
| **93** | **Shaffer, 1999** | 1996-1997 | Thailand | Existing | 195 | Perinatal | Included |  |  |  |  | No |
| **94** | **Shapiro, 2006** | 2005-2006 | Botswana | Dual ARV | 345 | Perinatal |  | Included |  |  |  | No |
|  |  |  |  | Option A | 345 |  |  | Included |  |  |  |  |
| **95** | **Shapiro, 2010** | 2006-2008 | Botswana | Option B | 553 | Perinatal |  | Included |  |  |  | No |
|  |  |  |  | Option B | 703 | Breastfeeding |  | Included |  |  |  |  |
|  |  |  |  | Started ART | 480 |  |  |  |  | Included |  |  |
| **96** | **Sibiude, 2023** | 2000-2017 | France | Started ART | 7448 | Perinatal |  |  | Included |  | Included | No |
|  |  |  |  | On ART | 6606 |  |  |  | Included |  | Included |  |
| **97** | **SWEN Study Team, 2008** | 2001-2007 | Ethiopia, Uganda, India | Single dose Nevirapine | 986 | Perinatal |  | Included |  |  |  | No |
| **98** | **Thomas, 2011** | 2003-2009 | Kenya | Option B | 487 | Perinatal |  | Included |  |  |  | No |
|  |  |  |  | Started ART | 487 |  |  |  | Included |  |  |  |
|  |  |  |  | Option B | 522 | Breastfeeding |  | Included |  |  |  |  |
|  |  |  |  | Started ART | 457 |  |  |  |  | Included |  |  |
| **99** | **Tiam, 2019** | 2014-2016 | Lesotho | Started ART | 370 | Perinatal |  |  | Included |  |  | Yes |
|  |  |  |  | On ART | 249 |  |  |  | Included |  |  |  |
| **100** | **Tonwe-Gold, 2007** | 2003-2005 | Ivory Coast | Option A | 122 | Perinatal |  | Included |  |  |  | No |
|  |  |  |  | Option B | 52 | Breastfeeding |  | Included |  |  |  |  |
|  |  |  |  | Single dose Nevirapine, CD4 >350 | 86 |  |  | Included |  |  |  |  |
| **101** | **Tookey, 2016** | 2003-2013 | United Kingdom | Started ART | 2905 | Perinatal |  |  | Included |  | Included | No |
|  |  |  |  | On ART | 968 |  |  |  | Included |  | Included |  |
| **102** | **Torpey, 2012** | 2007-2010 | Zambia | Option A | 2366 | Perinatal |  | Included |  |  |  | No |
|  |  |  |  | Single dose Nevirapine | 1143 |  |  | Included |  |  |  |  |
| **103** | **Tovo, 1991** | 1980-1989 | Italy | Infection | 10 | Perinatal |  | Included |  |  |  | No |
| **104** | **Townsend, 2014** | 2000-2011 | United Kingdon, Ireland | Existing | 54 | Perinatal | Included |  |  |  |  | No |
|  |  |  |  | Started ART | 3422 |  |  |  | Included |  |  |  |
|  |  |  |  | On ART | 2105 |  |  |  | Included |  |  |  |
| **105** | **Tubiana, 2013** | 2007-2010 | France | Started ART | 36 | Perinatal |  |  | Included |  | Included | No |
| **106** | **Van de Perre, 1991** | 1988 | Rwanda | Infection | 15 | Breastfeeding |  | Included |  |  |  | No |
| **107** | **Van Schalkwyk, 2013** | 2008-2010 | South Africa | Started ART | 127 | Perinatal |  |  | Included |  |  | No |
| **108** | **Wiktor, 1999** | 1996-1998 | Ivory Coast | Existing | 119 | Perinatal | Included |  |  |  |  | No |
|  |  |  |  |  | 115 | Breastfeeding | Included |  |  |  |  |  |
| **109** | **Yusuf, 2022** | Not reported | United States | On ART | 10 | Perinatal |  |  | Included |  | Included | Yes |
|  |  |  |  |  | 9 | Breastfeeding |  |  |  | Included |  |  |
| **110** | **Zijenah, 2022** | 2017-2018 | Zimbabwe | Started ART | 179 | Perinatal |  |  | Included |  | Included | Yes |
|  |  |  |  | On ART | 272 |  |  |  | Included |  | Included |  |
|  |  |  |  | Started ART | 61 | Breastfeeding |  |  |  | Included |  |  |

#### 6.1 Study references

**18.** Colebunders R. Breastfeeding and transmission of HIV. *The Lancet* 1988; : 1487.

**24**

Dinh T-H, Delaney KP, Goga A, *et al.* Impact of maternal HIV seroconversion during pregnancy on early mother to child transmission of hiv (MTCT) measured at 4-8 weeks postpartum in South Africa 2011-2012: a national population-based evaluation. *PLOS ONE* 2015; **10**: e0125525.

**102**

Torpey K, Mandala J, Kasonde P, *et al.* Analysis of HIV early infant diagnosis data to estimate rates of perinatal HIV transmission in Zambia. *PLOS ONE* 2012; **7**: e42859.
